# Emulation of placebo-controlled index trials using observational data with cloning, censoring and weighting: Empirical assessment of constraints and credibility

**DOI:** 10.1101/2025.10.20.25337820

**Authors:** Anna-Janina Stephan, Gerard Portela, Raisa Levin, Nils Krüger, Sebastian Schneeweiss, Rishi J. Desai

## Abstract

**Objective:** Target trial emulation (TTE) has become a prominent approach to conducting observational effectiveness studies, yet limited attention has been paid to the nuances of emulating placebo-controlled trials in this framework using claims data. As a demonstration, we aimed to expand evidence generated by the TOPCAT trial comparing spironolactone versus placebo in patients with heart failure with preserved ejection fraction (HFpEF) to the U.S. HFpEF population.

**Methods and Analysis:** We estimated the observational analogue of the per-protocol effect for spironolactone initiation and continued use versus non-initiation in 2012-2020 Medicare claims with the clone-censor-weight approach. We evaluated two composite effectiveness endpoints of heart failure hospitalization (HHF) and cardiac arrest with either all-cause or cardiovascular mortality, respectively, as well as each component except cardiac arrest as an individual endpoint. Anticipating threats to validity through residual confounding, we pre-specified two guardrails: 1) benchmarking against results from TOPCAT Americas, and 2) evaluation of non-cardiovascular mortality as negative control outcome to quantify and correct for the magnitude of residual bias. To demonstrate investigator-induced biases frequently seen in studies not using the TTE framework, we additionally implemented a ‘naïve’ ever- vs never-user comparison that misclassified immortal person-time before spironolactone initiation as exposed.

**Results:** We included 320,881 patients with HFpEF in the overall Medicare cohort (mean age 80.6 years (SD 8.37); female 62%), of which 49,729 qualified for benchmarking against TOPCAT. In the benchmarking cohort, relative risks with spironolactone use compared to non-use for effectiveness outcomes ranged between 0.97 (95%-CI = [0.94; 1.01]) for the composite with cardiovascular death and 1.14 (95%-CI = [1.11; 1.18]) for all-cause mortality. The negative control of non-cardiovascular mortality suggested presence of residual confounding. After bias correction, our relative risks were in line with TOPCAT hazard ratios for HHF-driven outcomes (e.g. composite with cardiovascular death 0.88 (95%-CI = [0.85; 0.91]) in our study vs. 0.82 (95%-CI = [0.69; 0.98]) in TOPCAT), but not for mortality outcomes (e.g. all-cause death 1.04 (95%-CI = [1.01; 1.07]) vs. 0.83 (95%-CI = [0.68; 1.02]) in TOPCAT). Estimates in the overall cohort were comparable to the benchmarking cohort. The naïve analysis of ever versus never-use produced substantially biased results (e.g. 1.22 (95%-CI = [1.13; 1.30], composite with cardiovascular death) to 0.58 (95%-CI = [0.53; 0.65], all-cause death, benchmarking cohort).

**Conclusion:** In emulations of placebo-controlled trials, residual confounding remains a persistent threat and it is critical to build in pre-specified guardrails to detect and address this bias.

**Key messages:**

- **What is already known on this topic** – Target trial emulation presents a principled framework of designing observational studies, and within this framework, the clone-censor-weight approach has been recommended to avoid immortal time bias when emulating placebo-controlled trials.
- **What this study adds** – Even after fully avoiding immortal time through the clone-censor-weight approach within the target trial framework, observational studies of non-use comparisons remain prone to other sources of bias. Bias analysis and benchmarking can help gauge the extent and direction of such bias.
- **How this study might affect research, practice or policy** – This study showcases how researchers can leverage pre-specified benchmarking and net bias analysis as guardrails when using the clone-censor-weight design for non-use-comparisons to ensure accurate interpretation. It also provides auxiliary evidence on the effects of spironolactone in HFpEF for the Medicare population beyond TOPCAT that may inform clinical decision-making.

## Introduction

In many clinical scenarios without an established standard of care treatment, a question of high clinical relevance is whether a treatment improves outcomes compared to no treatment. Randomized controlled trials (RCTs) address this question by comparing outcomes between groups assigned to receive either an experimental treatment or placebo^1^. The function of placebo control in this setting is to eliminate subjectivity in outcome assessment. However, investigators may additionally want to assess the effectiveness of such treatments without an established alternative in a broader population beyond the original RCT, or where no such RCT exists. In these cases, a common approach is the analysis of observational data generated during provision of routine clinical care^2^.

A limitation of routine care data is that it does not comprise of placebo controls as potential comparators, and in some cases no other suitable active comparator may exist. In this situation, investigators may turn towards a non-user comparison, an observational study design that is prone to known challenges^3,4^. Specifically, such comparisons can induce immortal time bias because of challenges in defining a clear inception point (“time zero”) from which to start follow-up for the non-use strategy. Target trial emulation with cloning, censoring and weighting is a principled approach to avoid immortal time bias in studies where treatment strategies are indistinguishable at time zero^5^. Briefly, with clone-censor-weighting, investigators ‘assign’ replicates (i.e., clones) of each observation to all treatment strategies of interest at time zero and thereby begin follow-up on a pre-defined common index time that is identifiable in all treatment arms. A clone is subsequently censored when their data becomes incompatible with the assigned strategy. The final weighting step remediates the informativeness of artificial censoring in the previous step. The most appealing feature of using the clone-censor-weight approach within the target trial emulation framework is that it allows investigators to study a well-defined causal question that could be addressed by a corresponding RCT.

Although the target trial emulation framework has become a prominent approach to conduct observational studies, very limited attention has been paid to the nuances of emulating placebo-controlled trials in this framework. Specifically, non-user comparisons may still be especially prone to confounding even when investigator-induced designs flaws are avoided. As a demonstrative use case, we aimed to expand the evidence generated by the “Treatment of Preserved Cardiac Function Heart Failure with an Aldosterone Antagonist” (TOPCAT) trial that evaluated efficacy of spironolactone, a mineralocorticoid receptor antagonist (MRA), against a placebo comparison in heart failure with preserved ejection fraction (HFpEF)^7^. Post hoc subgroup analyses for TOPCAT’s American study sites suggested a reduced risk of hospitalization for heart failure (HHF, hazard ratio (HR) 0.91 (95%-CI = [0.78; 1.06]) cardiovascular death (HR=0.74, 95%-CI = [0.57; 0.97]), all-cause death (HR=0.83, 95%-CI = [0.68; 1.02]), and the primary composite endpoint of HHF, cardiac arrest and cardiovascular death (HR=0.82, 95%-CI = [0.69; 0.98]).

We aimed to evaluate effectiveness of spironolactone in the overall U.S. HFpEF population beyond the strict eligibility criteria of TOPCAT, and in subgroups characterized by different HFpEF phenotypes and socio-demographic characteristics. We emulated a target trial using cloning, censoring and weighting in the nationwide population of U.S. Medicare enrolees with HFpEF between 2012-2020 to compare outcomes of initiation and sustained spironolactone use against non-use. As there was no definitive standard of care treatment strategy for HFpEF in the study period, we followed the TOPCAT placebo comparison rationale which suggested that the most clinically relevant comparator in this population would be non-use. Anticipating potential threats to validity due to unmeasured differences between active treatment and non-use groups, we pre-specified two mechanisms to detect net bias^8^ as guardrails: 1) emulating the design and population of an index trial (TOPCAT) as closely as possible to benchmark the results, 2) adding a negative control outcome of non-cardiovascular mortality to the outcomes of interest to quantify and correct for the magnitude of residual bias.

## Materials and Methods

In the following, we first describe the target trial and its respective emulation in observational data for spironolactone in HFpEF as a demonstrative use case. Subsequently, we describe the guardrails we pre-specified to assess anticipated threats to validity due to design-related biases and confounding in this observational non-use comparison.

### Demonstrative use case: Real-world effectiveness of spironolactone in HFpEF

We followed recent guidelines for the planning, implementation and reporting of real-world evidence studies (PRINCIPLED^8^, HARPER^9^, STaRT-RWE^10^, TARGET^11^). A pre-specified protocol (Registration DOI: https://doi.org/10.17605/OSF.IO/6MNCF) was prospectively registered on the Open Science Framework RWE Registry. Detailed information, including minor protocol adjustments^12^ can be found on the associated OSF project page (osf.io/4rqbd). Patients or the public were not involved in the design, or conduct, or reporting, or dissemination plans of our research. The study was approved as exempt by the Mass General Brigham Institutional Review Board (2024P001297).

Table 1 outlines the protocol of the pragmatic target trial and the emulation that was implemented using real-world observational data.

**Table 1:** Target trial emulation table for the hypothetical trial.

| **Element** | **Hypothetical Target Trial Specification** | **Target Trial Emulation using real world data sources** |
| --- | --- | --- |
| **Eligibility criteria** | Individuals with HFpEF and at least one heart failure hospitalization in the six months before baseline, aged $\geq 65$ years and treated in the US between 01/2012 and 12/2020 <u>without contra-indication for spironolactone treatment.</u> | Same as target trial. |
| **Treatment strategies** | Treatment of interest: Initiation within the first 6 months after a qualifying HF diagnosis, and subsequent continuous treatment with spironolactone unless contraindications arise or a surgical treatment removes the spironolactone indication, after which patients are free to choose to discontinue or remain on spironolactone<br>Comparator: Initiation of placebo at qualifying HF diagnosis and subsequent continuous treatment with placebo thereafter, unless contraindications arise or a surgical treatment removes the potential spironolactone indication, after which patients are free to choose to initiate spironolactone or not. | Treatment of interest: Same as target trial. Proxies for contraindications: lithium prescription fills and hyperkalemia hospitalization. Proxies for surgical treatments removing the spironolactone indication: ESRD, heart transplant, VAD transplant.<br>Comparator: Spironolactone non-initiation at qualifying HF diagnosis and subsequent continued abstention, unless contraindications arise or a surgical treatment removes the potential spironolactone indication, after which patients are free to choose to initiate or spironolactone or not |
| **Treatment assignment** | Randomized, blinded. | Non-blinded and assumed to be as randomized within levels of measured confounders*.<br>Individuals with baseline data that are compatible with both strategies will be cloned and assigned to both strategies until they deviate to avoid immortal time bias. |
| **Follow-up start (time 0)** | At assignment. | Same as target trial. |
| **Follow-up end** | Development of outcome of interest, death (where death is not the outcome of interest) or loss-to-follow-up, whichever occurs first. | Same as target trial.<br>Additional data-base specific follow-up ends include nursing home or |
|  |  | hospice placement and disenrollment, and end of database. |
| **Outcomes** | Time to <u>cardiovascular</u> mortality, aborted cardiac arrest or hospitalization for the management of heart failure, as a composite.<br>Time to <u>all-cause</u> mortality, aborted cardiac arrest or hospitalization for the management of heart failure, as a composite.<br>All single components of the above composite outcomes except aborted cardiac arrest.<br>non-cardiovascular mortality (negative control)<br>hyperkalemia (safety outcome) | Same as target trial, with restricted data availability period for outcomes including cause-specific mortality (secondary objectives). |
| **Causal contrast** | Intention-to-treat effect (effect of being assigned to the treatment) and per-protocol effect (effect of adhering to treatment). | An observational analogue of a per protocol or as treated effect since we will censor people as soon as they deviate from their assigned treatment, unless this treatment deviation is in line with the definition of treatment strategies. |
| **Data analysis** | Pooled logistic model with stabilized truncated inverse probability of treatment and censoring weights using time-invariant and time-varying covariates to adjust for selection bias through censoring over time. | Same as target trial. |

### Hypothetical target trial

Trial participants would be 65 years or older, with prevalent HFpEF, and at least one previous hospitalization for heart failure (outside the context of a myocardial infarction) in the 180 days preceding trial start. Exclusion criteria would comprise any history of heart or left ventricular assist device (LVAD) transplant, or end-stage renal disease (ESRD), and prescriptions of lithium, spironolactone or other potassium-sparing medications in the 180 days before trial start.

Trial participants would be randomized to either initiation and continued spironolactone use or to placebo. Both treatment strategies would allow for deviation from the assigned strategy at physician discretion after intercurrent events that pose either clinical contraindications to spironolactone use (a hyperkalemia diagnosis leading to hospitalization, an ESRD diagnosis, prescription of lithium) or make it potentially redundant (LVAD or heart transplant).

Effectiveness outcomes of interest would be 12-month relative risk and risk difference for 1) first HF hospitalization, 2) all-cause death, 3) cardiovascular death, and two composite endpoints consisting of first HF hospitalization, aborted cardiac arrest and either 4) all-cause or 5) cardiovascular mortality, respectively. Secondarily, hyperkalemia hospitalization would be evaluated as a safety endpoint. Rather than identifying the intention-to-treat effect of a simple point intervention (initiation compared to no initiation), the analysis would aim at identifying the per-protocol effect, censoring individuals at deviation from their assigned treatment strategy.

#### Covariates

Because per-protocol analysis breaks the randomization, we identified 143 time-constant potential confounders that comprise socio-demographic information as well as information on comorbidities, frailty status, medication fills, health care utilization and preventive services use. Of these, a subset related to health care utilization (number of prescribed medications, prescribing and treating physicians, outpatient, emergency department, cardiologist and hospital visits for any reason, number of heart failure hospitalizations), and to prescription fills (angiotensin-converting enzyme inhibitors, angiotensin receptor blockers, beta blockers, calcium channel blockers, loop diuretics, thiazide diuretics, hydralazine, nitrates, digoxin, sacubitril/valsartan, and sodium-glucose co-transporter 2 inhibitors) would additionally be considered as time-varying covariates. These would be updated every month to account for potential treatment-confounder feedback in the per-protocol analysis and to emulate complete observations over time without loss-to follow-up.

#### IP weighting

Interval-specific conditional probabilities for the two types of censoring mechanisms (treatment deviation or administrative reasons including loss to follow-up and death, where death is not part of the outcome definition) contributing to the IP weight denominator would be estimated using pooled logistic regression models, considering all time-constant and time-varying confounders (Supplemental Figure S1). As IP weights tend to be prone to extreme values, we would stabilize the weights using numerators that reflect treatment or censoring probabilities conditional on the time-constant covariates. These would also include the baseline values of the time-varying covariates. Both the underlying models for probabilities contributing to the denominator and the numerator of the weights would include linear and quadratic terms for follow-up time. We would calculate the interval-specific treatment and censoring weights separately by dividing the cumulative product of the respective numerator model’s predictions across all previous intervals by the cumulative product of the respective denominator model’s predictions across all previous intervals. The product of treatment and administrative censoring weights would be calculated as overall IP weights at each interval. The resulting stabilized weights would be truncated at the 99^th^ percentile.

#### Outcome models

Outcome models would be fit as pooled IP weighted logistic regression models and include terms for the assigned treatment strategy, linear and quadratic terms for follow-up time as well as interactions between treatment strategy and each of the two follow-up terms, respectively. Outcome models would additionally control for the time-constant confounders as covariates^13^.

### Target trial emulation in claims

#### Medicare Data

To emulate the hypothetical target trial described above, we used 2012 to 2020 U.S. Medicare claims of individuals with coverage for inpatient services (Medicare part A), outpatient services (part B), and outpatient prescription drug dispensing (part D). Medicare is a U.S. federal health insurance program for individuals aged 65 and above, and individuals below age 65 with a disability status ascertained by U.S. Social Security Administration. Because HFpEF is typically an age-related condition, Medicare covers the most suitable target population for our observational effectiveness study of spironolactone in HFpEF. Information on cause of death was available through linkage with the National Death Index until 12/2016 ^14^.

Individuals observed in the Medicare database were considered potentially qualifying for our target trial emulation if they had available information on age and gender, and at least one inpatient or outpatient heart failure diagnosis without previously recorded nursing home or hospice placement. If multiple HF diagnoses that could potentially qualify as index date were recorded across the observed patient history, we selected the earliest one where the patient was aged 65 years and older, had an enrolment window of at least 365 days of uninterrupted prior observation in the database and at least one previous hospitalization for heart failure (without evidence of a myocardial infarction) in the 180 days preceding the potential index date. We restricted the sample to HF cases probabilistically assigned to have HFpEF at the index date according to a previously validated phenotyping algorithm (positive predicted value 0.91; sensitivity 0.84 at probability cutoff 0.80) ^15^ and to individuals without records for exclusion criteria. Supplemental Figure S1 shows the study design diagram with temporal anchors for eligibility assessment, start of follow-up and covariate measurements, and the sequence in which eligibility criteria were applied. All International Classification of Diseases (ICD) version 9 and 10 diagnosis codes, procedural codes from ICD-9/ICD-10, Healthcare Common Procedure Coding System (HCPCS) and Current Procedural Terminology (CPT) as well as names of active ingredients for filled prescription medications that were used to operationalize eligibility criteria are available on OSF.

#### Treatment strategy emulation via the clone-censor-weight approach

We emulated a trial among patients who met eligibility criteria at one or more dates of a qualifying heart failure diagnosis, using the earliest qualifying diagnosis date as index. Based on observational healthcare data, we do not know the physician’s intention to initiate spironolactone treatment when they evaluate a patient and document a HF diagnosis during a medical visit; consequently, a patient’s observed data can be consistent with both the “initiation” and “non-initiation” strategies at index. To avoid immortal time bias, ^5,16^ we created 2 copies (clones) of each patient’s data, of which one was assigned to each possible strategy at index (i.e. one initiator and one non-initiator clone) and subsequently followed in this assigned strategy. We then censored each clone when their observed data became incompatible with their assigned strategy. After intercurrent events, clones were not censored even if their observed treatment trajectories deviated from their assigned treatment strategy. We subsequently followed each clone until the trial endpoint, death (where death was not part of the outcome definition), treatment switches without clinical indication, nursing home or hospice admission, interruption/discontinuation of Medicare enrolment, or December 31, 2020. For endpoints comprising cause-specific mortality information, December 31, 2016 represented an additional censoring reason, as information on cause of death was not available in the database after this date.

#### Follow-up

Follow-up time was discretized into 30-day intervals. Continuity of spironolactone treatment after initiation was defined based on days’ supply of the respective previous prescription fill plus a 30-day grace period. Treatment was considered continued in any subsequent monthly interval if at least one day of that interval was covered by either a previous prescription fill or by the 30-day grace period after the last fill.

#### Outcome ascertainment emulation

We used previously validated claims algorithms to emulate outcome ascertainment for heart failure^17^ and hyperkalemia^18,19^ hospitalizations and cardiac arrest^20^. For each outcome, we focused on the first observed event. Supplemental Table S1 lists all ICD-9 and ICD-10 codes that were used to operationalize outcomes.

Additional details on the data preparation process (treatment of missing variables, dataset structures) are provided in Supplemental Text S1. To ensure a logically sound chronological sequence of measurements that is compatible with causality, we incorporated one-month time lags between covariate, treatment and outcome measurements, respectively (Supplemental Figure S2). Individuals with outcome events in the first follow-up interval were excluded from the respective outcome analysis, resulting in slightly different analysis datasets for each outcome.

#### Causal estimand emulation

As we were interested in the comparative outcomes of two deterministic, sustained, static treatment strategies^21^, we used an as-treated analysis to estimate an observational analogue of the per-protocol effect. IP weighting was used to account for the fact that censoring for treatment deviations is likely informative. Additional details on the weighting process are provided in Supplemental Text S2 and Supplemental Figure S3.

#### Statistical analysis

We calculated descriptive statistics (mean and standard deviation of for continuous variables, and absolute and relative frequencies for categorical variables) for all covariates for the overall Medicare cohort, and for the restricted cohort that we used for benchmarking (see section on assessment of validity threats below). Where published, we also provided descriptive characteristics from the TOPCAT trial (Americas randomization stratum) for comparison. IP weights and outcome models were estimated analogously to the hypothetical target trial outlined above, with the only difference that outcome models additionally included a linear term for cumulative treatment history (number of previous intervals on spironolactone).

Absolute risks over the follow-up time were estimated from these outcome models stratified by treatment strategies and standardized to the distribution of baseline variables in the observed Medicare HFpEF population for each follow-up interval. These allowed us to derive estimates of absolute risk differences and relative risk for the 12-month follow-up time point. We obtained 95%-quantile based confidence intervals from bootstrapping individuals from the original cohort using 200 random samples with replacement, with each bootstrapped sample of the same size as the original cohort, and repeating the clone-censor-weight design and analysis in each of them.^13^

#### Subgroup analyses

Analyses were conducted in the overall cohort and within subpopulations characterized by age, gender, race, frailty status, polypharmacy status, and with concomitant diabetes, obesity, atrial fibrillation, pulmonary hypertension, coronary artery disease, or kidney disease, respectively.

#### Deterministic sensitivity analyses

We conducted three pre-specified deterministic sensitivity analyses in which we 1) reduced the time period between the index diagnosis and spironolactone initiation from 6 months to 3 months, 2) restricted the population to individuals with no recorded spironolactone prescriptions at any point in their observed treatment history instead of a 180-day washout, and 3) truncated IP weights at the 95^th^ instead of the 99^th^ percentile. Additionally, we conducted a post-hoc sensitivity analyses in which we 4) ran the outcome models with unstabilized truncated weights, and 5) repeated the analyses for the all-cause death outcome in the dataset restricted to periods where cause-specific death information was available.

### Assessment of threats to validity due to design-related biases and confounding in an observational non-use comparison

To assess the extent to which the target trial emulation via clone-censor-weighting alleviated known potential threats to validity when comparing an active treatment with non-use in observational data, we conducted a series of pre-specified diagnostics and post-hoc sensitivity analyses.

#### Measured confounders

To assess the potential for confounding by indication according to measured confounders at baseline, we 1) calculated standardized differences for time-fixed baseline covariates between spironolactone initiators and non-initiators before cloning. To assess the extent to which controlling for these measured confounders altered the results, we 2) presented effect estimates from unweighted outcome models with and without baseline covariate adjustment. To assess the success of balancing measured time-varying confounders, we 3) created plots of standardized differences in the unweighted and IP weighted pseudo-population with untruncated unstabilized and untruncated stabilized IP weights for all time-varying confounders at baseline, 6, and 11 months follow-up to allow for visual assessment of the impact of the time-varying weighting on covariate balance over time. The 6-month time point was chosen as it represents the end of the grace period where treatment weights start exerting a considerable influence on the pseudo-population of the cloned arm assigned to treatment non-initiation for the first time. The 11-month time point was chosen as it reflects the last treatment interval relevant for the 12-month outcomes analyses. To gauge the additional impact of IP weighting based on these measured time-varying confounders (the attempt to remediate potential treatment-confounder-feedback) on the results, we compared effect estimates from our main analysis to effect estimates of the unweighted outcome models with baseline covariate adjustment.

#### Immortal time bias

To illustrate the immortal time bias that we were able to avoid by using the clone-censor-weight approach, we repeated the analysis conducting a “naïve” ever- vs never-user comparison. We started follow-up for all individuals immediately after the index HF diagnosis. However, in contrast to the main analysis, we misclassified the subsequent immortal follow-up time for initiators between the index HF diagnosis and their observed spironolactone initiation as “exposed” (Supplemental Text S3).^22^.

#### Unmeasured confounders

To assess the net bias due to residual unmeasured confounding, we pre-specified two guardrails: 1) assessing the effect on non-cardiovascular mortality as a negative control outcome with a subsequent net bias correction, and 2) benchmarking against the TOPCAT trial results after restricting to a population meeting similar eligibility criteria.

##### Net bias assessment and net bias correction

To account for residual bias, we assessed the effects of spironolactone initiation and continued use on non-cardiovascular mortality as a negative control outcome. Non-cardiovascular mortality was considered a suitable negative control outcome as it should not be causally affected by spironolactone treatment, and it can be assumed to share important unmeasured confounders with cardiovascular mortality. With no residual confounding, the unbiased effect of an exposure on negative control outcomes should be null. Under a monotonicity assumption, bias adjusted results for the study endpoints can be derived by subtracting the treatment effect estimate on the negative control outcome (i.e. the net bias) from the treatment effect estimate on the primary outcome^23^. Results were presented with and without net bias adjustment based on the results for non-cardiovascular death in the overall population and in each subgroup to reflect the subgroup-specific confounding structure.

##### Benchmarking

The main analysis was accompanied by a benchmarking exercise, where we used available evidence on the size and direction of spironolactone effects on outcomes reported for the 2014 TOPCAT trial. Given a marked geographic variation observed in TOPCAT, we decided to specifically benchmark against selected TOPCAT results for the Americas. Where possible, we benchmarked against results from the Americas and restricted to the TOPCAT randomization stratum that was included on the basis of a prior hospitalization for heart failure. We prioritized these estimates because HF hospitalizations were an eligibility criterion that we could also replicate in claims, which was not possible for the other randomization stratum included based on Brain Natriuretic Peptide (BNP) measurements.

To construct our benchmarking cohort, a subsample of the overall cohort used in the main analysis, we followed TOPCAT eligibility criteria (see Supplemental Figure S1) and excluded individuals with records for Coronary Artery Bypass Graft (CABG) procedures, a hospitalization for stroke or myocardial infarction in the 90 days, or records for Percutaneous Coronary Intervention (PCI) in the 30 days before index. Furthermore, individuals were excluded if they had a diagnosis for cardiomyopathy, valve disorder, pulmonary disease combined with prescription fills for oral steroids, used home oxygen, were hospitalized for COPD exacerbations, diagnosed with orthostatic hypotension, severe chronic kidney disease (≥ stage 4), hepatic disease, gastrointestinal diseases thought to interact with drug absorption, alcohol or drug use, dementia or Alzheimer’s (to emulate risk of non-adherence) in the 365 days before index, or had a combined comorbidity index score^24,25^ above the 95% quantile or diagnosis for metastatic cancer (to emulate short expected life expectancy). We used the same clone-censor-weight design and IP weighting approach as for the main analysis (Supplemental Text S4).

For comparison of our observational benchmarking results to the published RCT results from TOPCAT, we assessed estimate agreement (i.e. if our 1-year RR estimate fell within the 95% CI for the TOPCAT HR estimate) and conducted a formal hypothesis test^26^ based on the standardized difference between the logarithm of effect estimates from our study and those from TOPCAT, which we then compared to the 95^th^ percentile of a standard normal distribution^27^.

All analyses were executed in SAS 9.4. Central programs are provided on OSF.

## Results

### Demonstrative use case: Real-world effectiveness of spironolactone in HFpEF

#### Study populations

A total of 320,881 individuals met all inclusion criteria in the overall cohort (Supplemental Table S2). Of these, 49,729 (15.5%) also fulfilled eligibility criteria for the benchmarking cohort. A total of 196,212 (61.1%) individuals overall and 31,603 (63.5%) in the benchmarking cohort had at least one time interval with information on cause-specific mortality (i.e. before 2017). Table 2 shows central descriptive baseline characteristics for both cohorts in comparison to the TOPCAT Americas subgroup^7,28^, or where the former were not available, the hospitalization stratum within this subgroup^29^. On average, both our overall and benchmarking cohorts were older (81 and 80 years, respectively) than TOPCAT participants (72 years); more often female (62.0 and 61.3%, respectively vs. 50%); and had a greater burden of comorbid conditions. Supplemental Table S3 shows additional baseline characteristics for our two study cohorts beyond those reported for TOPCAT Americas.

**Table 2.**
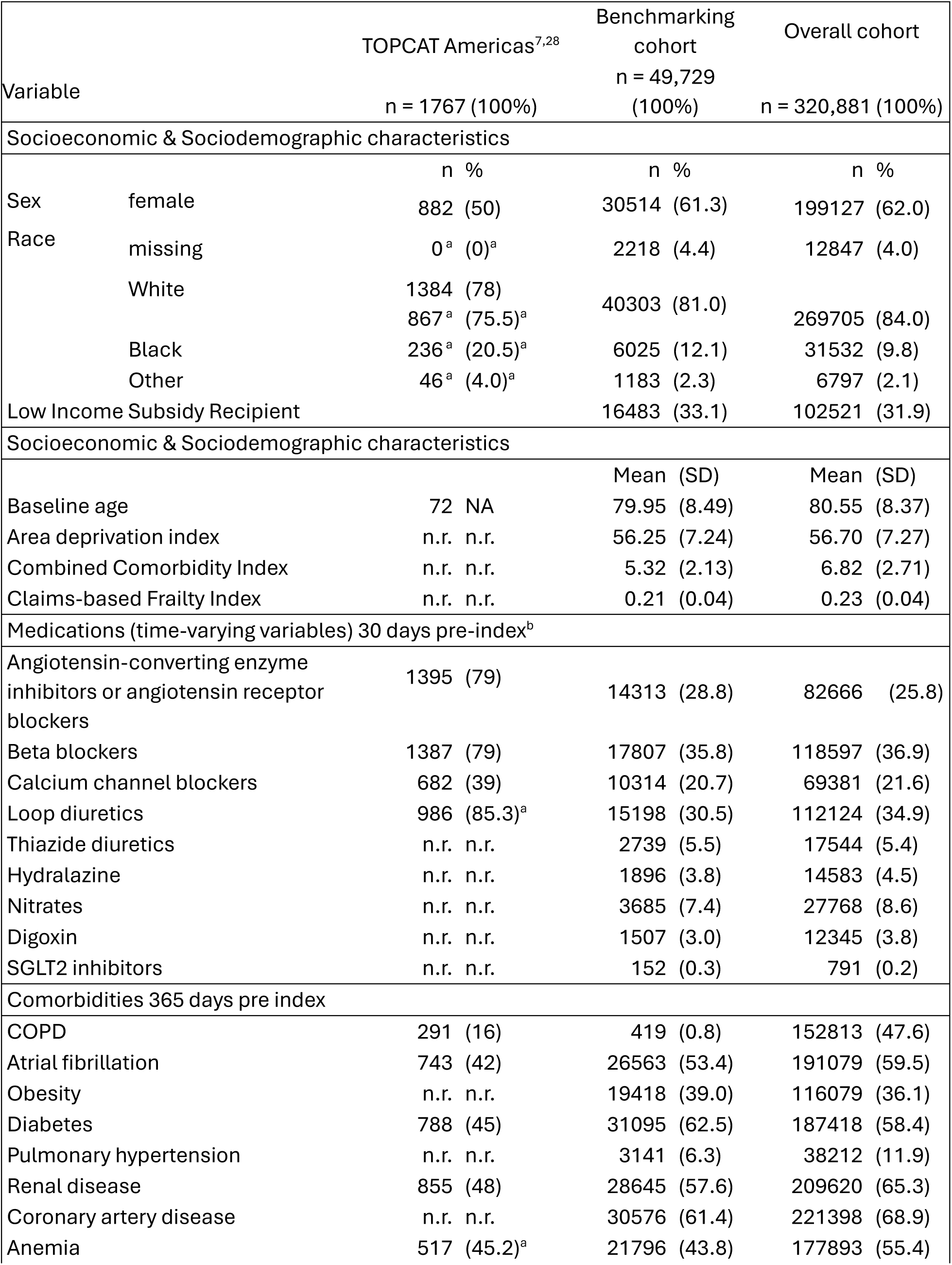
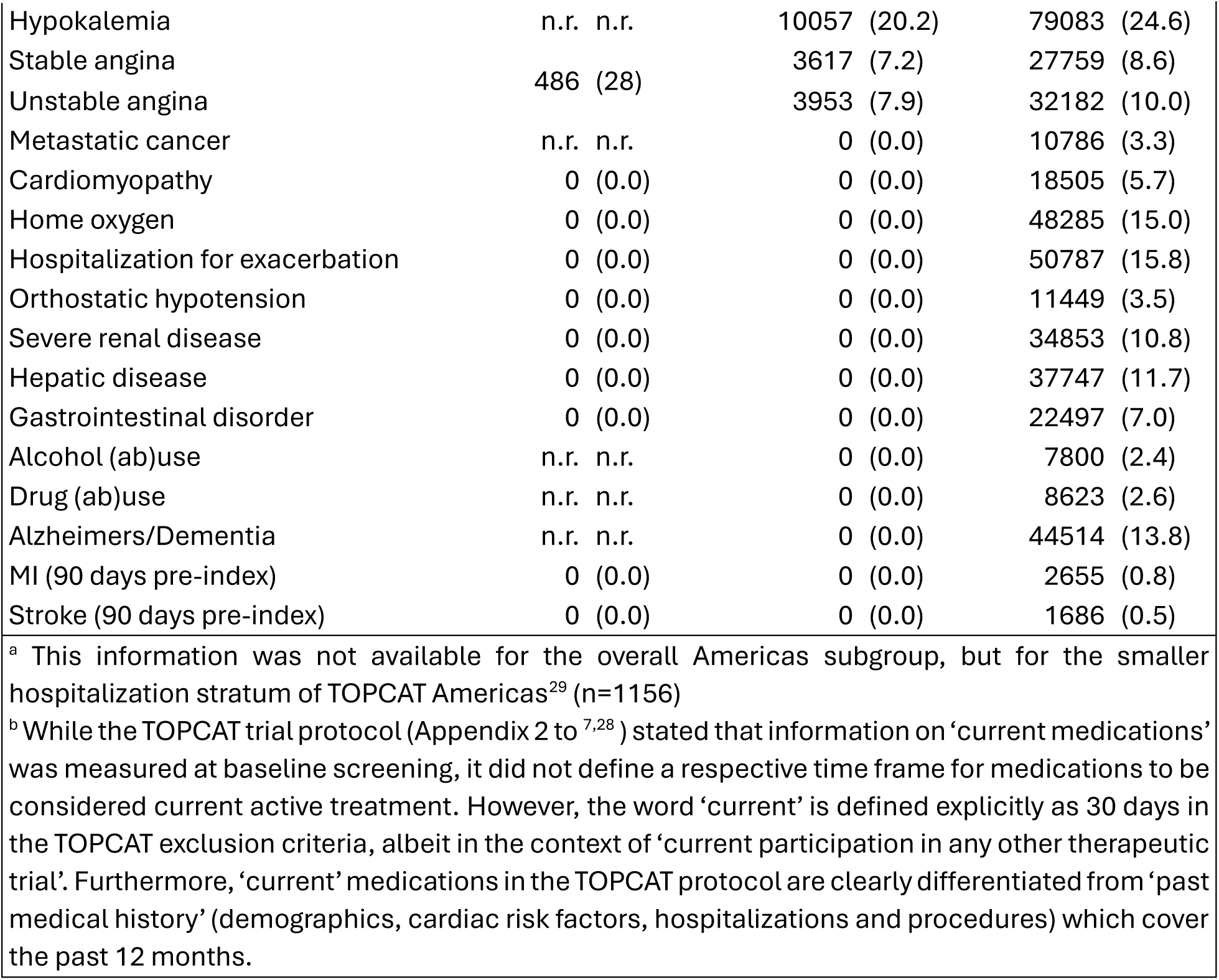
Descriptive characteristics for central variables.

#### Exposure patterns

Among the 320,881 individuals in the overall cohort, 29,762 (9.3%) filled a spironolactone prescription within 6 months of their index HFpEF diagnosis. Most prescription fills (64.3%) occurred in the first month after index, dropping to 11.9% and 8.1% in the second and third months, respectively (Supplemental Table S4). Descriptives for time on treatment can be found in Supplemental Figure S4 and Supplemental Table S5).

#### Pseudo-population created through cloning, censoring and weighting

A detailed description of analytic decisions and results from validity checks conducted for the cloning, censoring and weighting process can be found in Supplemental Text S5, Supplemental Tables S6-S15 and Supplemental Figures S5-S6.

### Outcome models

#### Results after cloning, censoring and weighting – overall cohort

Figure 1 shows parametric risk curves over 12 months for effectiveness outcomes and non-cardiovascular death as negative control outcome in the overall cohort. Absolute 12-month outcome risks ranged between 6.5% (6.9%) for non-cardiovascular death and 32.2% (32.6%) for the composite with all-cause death in the non-initiator (initiator) arm, respectively (Supplemental Table S16).

**Figure 1.**
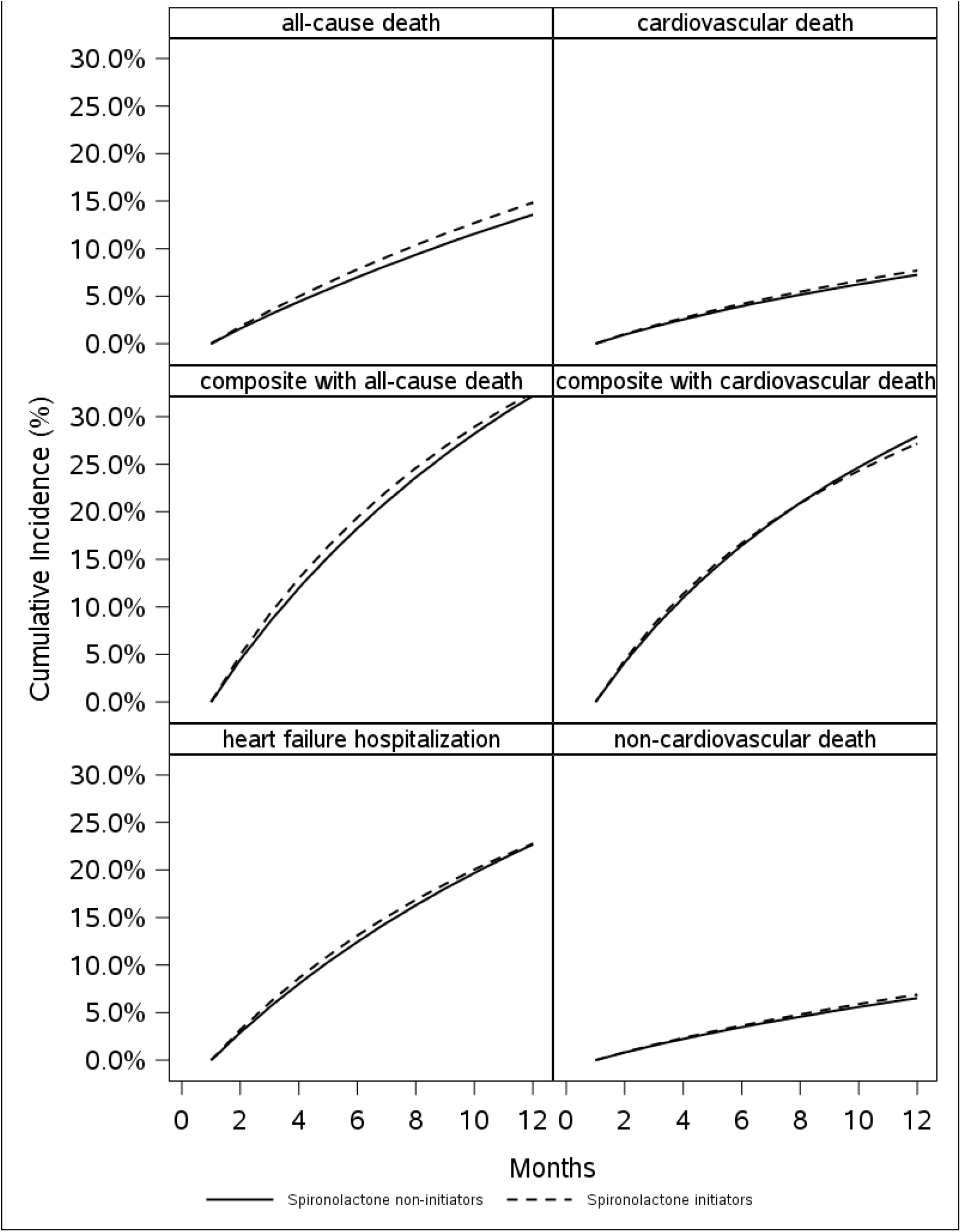
Cumulative incidence curves – overall cohort. Note: Parametric risk estimates derived from outcome models with covariate average values of the Medicare HFpEF population.

Without net bias adjustment, relative risks for effectiveness outcomes ranged between 0.97 (CI: 0.96; 0.98) for the composite with cardiovascular death and 1.09 (1.08; 1.10) for all-cause death (Figure 2), with the respective absolute risk differences ranging between -0.8% (CI: -1.1%; -0.5%) and 1.3% (CI: 1.1%; 1.4%) (Supplemental Table S17). Relative risk for hyperkalemia hospitalization was 1.22 (CI: 1.20; 1.24). Results were mostly consistent across subgroups (Supplemental Figures S7-S11) and deterministic sensitivity analyses (Supplemental Figure S12).

**Figure 2.**
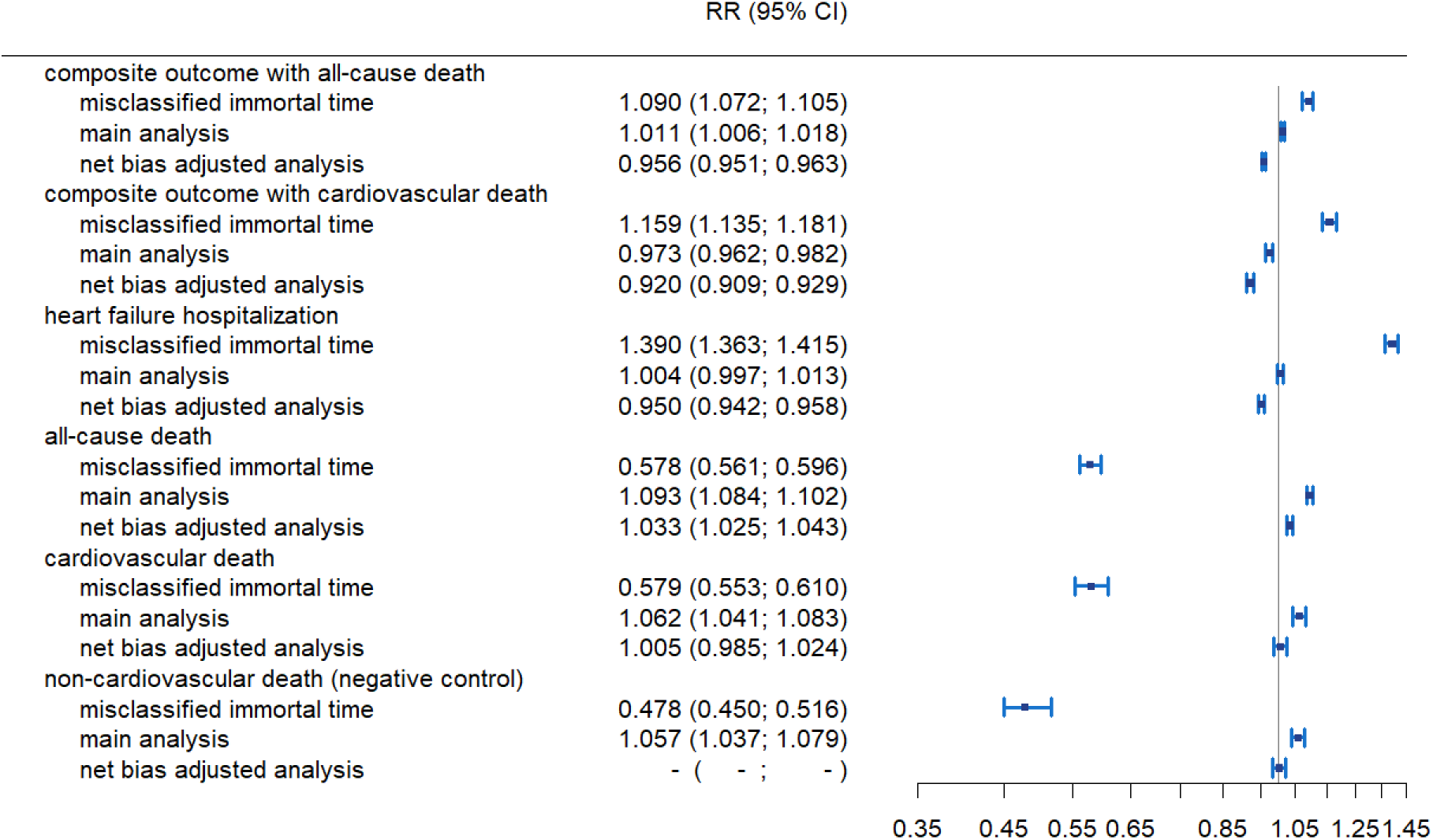
Forest plot illustrating changes in results with and without immortal time bias and net bias adjustment – overall cohort. Note: Confidence intervals are based on bootstrapping with 200 iterations for the main and net bias adjusted analysis, and 100 iterations for all other analyses, respectively.

### Assessment of threats to validity due to design-related biases and confounding in an observational non-use comparison

#### Measured confounders

A descriptive comparison of time-fixed covariates between initiators and non-users indicated that spironolactone initiators were more frequently obese and had recorded sleep apnea, but lower prevalence of kidney disease and dementia (Supplemental Table S18). Healthcare utilization showed more imbalances after baseline at month 6 and 11 without weighting, which were slightly reduced with stabilized weights (Supplemental Figures S13). Supplemental Figure S14 illustrates contributions of baseline adjustment and weighting on results in the overall cohort.

*Immortal time bias and unmeasured confounders*: Net bias assessment, net bias correction and benchmarking

The naïve analysis which misclassified immortal time by comparing ever-users against never-users resulted in higher relative risk estimates compared to the clone-censor-weight design for all outcomes comprising heart failure hospitalizations, and much lower relative risk estimates compared to the clone-censor-weight design when solely looking at mortality outcomes (Figures 2-3).

The effect estimate for the negative control outcome non-cardiovascular death (a measure of residual bias) in the main clone-censor-weight analysis was 1.06 (CI: 1.04; 1.08) in the overall and 1.10 (CI: 1.02; 1.19) in the benchmarking cohort. After correcting for the magnitude of estimated bias, relative risk estimates from the main clone-censor-weight analysis for all effectiveness outcomes except all-cause and cardiovascular death in both cohorts moved back to below 1, ranging between 0.92 (CI 0.91; 0.93) for the composite with cardiovascular death, and 1.03 (CI: 1.03; 1.04) for all-cause death in the *overall* cohort.

Both before (Supplemental Figure S15, Supplemental Table S19) and after net bias correction, estimate agreement between the benchmarking cohort and TOPCAT was reached only for the two outcomes that comprised heart failure hospitalization but not for the two single mortality outcomes for which published TOPCAT results were available (Figure 3, Supplemental Table S20).

**Figure 3.**
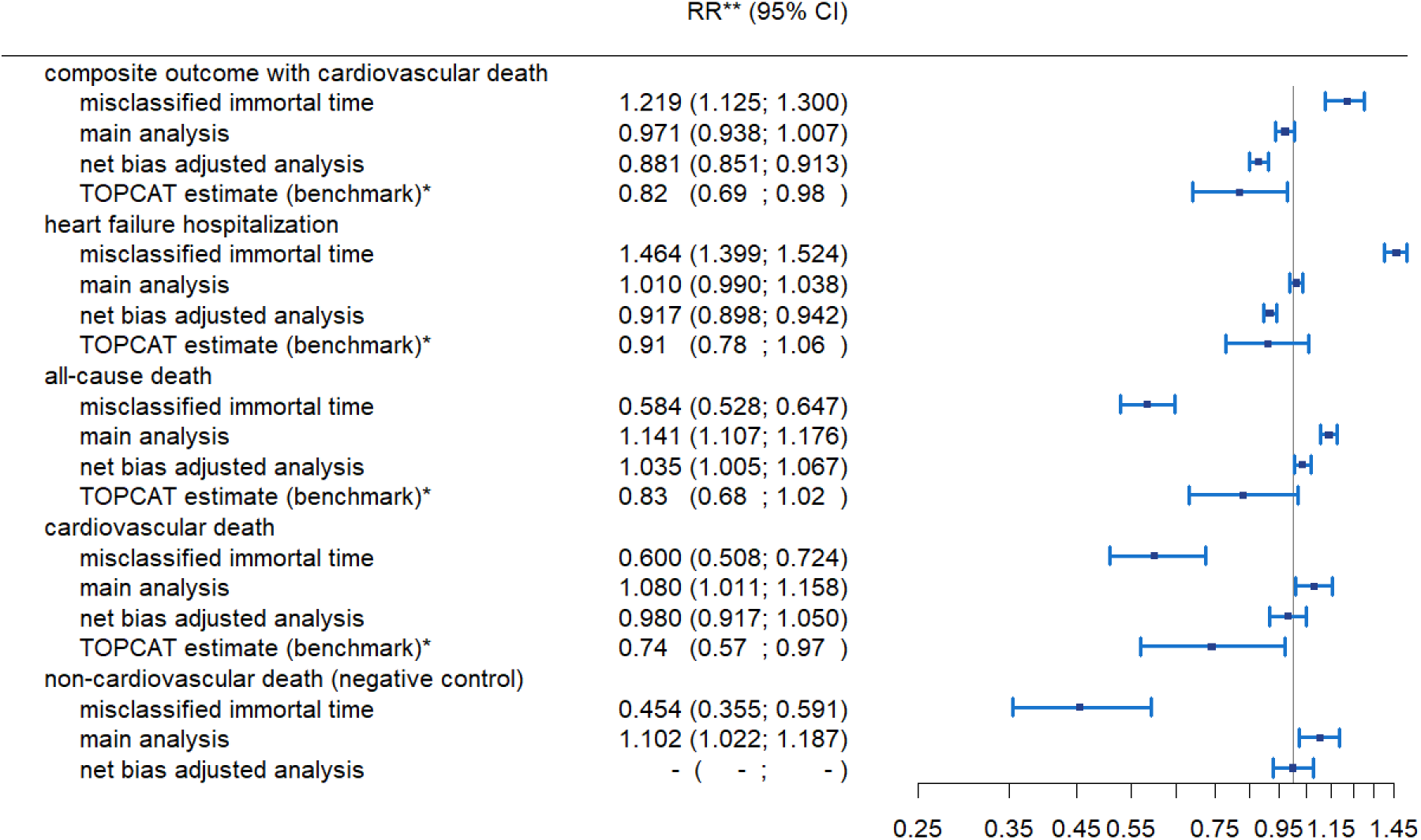
Forest plot illustrating changes in results with and without immortal time bias and net bias adjustment – benchmarking cohort. * estimates from the TOPCAT Americas subgroup, and in case of heart failure hospitalization from the hospitalization stratum. **Relative risks for the benchmarking cohort, hazard ratios for TOPCAT. Confidence intervals for the benchmarking cohort based on bootstrapping with 200 iterations for the main and net bias adjusted analysis, and 100 iterations for all other analyses, respectively.

## Discussion

In this study, we implemented a target trial emulation of a placebo-controlled trial for spironolactone in patients with HFpEF using the clone-censor-weight design. Based on pre-specified net bias analyses that comprised of benchmarking against an index trial (TOPCAT) and a negative control outcome (non-cardiovascular death), we observed potential residual bias in our emulated target trial. While the clone-censor-weight design avoided immortal time bias, our results also point to the importance of implementing additional guardrails to detect residual bias, especially in emulations of placebo-controlled trials.

As illustrated by our naïve analysis, observational emulations of active treatment versus non-use are prone to immortal time bias, which occurs when outcomes that are observed in individuals who survive long enough to initiate treatment are compared to outcomes in those who do not, and the difference is erroneously interpreted as a causal effect of treatment. In the spironolactone example, a naive comparison of ever-users against never-users attributed all deaths to the non-use group that occurred too early after index to allow spironolactone initiation, resulting in seemingly large protective effects of spironolactone. In contrast, for outcomes driven by the non-fatal hospitalization component, it resulted in seemingly adverse effects of spironolactone, likely due to reverse causation: heart failure hospitalizations that led to subsequent spironolactone initiation were erroneously attributed to this treatment strategy. Approaches such as a landmark analysis or a person-time analysis are frequently recommended to address immortal person time; however, these approaches do not explicitly specify the target trial and change the causal question being addressed^5,30^. A landmark analysis typically involves assigning subjects into treatment versus non-use groups based on treatment initiation within a pre-specified time period (i.e., the landmark) and starting follow-up after this time period. This approach can only estimate treatment effects among those who survive at least the landmark period, which is a causal question that does not correspond to a prospective RCT where follow-up starts immediately after treatment assignment. A person-time analysis avoids immortal time by allowing subjects to contribute person-time to the non-use group prior to treatment initiation and to the treatment group afterwards. However, this approach may be biased when risk of the outcomes varies over time and constant hazard ratios cannot be assumed during the follow-up time. The clone-censor-weight approach provides a principled way within the target trial framework to avoid immortal time bias in observational studies involving a non-use comparison while asking a clear causal question.

An important contribution of our investigation is the reminder it provides, that even when design-induced bias is avoided through the clone-censor-weight approach within the principled target trial framework, observational studies of non-use comparisons remain prone to other sources of bias that are well described in the pharmacoepidemiologic literature^31,32^. Specifically, unmeasured confounding by indication is a major concern when physicians may preferentially avoid treatment among patients who they perceive to be in a stable state based on factors that are incompletely recorded in routinely collected healthcare data. Symptoms and disease severity are two such factors for which we may not have been able to control despite our large pool of observed time-constant and time-varying confounders. It is likely that spironolactone initiators suffered more frequently from uncontrolled heart failure symptoms or a more severe disease stage, which introduces upward confounding away from the null as observed in our results. Another potential concern is differential surveillance because patients who are actively prescribed a treatment are more likely to also be closely monitored compared to patients who are not treated.

For research questions where a non-use comparison presents the most viable clinical alternative and prospective RCTs are not feasible due to ethical, logistical, or other reasons, emulation of target trials with observational data will continue to be a valuable alternative. However, as we demonstrate, it is especially important to build in pre-specified guardrails to detect residual bias whenever a target trial is emulated with non-use comparison. Benchmarking against a completed high-quality RCT (an index trial), mimicking the inclusion and exclusion criteria as closely as possible, is one approach for net bias evaluation. However, such trials may not always exist. Therefore, consideration of control outcomes that share similar confounding structures with the primary outcome, will likely be the most useful practical approach for net bias evaluation. Under a monotonicity assumption, subtracting the log-hazard ratio of the treatment effect on the negative control outcome from that for the primary outcome can allow investigators to indirectly correct for unmeasured confounding that is shared by the primary outcome and the negative control outcome^23^.

Alternative design choices and data sources may also be helpful in emulating target trials with placebo comparisons. In certain circumstances, investigators may be able to specify an alternative treatment, that is used for a similar indication but is known to have no effect on the outcome of interest (i.e. an active comparator), to emulate a placebo group and mitigate residual bias. For some research questions, mitigation of confounding bias may require turning to alternative data sources (if available) with more adequate measurement on disease severity and patient-specific factors such as socioeconomic status. However, even the use of more comprehensive observational data sources does not always protect against residual bias, as shown in a recent prospective target trial emulation on the effect of beta blockers in HFpEF after myocardial infarction^6,33^

Our study has some important limitations. First, we relied on administrative claims data from the Medicare program to emulate our target trial. While the population-based nature and large size of the sample are key strengths, misclassification of HFpEF is possible because it is not reliably coded in claims. However, we note that the phenotyping algorithm we used has been noted to have a very strong performance for identifying HFpEF in external samples ^34^ and has been used widely in defining HFpEF in prior high-quality observational studies^35,36^ conducted using Medicare data. Second, results for cause-specific outcomes were based on smaller sample sizes as Medicare linkage to NDI was not available after 2016. This reduced precision of estimates for outcomes with cause-specific death as a component. Finally, the index trial we considered for benchmarking (TOPCAT) has been subject to much uncertainty, with reports of irregularities in data from Russian and Georgian sites. Therefore, we restricted our benchmarking to the Americas subgroup. However, some limitations to emulating this trial remain: First, only results for the heart failure hospitalization outcome were available separately for each randomization stratum in the published TOPCAT Americas subgroup. For all other outcomes, published TOPCAT Americas results may be partially driven by the BNP randomization stratum which we could not emulate. Second, we observed differences in the distribution of characteristics between our benchmarking cohort and TOPCAT, which means that if treatment effects are modified by patient characteristics, the discrepancy between the emulation and the trial may be explained by heterogeneity and not solely by bias. Third, the median follow-up time in TOPCAT was 3.5 years, with approximately one third of participants in each group who deviated from their assigned treatment strategy over the 6-year study period contributing data to the intention-to-treat analysis. In contrast, due to the observed shorter median duration of spironolactone use in the Medicare population, we provide 12-month effect estimates only and focus on an as-treated analysis as treatment intentions are not recorded in claims. The benchmarking failure that we observed for the single mortality outcomes may therefore also be partially explained by the fact that benefits on cardiovascular death were also shown in TOPCAT to take longer to materialize than outcomes comprising heart failure hospitalization^28^. Lastly, it should be kept in mind that the net bias evaluation and correction rely on the untestable assumption of a consistent pattern of residual confounding for both the target effect estimate and the negative control outcome.

In conclusion, our study demonstrated practical challenges when emulating placebo-controlled target trials using observational data. These emulations have potential for residual bias even when known sources of bias such as immortal time can be avoided using principled approaches such as clone-censor-weighting. Therefore, a heighted scrutiny is needed when interpreting results from emulation of placebo-controlled target trials.

## Data Availability

Data use agreements and licensing agreements do not allow sharing of patient-level claims data with third parties. However, data can be requested at the vendor directly (Centers for Medicare & Medicaid Services, CMS).

## References

1. Stanley K. Design of randomized controlled trials. Circulation. 2007;115:1164–1169.

2. Franklin JM, Schneeweiss S. When and how can real world data analyses substitute for randomized controlled trials? Clinical Pharmacology & Therapeutics. 2017;102:924–933.

3. Acton EK, Willis AW, Hennessy S. Core concepts in pharmacoepidemiology: key biases arising in pharmacoepidemiologic studies. Pharmacoepidemiology and drug safety. 2023;32:9–18.

4. Suissa S. Immortal time bias in observational studies of drug effects. Pharmacoepidemiology and drug safety. 2007;16:241–249.

5. Hernán MA, Sterne JAC, Higgins JPT, Shrier I, Hernández-Díaz S. A Structural Description of Biases That Generate Immortal Time. Epidemiology. 2025;36:107–114. doi: 10.1097/ede.0000000000001808

6. Matthews AA, Dahebreh IJ, MacDonald CJ, Lindahl B, Hofmann R, Erlinge D, Yndigegn T, Berglund A, Jernberg T, Hernán MA. Prospective benchmarking of an observational analysis in the SWEDEHEART registry against the REDUCE-AMI randomized trial. European Journal of Epidemiology. 2024;39:349–361.

7. Pitt B, Pfeffer MA, Assmann SF, Boineau R, Anand IS, Claggett B, Clausell N, Desai AS, Diaz R, Fleg JL. Spironolactone for heart failure with preserved ejection fraction. New England Journal of Medicine. 2014;370:1383–1392.

8. Desai RJ, Wang SV, Sreedhara SK, Zabotka L, Khosrow-Khavar F, Nelson JC, Shi X, Toh S, Wyss R, Patorno E. Process guide for inferential studies using healthcare data from routine clinical practice to evaluate causal effects of drugs (PRINCIPLED): considerations from the FDA Sentinel Innovation Center. bmj. 2024;384.

9. Wang SV, Pottegård A, Crown W, Arlett P, Ashcroft DM, Benchimol EI, Berger ML, Crane G, Goettsch W, Hua W. HARmonized Protocol Template to Enhance Reproducibility of hypothesis evaluating real-world evidence studies on treatment effects: a good practices report of a joint ISPE/ISPOR task force. Value in Health. 2022;25:1663–1672.

10. Wang SV, Pinheiro S, Hua W, Arlett P, Uyama Y, Berlin JA, Bartels DB, Kahler KH, Bessette LG, Schneeweiss S. STaRT-RWE: structured template for planning and reporting on the implementation of real world evidence studies. Bmj. 2021;372.

11. Cashin AG, Hansford HJ, Hernán MA, Swanson SA, Lee H, Jones MD, Dahabreh IJ, Dickerman BA, Egger M, Garcia-Albeniz X. Transparent reporting of observational studies emulating a target trial: the TARGET Statement. bmj. 2025;390.

12. Wang SV, Schneeweiss S. Data checks before registering study protocols for health care database analyses. Jama. 2024;331:1445–1446.

13. Yoshida K, Liu J, Desai RJ, Glynn RJ, Solomon DH, Kim SC. Comparative safety of gout treatment strategies on cardiovascular outcomes using observational data: Clone-censor-weight target trial emulation approach. Epidemiology. 2023;34:544–553.

14. Desai RJ, Levin R, Lin KJ, Patorno E. Bias Implications of Outcome Misclassification in Observational Studies Evaluating Association Between Treatments and All-Cause or Cardiovascular Mortality Using Administrative Claims. Journal of the American Heart Association. 2020;9:e016906.

15. Desai RJ, Lin KJ, Patorno E, Barberio J, Lee M, Levin R, Evers T, Wang SV, Schneeweiss S. Development and preliminary validation of a Medicare claims–based model to predict left ventricular ejection fraction class in patients with heart failure. Circulation: Cardiovascular Quality and Outcomes. 2018;11:e004700.

16. Fu EL, Harhay M, Schneeweiss S, Desai R, Hernán MA. Starting right: aligning eligibility and treatment assignment at time zero when emulating a target trial. Available at SSRN 5177135. 2025.

17. McCormick N, Lacaille D, Bhole V, Avina-Zubieta JA. Validity of heart failure diagnoses in administrative databases: a systematic review and meta-analysis. PloS one. 2014;9:e104519.

18. Raebel MA, Smith ML, Saylor G, Wright LA, Cheetham C, Blanchette CM, Xu S. The positive predictive value of a hyperkalemia diagnosis in automated health care data. Pharmacoepidemiology and drug safety. 2010;19:1204–1208.

19. Fleet JL, Shariff SZ, Gandhi S, Weir MA, Jain AK, Garg AX. Validity of the International Classification of Diseases 10th revision code for hyperkalaemia in elderly patients at presentation to an emergency department and at hospital admission. BMJ open. 2012;2:e002011.

20. Shelton SK, Chukwulebe SB, Gaieski DF, Abella BS, Carr BG, Perman SM. Validation of an ICD code for accurately identifying emergency department patients who suffer an out-of-hospital cardiac arrest. Resuscitation. 2018;125:8–11.

21. Hernan MA, Robins JM. Causal Inference: What if. Boca Raton: Chapman & Hall/CRC; 2020.

22. Duchesneau ED, Jackson BE, Webster-Clark M, Lund JL, Reeder-Hayes KE, Nápoles AM, Strassle PD. The timing, the treatment, the question: comparison of epidemiologic approaches to minimize immortal time bias in real-world data using a surgical oncology example. Cancer Epidemiology, Biomarkers & Prevention. 2022;31:2079–2086.

23. Richardson DB, Laurier D, Schubauer-Berigan MK, Tchetgen ET, Cole SR. Assessment and indirect adjustment for confounding by smoking in cohort studies using relative hazards models. American journal of epidemiology. 2014;180:933–940.

24. Gagne JJ, Glynn RJ, Avorn J, Levin R, Schneeweiss S. A combined comorbidity score predicted mortality in elderly patients better than existing scores. Journal of clinical epidemiology. 2011;64:749–759.

25. Sun JW, Rogers JR, Her Q, Welch EC, Panozzo CA, Toh S, Gagne JJ. Adaptation and validation of the combined comorbidity score for ICD-10-CM. Medical care. 2017;55:1046–1051.

26. Franklin JM, Patorno E, Desai RJ, Glynn RJ, Martin D, Quinto K, Pawar A, Bessette LG, Lee H, Garry EM. Emulating randomized clinical trials with nonrandomized real-world evidence studies: first results from the RCT DUPLICATE initiative. Circulation. 2021;143:1002–1013.

27. Franklin JM, Pawar A, Martin D, Glynn RJ, Levenson M, Temple R, Schneeweiss S. Nonrandomized real-world evidence to support regulatory decision making: process for a randomized trial replication project. Clinical Pharmacology & Therapeutics. 2020;107:817–826.

28. Pfeffer MA, Claggett B, Assmann SF, Boineau R, Anand IS, Clausell N, Desai AS, Diaz R, Fleg JL, Gordeev I. Regional variation in patients and outcomes in the Treatment of Preserved Cardiac Function Heart Failure With an Aldosterone Antagonist (TOPCAT) trial. Circulation. 2015;131:34–42.

29. Szabo B, Benson L, Savarese G, Hage C, Fudim M, Devore A, Pitt B, Lund LH. Previous heart failure hospitalization, spironolactone and outcomes in heart failure with preserved ejection fraction–a secondary analysis of TOPCAT. American Heart Journal. 2024.

30. Jackson BE, Greenup RA, Strassle PD, Deal AM, Baggett CD, Lund JL, Reeder-Hayes KE. Understanding and identifying immortal-time bias in surgical health services research: an example using surgical resection of stage IV breast cancer. Surgical Oncology. 2021;37:101539.

31. Sendor R, Stürmer T. Core concepts in pharmacoepidemiology: confounding by indication and the role of active comparators. Pharmacoepidemiology and drug safety. 2022;31:261–269.

32. Her QL, Rouette J, Young JC, Webster-Clark M, Tazare J. Core Concepts in Pharmacoepidemiology: New-User Designs. Pharmacoepidemiology and drug safety. 2024;33:e70048.

33. Yndigegn T, Lindahl B, Mars K, Alfredsson J, Benatar J, Brandin L, Erlinge D, Hallen O, Held C, Hjalmarsson P. Beta-blockers after myocardial infarction and preserved ejection fraction. New England Journal of Medicine. 2024;390:1372–1381.

34. Sepehrvand N, Dover DC, Islam S, Kaul P, McAlister FA, Miller RJ, Fine NM, Howlett JG, Armstrong PW, Ezekowitz JA. Predicting heart failure with reduced or preserved ejection fraction from health records: external validation study. Heart Failure. 2023;11:1018–1020.

35. Fu EL, Patorno E, Everett BM, Vaduganathan M, Solomon SD, Levin R, Schneeweiss S, Desai RJ. Sodium–glucose cotransporter 2 inhibitors vs. sitagliptin in heart failure and type 2 diabetes: an observational cohort study. European heart journal. 2023;44:2216–2230.

36. Krüger N, Schneeweiss S, Fuse K, Matseyko S, Sreedhara SK, Hahn G, Schunkert H, Wang SV. Semaglutide and Tirzepatide in Patients With Heart Failure With Preserved Ejection Fraction. JAMA. 2025.

