## Supplemental Material for "Emulation of placebo-controlled index trials using observational data with cloning, censoring and weighting: Empirical assessment of constraints and credibility"

Supplemental Figure S8: Forest plot for results on the composite outcome with all-cause death at 12 months by subpopulation. Weighted outcome analyses using stabilized inverse probability of treatment and censoring weights truncated at the 99th percentile, with and without net bias adjustment based on the subgroup specific results for non-cardiovascular death – overall cohort.. 21

Supplemental Figure S9: Forest plot for results on all-cause death at 12 months by subpopulation. Weighted outcome analyses using stabilized inverse probability of treatment and censoring weights truncated at the 99th percentile, with and without net bias adjustment based on the subgroup specific results for non-cardiovascular death – overall cohort............................................................. 22

Supplemental Figure S10: Forest plot for results on the composite outcome with cardiovascular death at 12 months by subpopulation. Weighted outcome analyses using stabilized inverse probability of treatment and censoring weights truncated at the 99th percentile, with and without net bias adjustment based on the subgroup specific results for non-cardiovascular death – overall cohort .......................................................................................................................................... 23

**Supplemental Text S1: Additional details on the data preparation process.**

*Treatment of missing covariate values*

Missing covariate values were modelled as a separate variable level, unless missingness on one data field introduced perfect correlation between several analysis variables. This applied to 3 Zip-code based variables, where, in case of missing ZIP Codes, we imputed the respective state's mean area deprivation score (if state was not missing) or the overall mean value in case of unknown state. If state was unknown, the individual was assigned to a missing value category for region, whereas for categorization into urban or rural residence, we used a list of urban ZIP codes from census. Only if a patient's ZIP code was in the list, then residence was coded as urban, all other values including those of individuals with missing ZIP code information, residence was coded as rural.

*Data structure*

*General structure*

The structure of the master dataset incorporating time lags between covariate, treatment and outcome measurements is shown in Supplemental Figure S2. Only complete intervals with regard to treatment were kept in the dataset. As a consequence, individuals with only one (incompletely observed) treatment interval after index were excluded.

For individuals who initiated spironolactone at some point during the observed follow-up and discontinued or interrupted their prescription refills at some timepoint thereafter, we excluded all observed time intervals after the first spironolactone interruption, unless it was preceded or concurrent to an intercurrent event, as these time intervals were incompatible with both treatment strategies of interest. Furthermore, for individuals with a first spironolactone prescription fill after the end of the grace period for initiation, we excluded all observed intervals after that first prescription fill unless it was preceded or concurrent to an intercurrent event.

*Outcome-specific datasets*

For each outcome of interest, a separate dataset was derived from the master dataset. Individuals who had the respective outcome of interest documented in the first follow-up interval were excluded (even though, for outcomes other than death, they could have at least one additional, potentially incompletely observed interval thereafter), resulting in slightly different analysis samples at baseline for each outcome. Where individuals experienced outcomes during their last (incompletely observed) interval, these outcomes were attributed to the previous (fully observed) treatment interval due to the lagged time structure and thereby kept in the data sets for outcome analysis. We excluded all treatment intervals observed after an individual's first lagged observed outcome of interest (i.e. all treatment intervals chronologically concurrent to or following the respective outcome event).

Where individuals did not experience outcomes during their last (incompletely observed) interval, they were classified as censored after that interval. As for outcomes, due to the lagged time structure, the censoring was attributed to the last (fully observed) treatment interval.

*Datasets used to model the conditional probability of treatment*

To model the conditional probability of treatment initiation and treatment strategy (dis-)continuation in each follow-up interval, we used the outcome-specific datasets described above, up to and including intervals of first spironolactone interruption and of first initiation after the end of the 6-month grace period, but excluded all observed treatment intervals concurrent with or following an intercurrent event.

*Datasets used to model the conditional probability of remaining uncensored*

For modelling the conditional probability of remaining uncensored in each follow-up interval, we used the outcome-specific datasets described above, excluding intervals of first spironolactone interruption at any time during follow-up and of first initiation after the end of the 6-month grace period, but including all observed treatment intervals concurrent with or following an intercurrent event.

*Datasets used for outcome models*

For modelling the conditional probability of experiencing the respective outcome of interest, we appended two cloned and censored versions of the outcome-specific datasets described above, one censored for spironolactone treatment strategy deviations and one censored for no-spironolactone treatment strategy deviations. The former dataset thus excluded intervals of and after first spironolactone interruption and intervals of and after first spironolactone initiation, if this initiation occurred after the end of the 6-month grace period. The latter excluded intervals of and after first initiation at any time during follow-up. For both cloned datasets, these exclusions were not applied if the treatment strategy deviation occurred concurrent with or after an intercurrent event.

*Further implications of the 6-month grace period for cloning and censoring*

Clones of individuals with a spironolactone prescription fill in the first month after index were immediately censored from the non-initiation strategy (equivalent to not being cloned in the first place) as their treatment strategy was distinguishable at start of follow-up, leading to slightly smaller sample sizes for the non-initiator treatment strategy as compared to the initiator strategy (always full sample size) after cloning and censoring before weighting at baseline.

All clones who did not initiate treatment during the 6-month grace period were censored from the initiators treatment strategy after month 5.

Supplemental Text S2: Additional details on the weighting process.

### Treatment and censoring models

We used indicators for discrete variables, linear terms for all (quasi-)continuous variables, and no product terms except for interactions of linear and quadratic follow-up interval with treatment. Treatment models estimated the probability of being on treatment conditional on not yet having experienced the respective outcome or an intercurrent event, not having interrupted a previously initiated spironolactone treatment before, and on the time-varying covariate values measured in the 30 days preceding the respective treatment interval. Administrative censoring models estimated the probability of remaining uncensored (i.e. having treatment and subsequent outcome information) in the subsequent interval, conditional on not yet having experienced the respective outcome event and on not having interrupted a previously initiated spironolactone treatment. Censoring status at time t+1 was attributed to the previous (fully observed, including the lagged outcome) treatment interval t and regressed on the covariates, and (as additional time-varying predictors in the models informing weight denominators) intercurrent events preceding the treatment interval (i.e. measured at time t-1) in the respective IP weight models.

All models contributing to the stabilized IP weights were estimated in the original dataset before cloning, and stratified by treatment status in the preceding interval, as we assumed that probability for initiating spironolactone would substantially differ from the probability of continuing spironolactone once initiated, and to allow for the possibility that the confounding effect of covariates might vary between initiation and (dis-)continuation of spironolactone. The same assumptions were made for censoring probabilities.

Each individual's interval-specific estimated conditional probabilities for treatment initiation or treatment continuation as well as estimated conditional probabilities for remaining uncensored were merged back into the cloned and censored dataset for outcome analysis to calculate the cumulative weights.

### Weight calculation

For all clones assigned to the treatment arm, weight contributions were set to 1 for the first analysis interval (no possibility of treatment deviation by definition), and, for those who had not previously initiated treatment, during the subsequent intervals up to the 5$^{th}$ grace period interval. From the interval after treatment initiation onwards, weight contributions estimated from the models for treatment continuation were used. For clones assigned to the non-initiator arm and without a previous or concurrent intercurrent event, complement probabilities to those estimated from the models for treatment initiation were used in each interval (Supplemental Figure S3).

For clones assigned to the treatment arm who initiated in the 6$^{th}$ interval of the grace period, weight contributions estimated from the models for treatment initiation in this interval were used to upweight these clones in this interval. Conceptually, this weighting is equivalent to forced initiation in interval 6 of all censored clones that did not initiate up to this point in timel[1]. As a consequence of the cumulative character of the interval-specific weight contributions over time, upweighted individuals in interval 6 remained influential throughout the remainder of their observed follow-up.

For treatment weight calculations, in both clone arms, interval-specific conditional predicted probabilities of individuals with previous intercurrent events were set to 1 from the interval of the intercurrent event onwards.

For "administrative" censoring weights, in both clone arms, interval-specific conditional predicted probabilities of remaining uncensored were used independently of previous intercurrent events, grace period interval and treatment initiation status. Predicted probabilities of remaining uncensored were used lagged by one interval, as they were derived from models predicting the probability of being censored in the subsequent interval.

We calculated the interval-specific treatment and censoring weights separately by dividing the cumulative product of the respective numerator model's predictions across all previous intervals by the cumulative products of the respective denominator model's prediction across all previous intervals.

### Supplemental Text S3: Additional details on the weighting process for the naïve approach

*Datasets used to model the conditional probability of treatment for the naïve analysis*

To model the conditional probability of treatment initiation and treatment strategy (dis-)continuation in each follow-up interval, we used the outcome-specific datasets described in Supplemental Text S1 and included data from the index HF diagnosis up to and including intervals of first spironolactone interruption independent of timing of first initiation, but excluded all observed treatment intervals concurrent with or following an intercurrent event.

*Datasets used to model the conditional probability of remaining uncensored for the naïve analysis*

For modelling the conditional probability of remaining uncensored in each follow-up interval, we used the outcome-specific datasets described in Supplemental Text S1, excluding intervals of first spironolactone interruption at any time during follow-up, but including all observed treatment intervals concurrent with or following an intercurrent event.

*Treatment and censoring models for the naïve analysis*

For the naïve approach, analogously to our approach within the main clone-censor-weight analysis, we calculated separate treatment initiation and continuation models using all observations before their first respective outcome or intercurrent event. The treatment initiation model was restricted to the first observed interval after index HFpEF diagnosis and estimated the baseline probability at index of ever being assigned to spironolactone use. In addition, the treatment continuation model estimated the probability of remaining on spironolactone treatment once actually initiated.

Censoring models were set up the same way as for the main clone-censor-weight analysis, but recalculated in the outcome-specific data sets for the naïve analysis (i.e. in datasets where no the 6-months grace period for spironolactone initiation was defined, and therefore intervals following later spironolactone initiation times were – in contrast to the main clone-censor-weight approach - not removed).

*Weight calculation for the naïve analysis*

At baseline, treatment weight contributions were the baseline interval-specific conditional predicted probability of being a spironolactone ever user for ever users, and the complement probabilities for never users. From the subsequent interval onwards until the actual interval of initiation, ever users received a weight contribution of 1. Once actually initiated, weight contributions estimated from the models for treatment continuation were used. For never users no further treatment weights were applied after the initial weighting at baseline.

For treatment weight calculations, in both never and ever users, interval-specific conditional predicted probabilities of individuals with previous intercurrent events were set to 1 from the interval of the intercurrent event onwards.

“Administrative” censoring weights were calculated analogously to the main clone-censor-weight analysis.

### Supplemental Text S4: Additional details on the weighting process for the benchmarking exercise

#### Treatment and censoring models

Treatment and censoring models for the benchmarking exercise were identical to those for the main analysis, with the exception that the list of time-constant covariates used to stabilize the weight calculation (and for adjustment in the outcome model) was reduced by 15 covariates which served as additional exclusion criteria for cohort selection and therefore showed no variation in the benchmarking cohort by design.

**Supplemental Text S5: Results from validity checks for creation of the pseudo-population through cloning, censoring and weighting**

*Cloning & Censoring*

Supplemental Table S5 shows how we arrived from the overall cohort with 320,881 individuals to the baseline sample sizes for each outcome-specific dataset and clone arm. Depending on the outcome of interest, between 143,455 (composite outcome with cardiovascular death) and 254,605 (all-cause death) individuals had more than one observed interval and no recorded outcome in the first interval, such that we could include them in the analysis. Amongst these, between 9,306 (composite outcome with cardiovascular death) and 17,499 individuals (hyperkalemia hospitalizations) had treatment strategies distinguishable at baseline (i.e. a spironolactone prescription in the first interval) and were therefore immediately censored from the non-initiator arm. Supplemental Table S6 shows the frequency of reasons for end of follow-up (censoring due to treatment deviation, outcome, administrative censoring including competing risk) for each outcome analysis stratified by clone arm, as well as the number of individuals with intercurrent events amongst those with outcomes and administrative censoring, respectively. While most clones assigned to the spironolactone strategy were censored for treatment deviation (at the end of the grace period), most clones assigned to the non-initiator strategy were administratively censored (at the end of their observation period). Outcomes were observed for between 7.0% (non-cardiovascular death) and 32.3% (composite outcome with all-cause death) of all clones. Supplemental Figure S5 illustrates the contributions of different censoring mechanisms to the reduction in sample size over time, stratified by treatment strategy, using the composite of HF hospitalizations, cardiac arrest and all-cause death as an example.

*IP Weighting*

Due to low or no observed frequencies that resulted in model convergence issues, we removed one time-varying and removed or recategorized 19 time-constant variables from all treatment, censoring and outcome models, resulting in utilization of 19 of the 20 time-varying covariates for weighting and 124 of the 143 time-constant covariates for stabilization and adjustment. Six additional time-constant and two time-varying variables we removed from all models relating to cause-specific death outcomes, which encountered more convergence issues due to the smaller sample sizes. Supplemental Tables S7-S8 show the distribution of untruncated unstabilized, as well as untruncated and truncated stabilized weights for the entire follow-up and the target 12-month follow-up period, respectively. Both stabilization and subsequent truncation substantially reduced the variability of weights. Mean stabilized weights for the 12-month period ranged between 0.999 (hyperkalemia hospitalizations) and

1.006 (heart failure hospitalizations). Supplemental Figure S6 shows the distribution of weights over time for the overall cloned and censored sample, as well as stratified by clone arm, the presence of intercurrent events and the treatment status at the end of grace period. As expected weights were larger towards the end of follow-up, in the initiator clone arm, and for end-of grace period initiators without previous intercurrent events.

Supplemental Tables S9-S12 illustrate the sample sizes of the pseudo-population based on unstabilized untruncated and stabilized untruncated weights for each treatment strategy in the first twelve months of follow-up using the heart failure hospitalization outcome as an example. Correctly specified weight models should result in a pseudo-population size derived from *unstabilized* weights that is (approximately) equal to the original study population *before* censoring[2]. While the predominant censoring mechanism was administrative reasons (in both clone arms until the end of the grace period, and in the unexposed clone arm also thereafter), the pseudo population outnumbered the *observed population at start of follow-up*. Conversely, in the exposed clone arm from the end of the grace period onwards the predominant censoring mechanism was treatment deviation, and in the respective intervals the pseudo population remained below the initial observed population size. Supplemental Tables S9-S10 therefore suggest some degree of residual misspecification of the treatment (lower than ideal weights) and censoring (higher than ideal weights) models.

Correctly specified weight models for *stabilized* weights should result in a pseudo population size (approximately) equal to the original study population *after* censoring[2]. In both clone arms until the end of the grace period, and in the exposed clone arm also thereafter, the pseudo population outnumbered the *observed population in the respective interval*. Conversely, in the unexposed clone arm from the end of the grace period onwards the pseudo population remained below the respective observed population size. Supplemental Tables S11-S12 therefore suggest some degree of residual misspecification of the treatment and censoring stabilized weights, potentially indicating that stabilization lead to higher than ideal treatment weights and lower than ideal censoring weights.

The number of actual initiators in the last month of the grace period who were upweighted from the sixth follow-up interval onwards and therefore largely influenced the composition of the pseudo-population thereafter ranged from n=398 (composite endpoint with cardiovascular death) to n=1156 (all-cause death, hyperkalemia hospitalizations) (Supplemental Table S13).

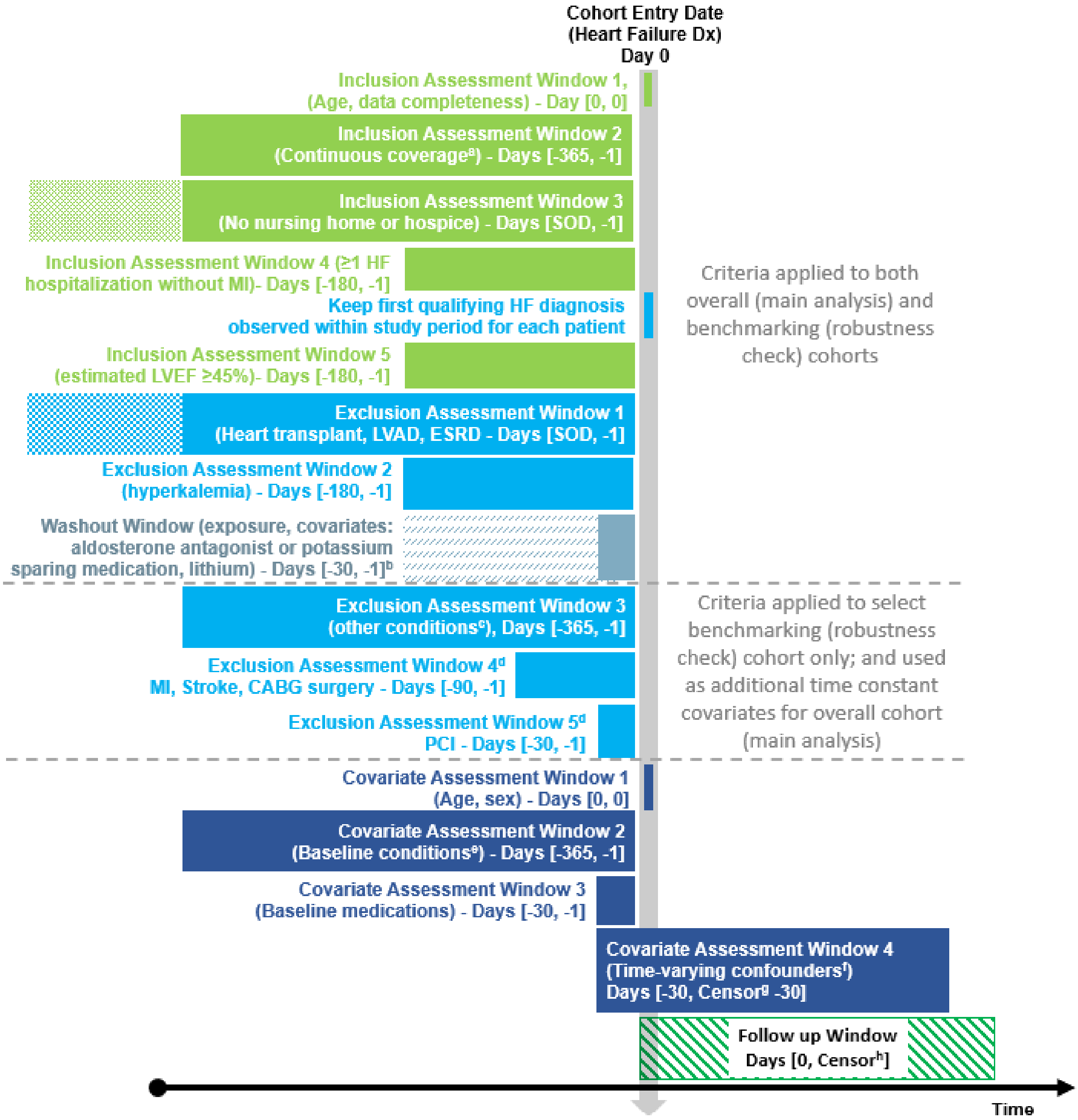

**Supplemental Figure S1. Study Design Diagram.**

Footnotes:

a. No gaps in medical or pharmacy enrollment allowed in the 365 days before index date ($T_0$)

b. Shaded grey area reflects the 6-month time period to be checked for potential previous spironolactone prescription fillings that might still be in use in the washout window, leading to potential spironolactone use in the 30 days pre-index

c. Other conditions will only serve as exclusion criteria for the benchmarking cohort subset and include LE<3 years (operationalized through metastatic cancer and ≥95 quantile of the combined comorbidity index), home O2, chronic pulmonary disease requiring oral steroid therapy, hospitalization for exacerbation, cardiomyopathy, valve disorder, orthostatic hypotension, gastrointestinal disorder, drug abuse, alcohol abuse, dementia or Alzheimer’s, severe renal dysfunction, hepatic disease

d. Exclusion assessment for benchmarking cohort only. For the overall cohort, this window serves as covariate assessment window for the respective time constant baseline condition variable (see footnote e)

e. **Baseline covariates (time constant) for both cohorts include**: Gender (male, female), Age, Race (black, white, other, unknown), Calendar year of index (2013, 2014, 2015, 2016, 2017, 2018, 2019, 2020), Index diagnosis recorded during an outpatient visit, Cardiac arrest, Kidney Disease, Frailty score, Diabetes, Obesity, Atrial fibrillation, Pulmonary hypertension, Coronary artery disease, Area deprivation of region of residence (socioeconomic status index), Low-income subsidy recipient status, Region of residence (Northeast, Midwest, South, West, unknown), Rural/urban place of residence, Statins*,**, Insulin preparations*,**, Hypoglycemic agents*,**, Antiplatelets*,**, Anticoagulants*,**, Gout medications*,**, COPD medications*,** (level 1, 2, 3, 4, 5, 6), Oral steroids*,**, NSAIDs*,**, Pressor amines*,**, Barbiturates*,**, Narcotics*,**, Cardiac glycosides other than Digoxin, Skeletal muscle relaxants*,**, Calcium channel blockers**, predicted probability of HFpEF according to the classification algorithm, Combined Comorbidity Score, Angioedema, Anemia, Depression, Diabetic nephropathy, Endocarditis, Hypertensive nephropathy, Hyperlipidemia, Hypertension, Hypokalemia, Hypotension, Other dysrhythmias, Peripheral arterial disease, Psychosis, Rheumatic heart disease, Sleep apnea, Stable angina, Thyroid dysfunction, Unstable angina, Hemorrhage, Cardiac Re-synchronization Therapy, Implantable cardioverter defibrillator, valve replacement, valve correction, valvular heart disease / valve disorder, COPD, chronic pulmonary disease other than COPD, No. of outpatient office visits for any reason, No. of physicians associated with prescription claims for patient, No. of hospitalizations for any reason, No. of emergency room visits for any reason, No. of cardiologist visits for any reason, Bone Mineral Density (BMD) testing, Colonoscopy, Influenza vaccination, Pneumococcal vaccine, Herpes zoster vaccination, Mammography, Pap or HPV test, Fecal occult blood test, prostate specific antigen test, Myocardial infarction (MI)****, Stroke***, Coronary Artery Bypass Graft (CABG) surgery****, Percutaneous Coronary Intervention (PCI)**, Smoking.

**Additional baseline conditions (time constant) for the overall cohort include**: Home O2, Hospitalization for exacerbation, Metastatic cancer, Orthostatic hypotension, Gastrointestinal disorder, Non-ESRD severe chronic kidney disease, Hepatic disease, Alcohol abuse, Drug abuse, Dementia & Alzheimer's disease, Myocardial infarction (MI)***, Stroke***, Coronary Artery Bypass Graft (CABG) surgery***, Percutaneous Coronary Intervention (PCI)*, Cardiomyopathy.

*in days 30-1 before index
**in days 365-31 before index
***in days 90-1 before index
****in days 365-91 before index

f. **Time varying covariates for both cohorts include**: No. of hospitalizations for heart failure*, No. of distinct medications*, ACE (Angiotensin-converting enzyme) inhibitors*, ARBs (Angiotensin receptor blockers)*, Beta blockers*, Calcium channel blockers*, Loop diuretics*, Thiazide diuretic*, Hydralazine*, Nitrate combinations*, Digoxin (Lanoxin)*, angiotensin-neprilysin inhibitors Sacubitril/valsartan (Entresto)*, Ivabradine (Corlanor)*, SGLT2*, No. of outpatient office visits for any reason*, No. of physicians associated with any claims for patient*, No. of physicians associated with medication claims for patient*,

No. of hospitalizations for any reason*, No. of emergency room visits for any reason*, No. of cardiologist visits for any reason*.

*baseline values of variable additionally used as time-constant covariate.

g. Time-varying confounders for each month of follow-up will be assessed as of the previous time interval preceding treatment. The length of assessment windows for time-varying confounders will consist of 30-day intervals.

h. Earliest of: outcomes of interest, switching or discontinuation of study drug (unless preceded by hyperkalemia, a heart transplant, lithium use initiation, LVAD, or ESRD), nursing home or hospice placement, disenrollment, death, end of the study period (12/31/2020)

HF = heart failure, LE = life expectancy, LVEF = left ventricular ejection fraction, MI = Myocardial infarction, GABG = Coronary artery bypass graft, PCI = Percutaneous Coronary Intervention, LVAD = left ventricular assist device

Grey /blue font: Variable removed from all or from cause-specific death models to avoid convergence issues.

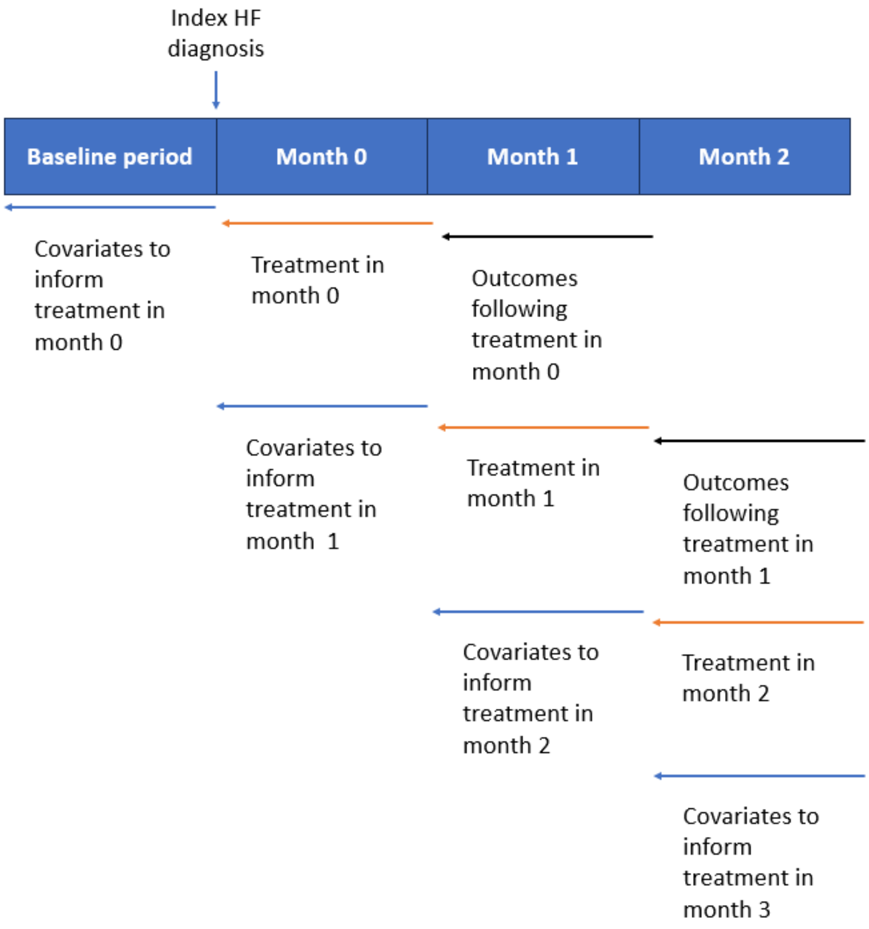

Supplemental Figure S2: Sequence of covariate, treatment and outcome measurements in the analysis dataset

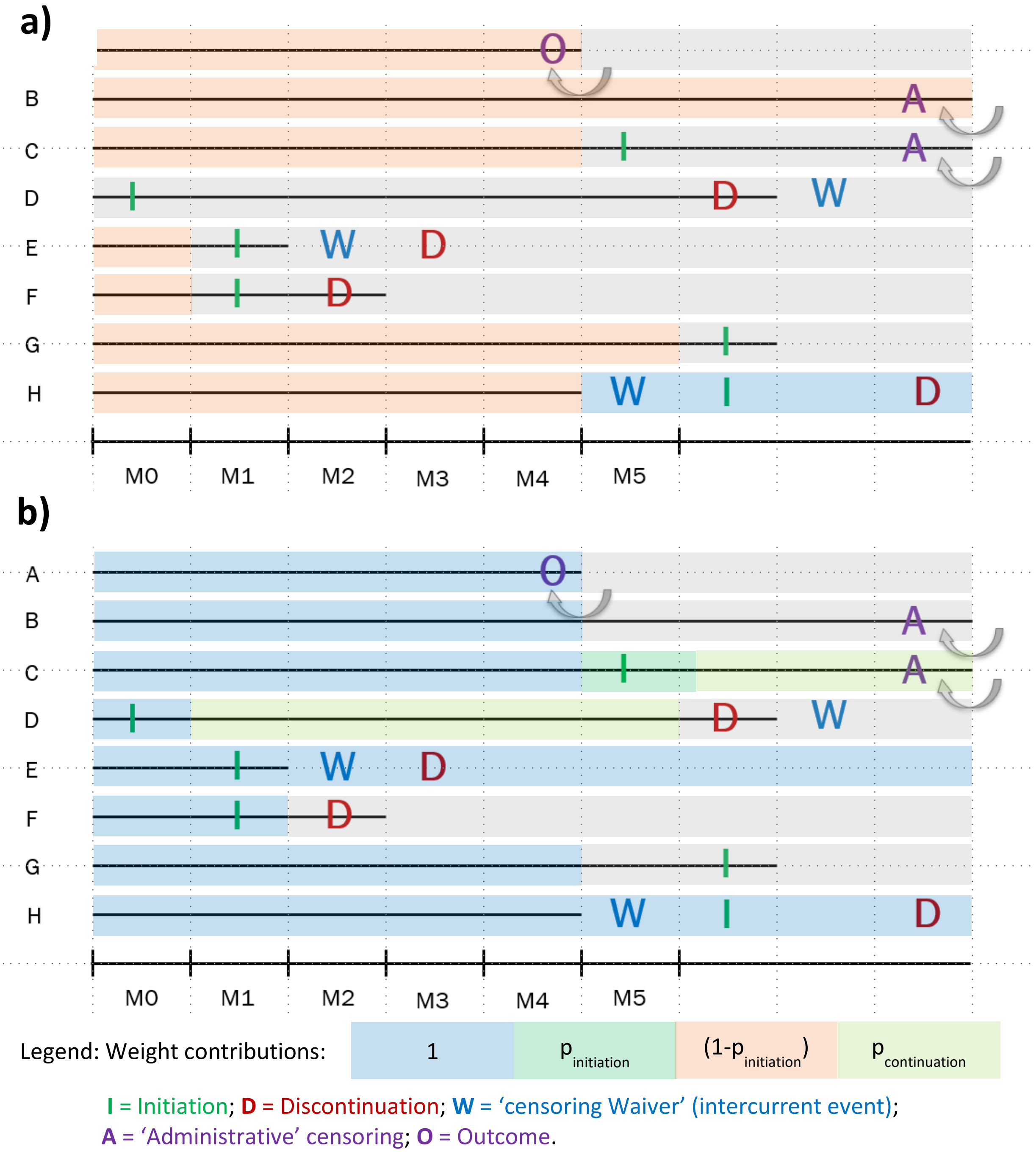

Supplemental Figure S3. Weighting process in the clone arm assigned to a) the no-treatment and b) the spironolactone treatment strategy

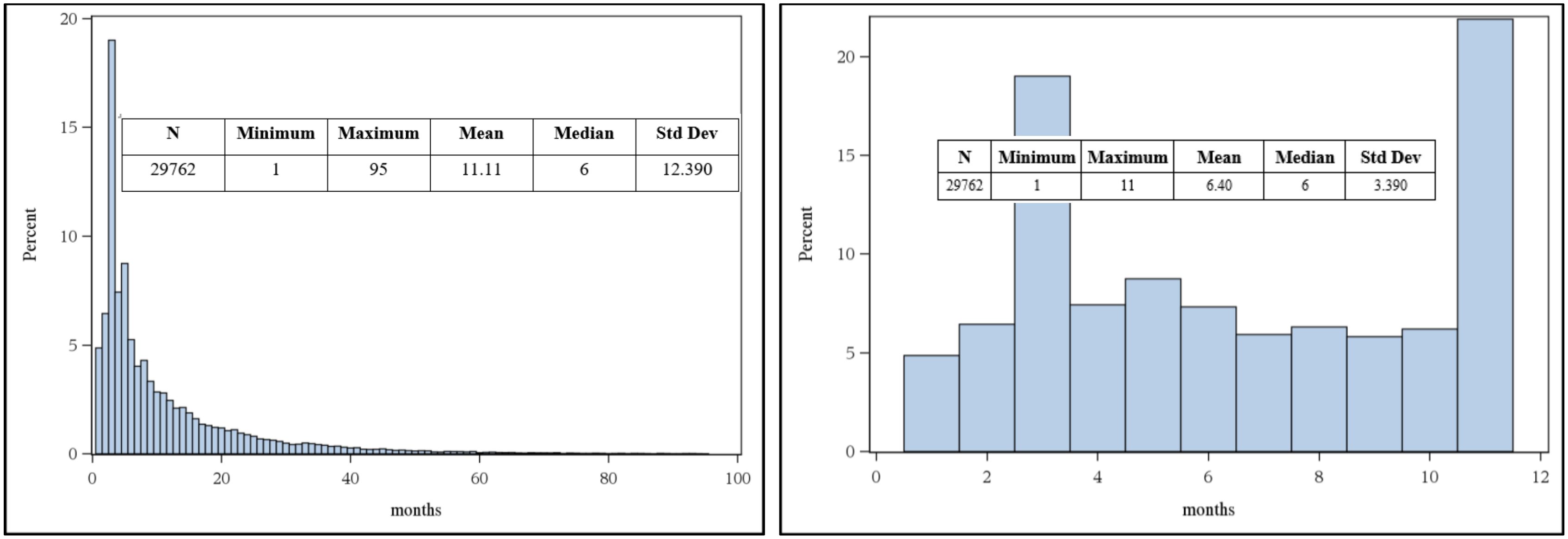

Supplemental Figure S4: Time on treatment calculated from initiation among individuals who initiated within the 6-month grace period – overall cohort.

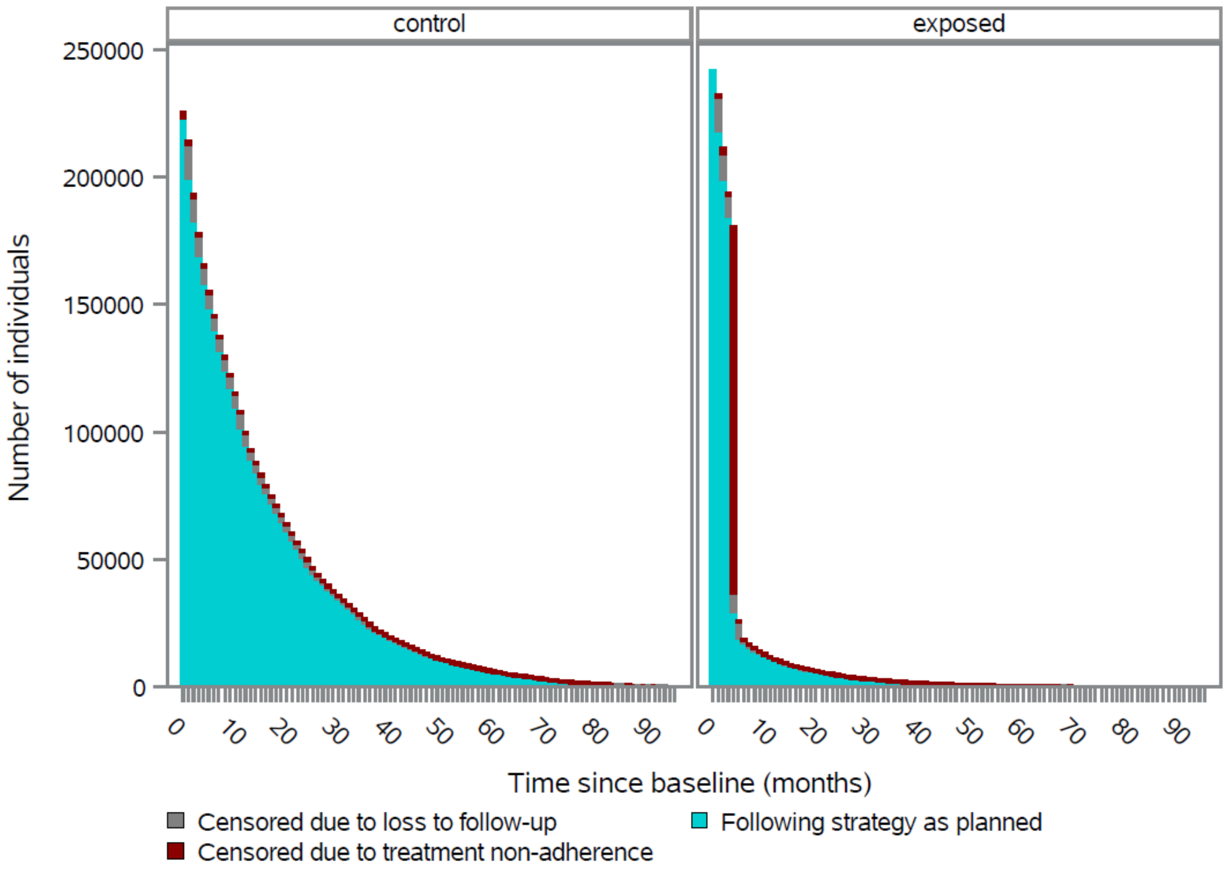

Supplemental Figure S5. Contributions of different censoring mechanisms to the reduction in sample size over time, stratified by treatment strategy, using the HF hospitalization outcome in the overall cohort as an example.

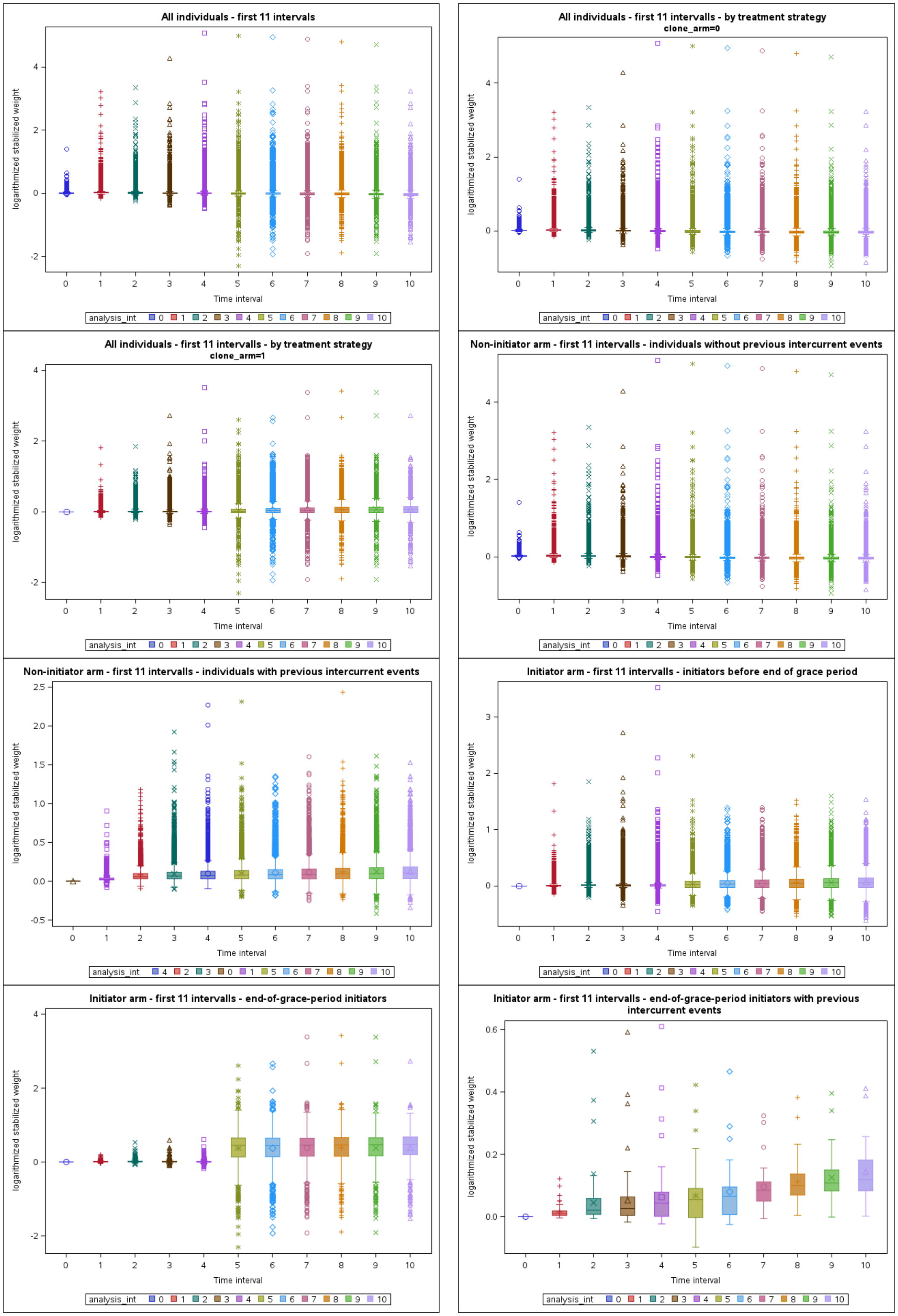

Supplemental Figure S6. Decomposition of the distribution of stabilized weights over the first 11 treatment months (i.e. all treatment intervals included in 12-month prediction of outcomes) – overall cohort, HF hospitalization outcome

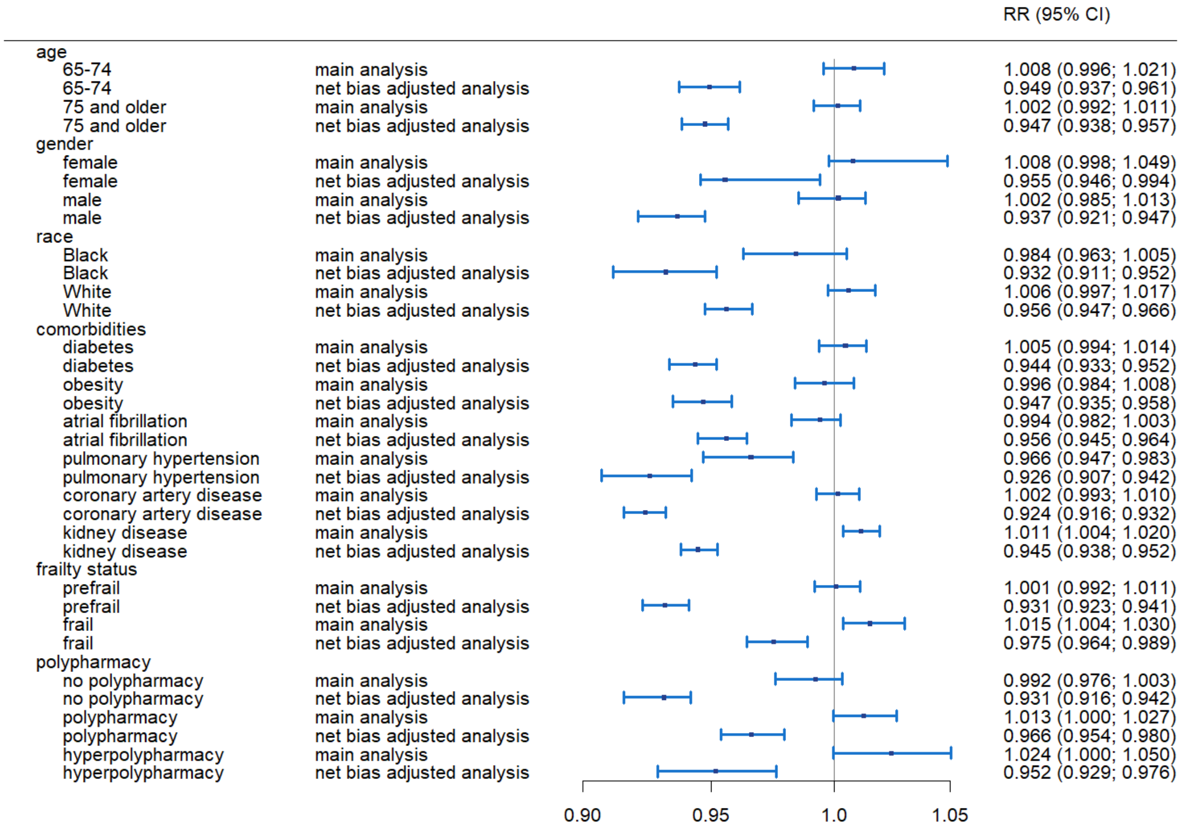

Supplemental Figure S7: Forest plot for results on heart failure hospitalizations at 12 months by subpopulation. Weighted outcome analyses using stabilized inverse probability of treatment and censoring weights truncated at the 99th percentile, with and without net bias adjustment based on the subgroup specific results for non-cardiovascular death – overall cohort

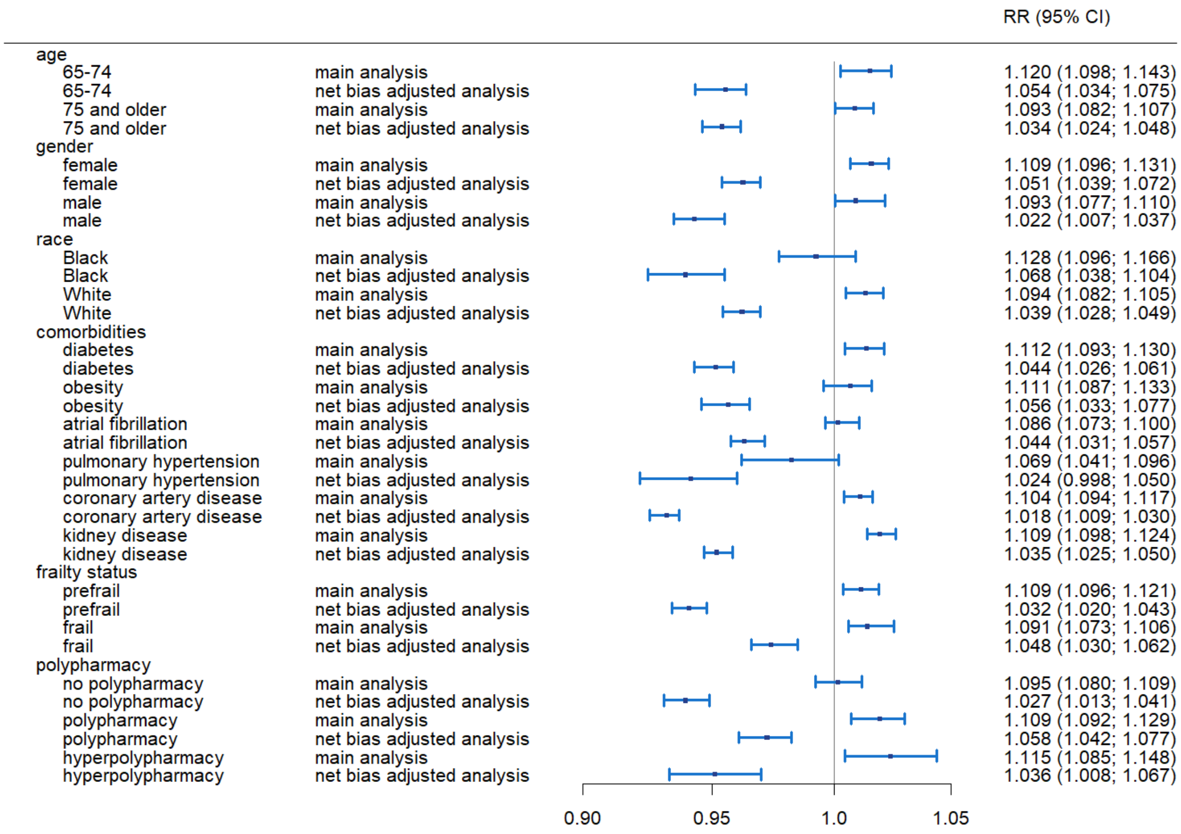

Supplemental Figure S8: Forest plot for results on the composite outcome with all-cause death at 12 months by subpopulation. Weighted outcome analyses using stabilized inverse probability of treatment and censoring weights truncated at the 99th percentile, with and without net bias adjustment based on the subgroup specific results for non-cardiovascular death – overall cohort

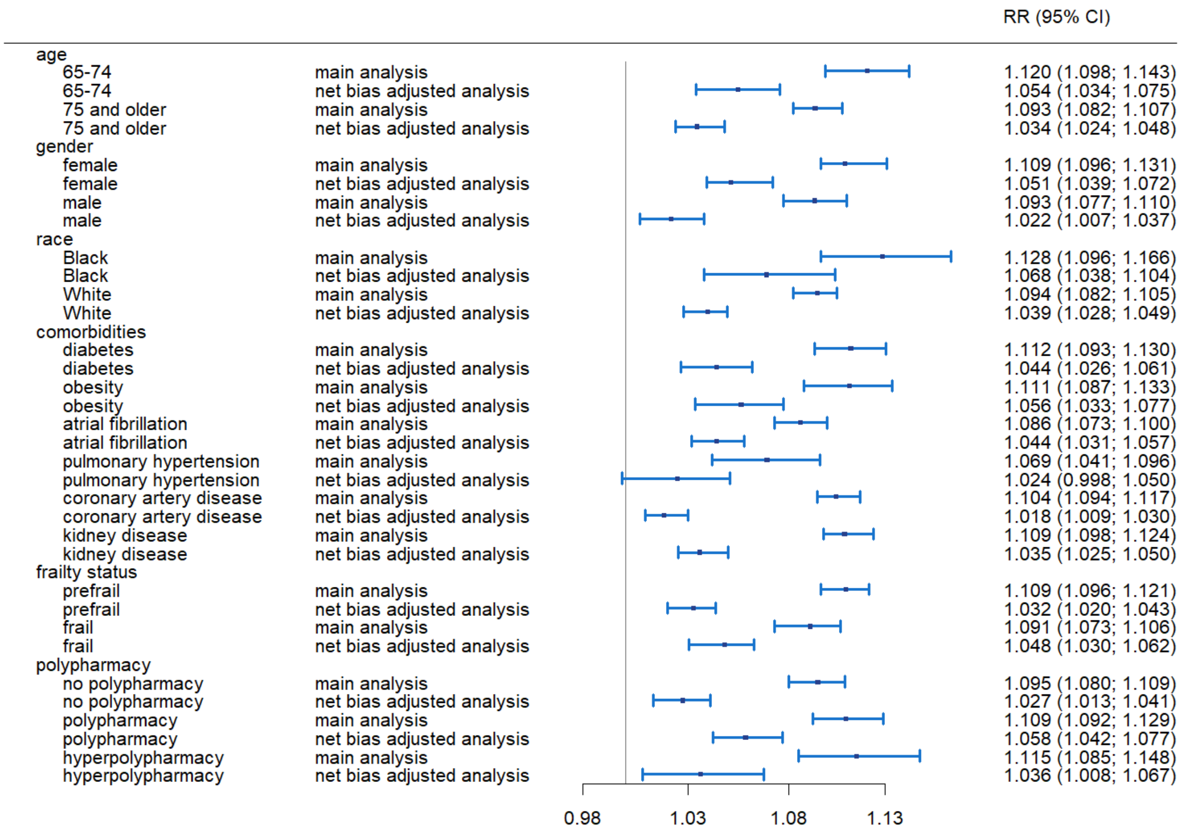

Supplemental Figure S9: Forest plot for results on all-cause death at 12 months by subpopulation. Weighted outcome analyses using stabilized inverse probability of treatment and censoring weights truncated at the 99th percentile, with and without net bias adjustment based on the subgroup specific results for non-cardiovascular death – overall cohort

| | | RR (95% CI) |
|---|---|---|
| age | | |
| 65-74 | main analysis | 0.977 (0.960; 0.994) |
| 65-74 | net bias adjusted analysis | 0.919 (0.903; 0.935) |
| 75 and older | main analysis | 0.970 (0.958; 0.984) |
| 75 and older | net bias adjusted analysis | 0.917 (0.906; 0.931) |
| gender | | |
| female | main analysis | 0.969 (0.955; 0.992) |
| female | net bias adjusted analysis | 0.918 (0.905; 0.940) |
| male | main analysis | 0.973 (0.955; 0.989) |
| male | net bias adjusted analysis | 0.909 (0.893; 0.925) |
| race | | |
| White | main analysis | 0.972 (0.960; 0.983) |
| White | net bias adjusted analysis | 0.923 (0.912; 0.933) |
| comorbidities | | |
| diabetes | main analysis | 0.975 (0.958; 0.989) |
| diabetes | net bias adjusted analysis | 0.915 (0.899; 0.929) |
| obesity | main analysis | 0.976 (0.951; 0.997) |
| obesity | net bias adjusted analysis | 0.928 (0.904; 0.948) |
| atrial fibrillation | main analysis | 0.967 (0.954; 0.979) |
| atrial fibrillation | net bias adjusted analysis | 0.930 (0.917; 0.941) |
| pulmonary hypertension | main analysis | 0.951 (0.917; 0.983) |
| pulmonary hypertension | net bias adjusted analysis | 0.911 (0.879; 0.943) |
| coronary artery disease | main analysis | 0.973 (0.959; 0.985) |
| coronary artery disease | net bias adjusted analysis | 0.897 (0.885; 0.909) |
| kidney disease | main analysis | 0.984 (0.973; 0.995) |
| kidney disease | net bias adjusted analysis | 0.919 (0.909; 0.929) |
| frailty status | | |
| prefrail | main analysis | 0.963 (0.948; 0.975) |
| prefrail | net bias adjusted analysis | 0.896 (0.882; 0.907) |
| frail | main analysis | 0.993 (0.976; 1.009) |
| frail | net bias adjusted analysis | 0.954 (0.937; 0.969) |
| polypharmacy | | |
| no polypharmacy | main analysis | 0.959 (0.946; 0.974) |
| no polypharmacy | net bias adjusted analysis | 0.900 (0.888; 0.914) |
| polypharmacy | main analysis | 0.985 (0.966; 0.999) |
| polypharmacy | net bias adjusted analysis | 0.940 (0.922; 0.953) |
| hyperpolypharmacy | main analysis | 0.981 (0.961; 1.012) |
| hyperpolypharmacy | net bias adjusted analysis | 0.911 (0.893; 0.940) |

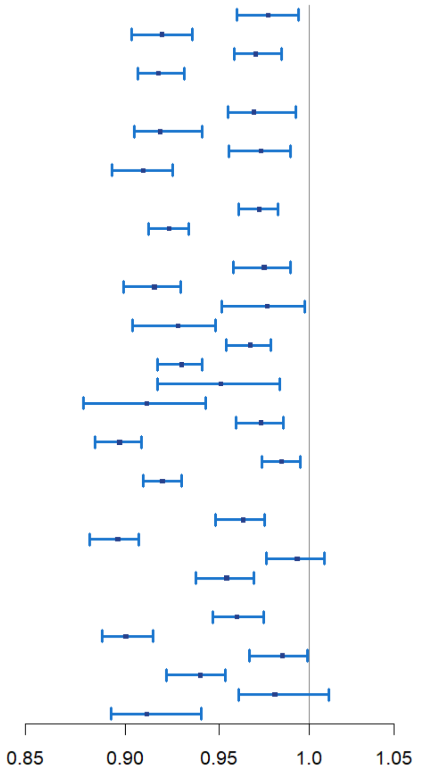

Supplemental Figure S10: Forest plot for results on the composite outcome with cardiovascular death at 12 months by subpopulation. Weighted outcome analyses using stabilized inverse probability of treatment and censoring weights truncated at the 99th percentile, with and without net bias adjustment based on the subgroup specific results for non-cardiovascular death – overall cohort

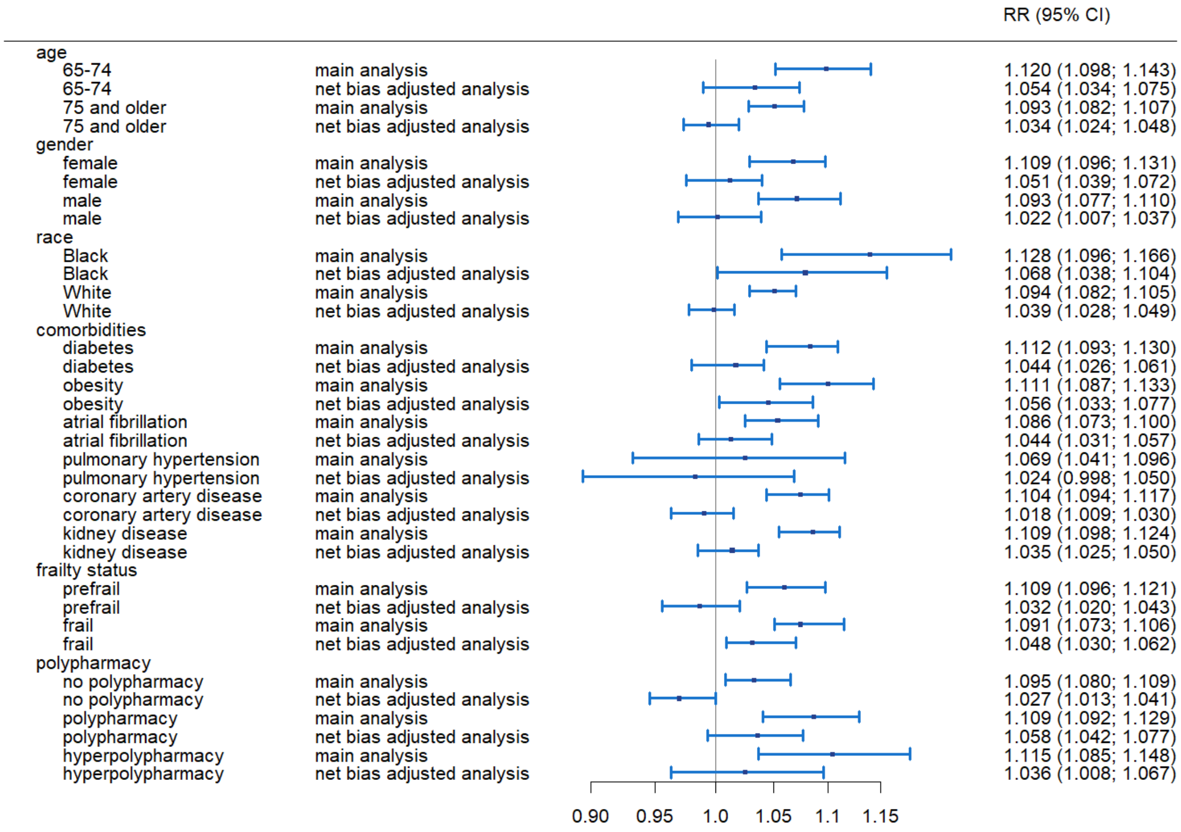

Supplemental Figure S11: Forest plot for results on cardiovascular death at 12 months by subpopulation. Weighted outcome analyses using stabilized inverse probability of treatment and censoring weights truncated at the 99th percentile, with and without net bias adjustment based on the subgroup specific results for non-cardiovascular death – overall cohort

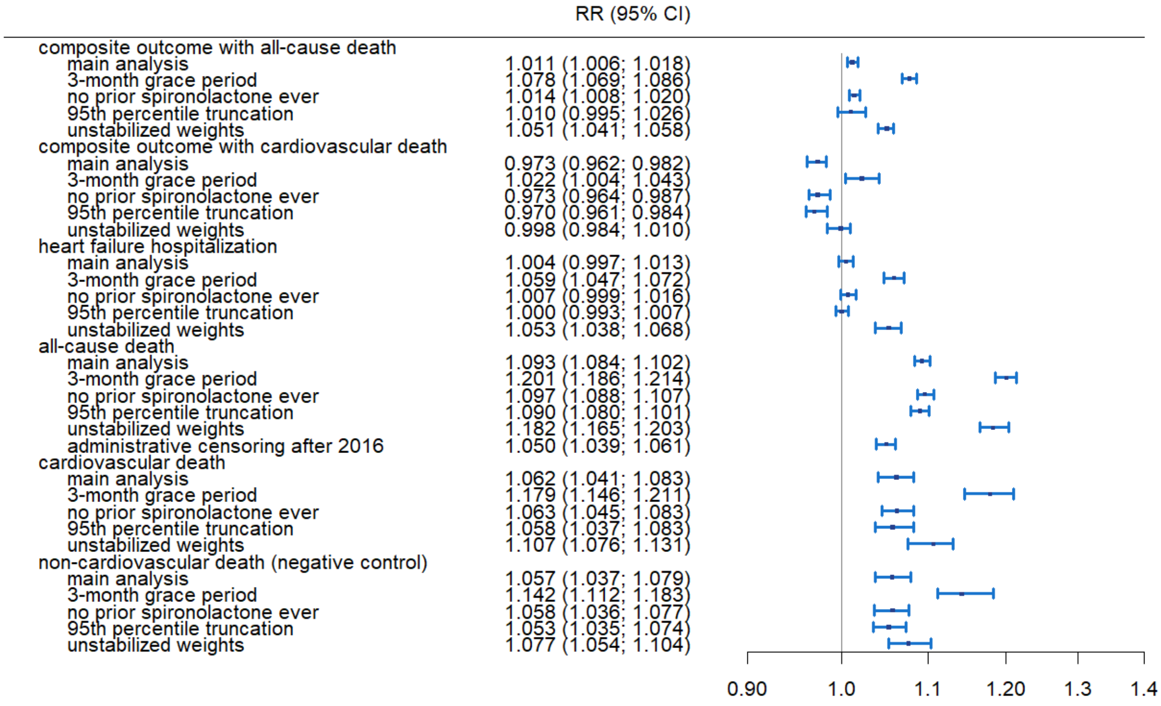

Supplemental Figure S12: Results of deterministic sensitivity analyses – overall cohort

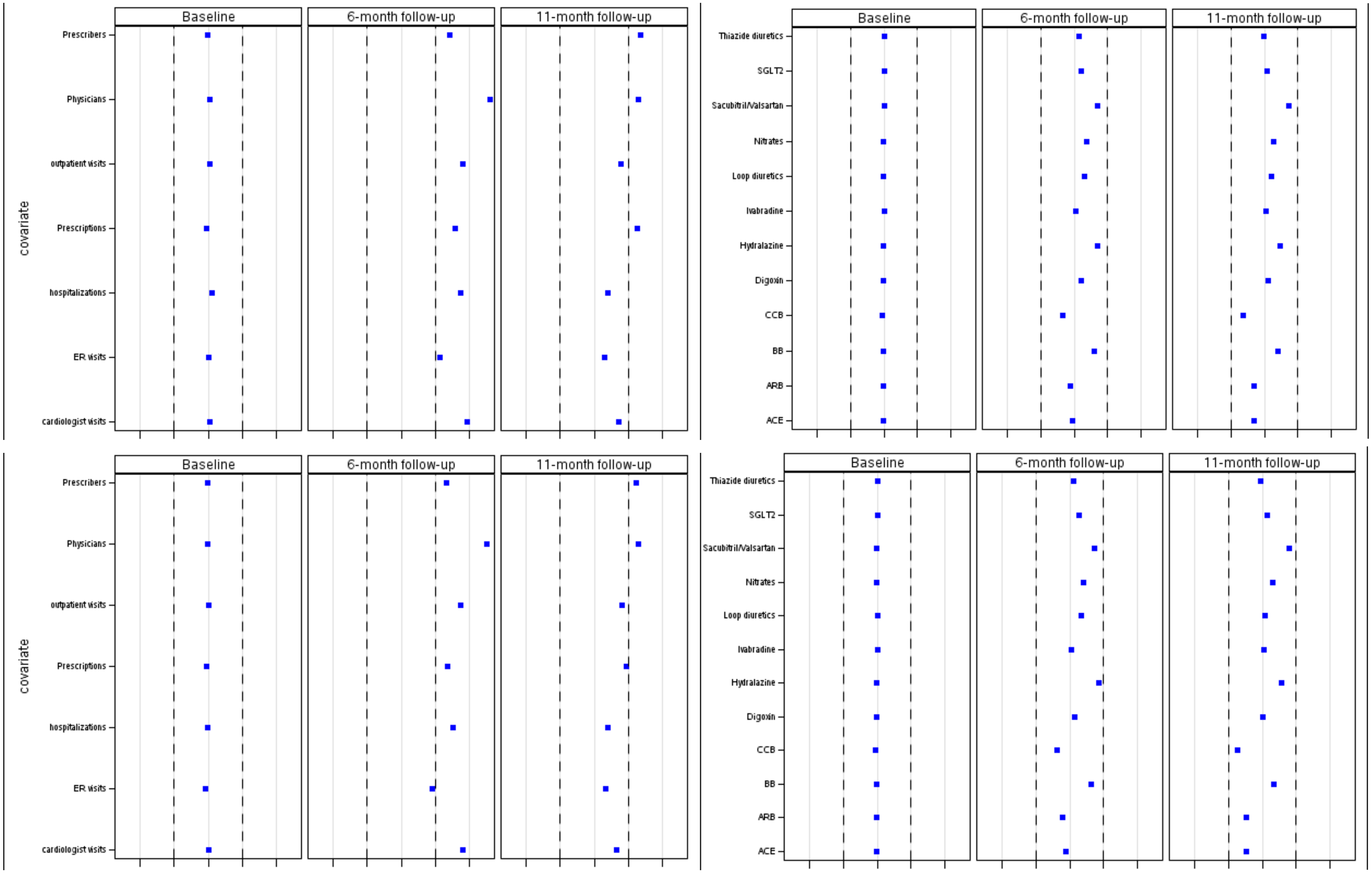

Supplemental Figure S13: Balance plots of standardized differences in time-varying covariates related to health care utilization (left) and prescription fills (right) before (top row) and after weighting with stabilized untruncated (bottom row) weights in the heart failure hospitalization dataset – overall cohort

| | RR (95% CI) |
|---|---|
| composite outcome with all-cause death | |
| main analysis | 1.011 (1.006; 1.018) |
| baseline adjusted unweighted model | 1.012 (1.006; 1.018) |
| unadjusted unweighted model | 1.013 (1.007; 1.019) |
| composite outcome with cardiovascular death | |
| main analysis | 0.973 (0.962; 0.982) |
| baseline adjusted unweighted model | 0.969 (0.958; 0.978) |
| unadjusted unweighted model | 0.970 (0.960; 0.980) |
| heart failure hospitalization | |
| main analysis | 1.004 (0.997; 1.013) |
| baseline adjusted unweighted model | 1.000 (0.994; 1.008) |
| unadjusted unweighted model | 1.007 (0.999; 1.015) |
| all-cause death | |
| main analysis | 1.093 (1.084; 1.102) |
| baseline adjusted unweighted model | 1.099 (1.089; 1.110) |
| unadjusted unweighted model | 1.089 (1.079; 1.098) |
| cardiovascular death | |
| main analysis | 1.062 (1.041; 1.083) |
| baseline adjusted unweighted model | 1.062 (1.040; 1.084) |
| unadjusted unweighted model | 1.044 (1.020; 1.068) |
| non-cardiovascular death | |
| main analysis | 1.057 (1.037; 1.079) |
| baseline adjusted unweighted model | 1.055 (1.027; 1.077) |
| unadjusted unweighted model | 1.054 (1.035; 1.082) |

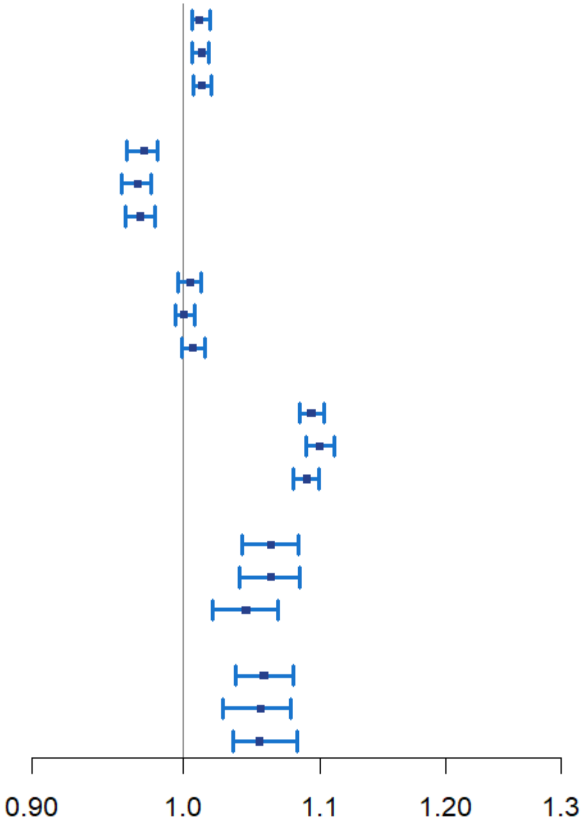

Supplemental Figure S14: Impact of controlling for observed confounders – overall cohort

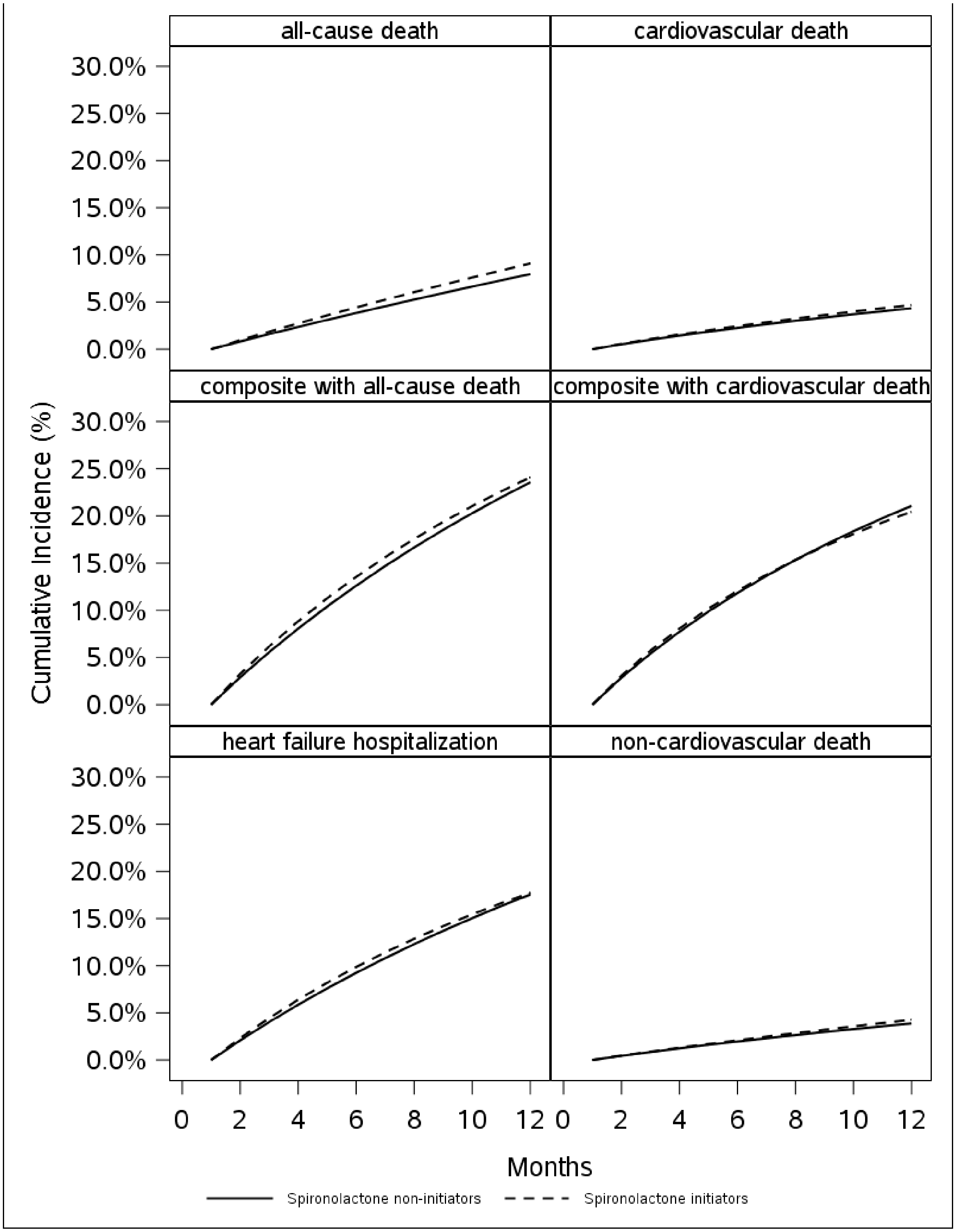

**Supplemental Figure S15. Cumulative incidence curves – benchmarking cohort**
Note: Parametric risk estimates derived from outcome models with covariate average values of the Medicare HFpEF population.

Supplemental Table S1: Outcomes operationalizations in Medicare data based on ICD-9 and ICD-10 codes

| Outcome | ICD_9_DX | ICD_10_DX |
|---|---|---|
| Heart failure | 428.xx | I50x |
| | 398.91 | I09.81x |
| | 402.01 | I11.0x |
| | 402.11 | I13.0x |
| | 402.91 | I13.2x |
| | 404.01 | |
| | 404.11 | |
| | 404.91 | |
| | 404.03 | |
| | 404.13 | |
| | 404.93 | |
| Cardiac arrest | 427.5 | I46.* |
| Hyperkalemia | 276.7x | E87.5 |

Supplemental Table S2: Attrition table/Eligibility flow

| Inclusion criteria | Medicare fee-for-service, 2012-2020 Remaining Patients | Excluded Patients |
|---|---|---|
| ≥ 1 HF diagnosis, 2013-2020 | 12,487,520 | |
| Enrolment in the 365 days prior to ≥ 1 HF diagnoses (part A, B, D) | 6,442,805 | 6,044,715 |
| No nursing home admission any time prior ≥ 1 potentially qualifying[a] HF diagnosis | 4,493,971 | 1,948,834 |
| Age ≥ 65 on ≥ 1 potentially qualifying[a] HF diagnosis, no missing gender | 4,472,169 | 21,802 |
| ≥ 1 primary HF hospitalization in the 180 days prior ≥ 1 potentially qualifying[a] HF diagnosis | 798,439 | 3,673,730 |
| No MI diagnosis within ≥ 1 of the abovementioned HF hospitalizations[b] | 768,157 | 30,282 |
| HFpEF probability >80% based on phenotyping algorithm applied to 180 days before index | 443,659 | 324,498 |
| No heart transplant, LVAD, ESRD any time prior | 395,294 | 48,365 |
| No IP/ER hyperkalemia diagnoses in the 180 days prior | 362,022 | 33,272 |
| No previous use of aldosterone antagonists, other potassium sparing medications, lithium | 352,448 | 9,574 |
| No spironolactone use in the 180 days prior | 320,881 | 31,567 |
| **Overall cohort** | **320,881** | |
| **[thereof with index date before Dec 31, 2016]** | **196,212** | |
| CCI percentile ≤95% | 310,963 | 9,918 |
| No metastatic cancer in the 365 days prior | 306,470 | 4,493 |
| No severe COPD or other pulmonary disease with steroid therapy in the 365 days prior | 283,454 | 23,016 |
| No Home O2 in the 365 days prior | 244,047 | 39,407 |
| No hospitalization for exacerbation in the 365 days prior | 219,534 | 24,513 |
| No orthostatic hypotension in the 365 days prior | 212,109 | 7,425 |
| No severe chronic kidney disease in the 365 days prior | 188,624 | 23,485 |
| No hepatic disease in the 365 days prior | 168,716 | 19,908 |
| No gastrointestinal disorder in the 365 days prior | 159,267 | 9,449 |

| | | |
|---|---|---|
| No alcohol or substance abuse in the 365 days prior | 153,762 | 5,505 |
| No dementia/Alzheimer's in the 365 days prior | 131,252 | 22,510 |
| No MI or stroke in the 90 days prior | 129,708 | 1,544 |
| No CABG in the 90 days prior | 129,011 | 697 |
| No PCI in the 30 days prior | 128,194 | 817 |
| No cardiomyopathy in the 365 days prior | 121,697 | 6,497 |
| No uncorrected valve disorder in the 365 days prior | 49,729 | 71,968 |
| **Benchmarking cohort** | **49,729** | |
| **[thereof with index date before Dec 31, 2016]** | **31,603** | |

[a] The term 'potentially qualifying' refers to the pool of all respective instances for a patient that fulfil all the previously listed inclusion criteria.

[b] If, after this step, more than one potentially qualifying HF diagnosis across the observed patient history remained, the earliest was designated as index diagnoses. After this point, eligibility was not re-evaluated, even if a subsequent inclusion criterion was not met.

Supplemental Table S3. Descriptive characteristics – comprehensive list

| Variable | | TOPCAT Americas[3,4] n = 1767 (100%) | | Benchmarking cohort n = 49,729 (100%) | | Overall cohort n = 320,881 (100%) | |
|---|---|---|---|---|---|---|---|
| Socioeconomic & Sociodemographic characteristics | | | | | | | |
| | | n | % | n | % | n | % |
| Index Year | 2013 | 0 | (0) | 9888 | (19.8) | 62452 | (19.4) |
| | 2014 | 0 | (0) | 8598 | (17.2) | 51826 | (16.1) |
| | 2015 | 0 | (0) | 7784 | (15.6) | 48105 | (14.9) |
| | 2016 | 0 | (0) | 5333 | (10.7) | 33829 | (10.5) |
| | 2017 | 0 | (0) | 4821 | (9.6) | 31777 | (9.9) |
| | 2018 | 0 | (0) | 4790 | (9.6) | 33044 | (10.2) |
| | 2019 | 0 | (0) | 4621 | (9.2) | 32923 | (10.2) |
| | 2020 | 0 | (0) | 3894 | (7.8) | 26925 | (8.3) |
| Sex | female | 882<br>565 | (50)<br>(48.9)[a] | 30514 | (61.3) | 199127 | (62.0) |
| Race | missing | 0 | (0)[a] | 2218 | (4.4) | 12847 | (4.0) |
| | White | 1384<br>867 | (78)<br>(75.5)[a] | 40303 | (81.0) | 269705 | (84.0) |
| | Black | 236 | (20.5)[a] | 6025 | (12.1) | 31532 | (9.8) |
| | Other | 46 | (4.0)[a] | 1183 | (2.3) | 6797 | (2.1) |
| Region | northeast | n.r. | n.r. | 10108 | (20.3) | 67246 | (20.9) |
| | midwest | n.r. | n.r. | 13069 | (26.2) | 78195 | (24.3) |
| | south | n.r. | n.r. | 19195 | (38.5) | 130082 | (40.5) |
| | unknown | n.r. | n.r. | 113 | (0.2) | 579 | (0.1) |
| | west | n.r. | n.r. | 7244 | (14.5) | 44779 | (13.9) |
| Medicare Subsidy Recipient | | | | 16483 | (33.1) | 102521 | (31.9) |
| Urban area | | n.r. | n.r. | 1261 | (2.5) | 7951 | (2.4) |
| Socioeconomic & Sociodemographic characteristics | | | | | | | |
| | | | | Mean | (SD) | Mean | (SD) |
| Baseline age | | 72 | NA | 79.95 | (8.49) | 80.55 | (8.37) |
| Area deprivation index | | n.r. | n.r. | 56.25 | (7.24) | 56.70 | (7.27) |
| Combined Comorbidity Index | | n.r. | n.r. | 5.32 | (2.13) | 6.82 | (2.71) |
| Claims-based Frailty Index | | n.r. | n.r. | 0.21 | (0.04) | 0.23 | (0.04) |
| Predicted probability of HFpEF | | n.r. | n.r. | 0.90 | (0.03) | 0.91 | (0.04) |
| Healthcare utilization (continuous variables) – 365 days pre-index | | | | | | | |
| Number of outpatient visits | | n.r. | n.r. | 11.05 | (8.04) | 13.84 | (9.55) |
| Number of prescribers | | n.r. | n.r. | 4.41 | (2.99) | 5.30 | (3.43) |
| Number of hospitalizations | | n.r. | n.r. | 1.37 | (0.73) | 1.75 | (1.15) |
| Number of emergency room visits | | n.r. | n.r. | 1.92 | (1.57) | 2.52 | (2.27) |
| Number of cardiologist visits | | n.r. | n.r. | 4.73 | (6.14) | 6.61 | (7.36) |
| Healthcare utilization (continuous time-varying variables) – 30 days pre-index | | | | | | | |
| Number of outpatient visits | | n.r. | n.r. | 1.12 | (1.24) | 1.36 | (1.38) |
| Number of hospitalizations | | n.r. | n.r. | 0.99 | (0.39) | 1.04 | (0.48) |
| Number of emergency room visits | | n.r. | n.r. | 0.97 | (0.61) | 1.03 | (0.70) |
| Number of cardiologist visits | | n.r. | n.r. | 1.02 | (1.44) | 1.27 | (1.75) |
| Number of HF hospitalizations | | n.r. | n.r. | 0.92 | (0.27) | 0.91 | (0.28) |

| | | | | | | |
|---|---|---|---|---|---|---|
| Number of prescribed medications | n.r. | n.r. | 4.42 | (3.39) | 4.87 | (3.58) |
| Number of prescribers | n.r. | n.r. | 1.59 | (1.30) | 1.84 | (1.43) |
| Number of outpatient physicians | n.r. | n.r. | 4.61 | (3.37) | 5.49 | (4.09) |
| Healthcare utilization for preventive measures – 365 days pre index | | | | | | |
| | N | % | N | % | N | % |
| Bone mineral density testing | n.r. | n.r. | 2734 | (5.4) | 19580 | (6.1) |
| Colonoscopy | n.r. | n.r. | 2722 | (5.4) | 24285 | (7.5) |
| Flu vaccination | n.r. | n.r. | 28623 | (57.5) | 195595 | (60.9) |
| Pneumococcal vaccination | n.r. | n.r. | 6331 | (12.7) | 45024 | (14.0) |
| Herpes zoster vaccination | n.r. | n.r. | 34 | (0.0) | 206 | (0.0) |
| Mammography | n.r. | n.r. | 6988 | (14.0) | 44560 | (13.8) |
| PAP/HPV test | n.r. | n.r. | 1048 | (2.1) | 7206 | (2.2) |
| Fecal occult blood test | n.r. | n.r. | 2691 | (5.4) | 20708 | (6.4) |
| Prostate specific antigen test | n.r. | n.r. | 3040 | (6.1) | 17530 | (5.4) |
| Medications (time-varying variables) 30 days pre-index | | | | | | |
| Angiotensin-converting enzyme inhibitors or angiotensin receptor blockers | 1395<br>945 | (79)<br>(81.7)[a] | 14313 | (28.8) | 82666 | (25.8) |
| Angiotensin-converting enzyme inhibitors | n.r. | n.r. | 7964 | (16.0) | 45137 | (14.0) |
| Angiotensin receptor blockers | n.r. | n.r. | 6541 | (13.1) | 40562 | (12.6) |
| Beta blockers | 1387<br>906 | (79)<br>(78.4)[a] | 17807 | (35.8) | 118597 | (36.9) |
| Calcium channel blockers | 682<br>453 | (39)<br>(39.2)[a] | 10314 | (20.7) | 69381 | (21.6) |
| Loop diuretics | 986 | (85.3)[a] | 15198 | (30.5) | 112124 | (34.9) |
| Thiazide diuretics | | | 2739 | (5.5) | 17544 | (5.4) |
| Hydralazine | n.r. | n.r. | 1896 | (3.8) | 14583 | (4.5) |
| Nitrates | n.r. | n.r. | 3685 | (7.4) | 27768 | (8.6) |
| Digoxin | n.r. | n.r. | 1507 | (3.0) | 12345 | (3.8) |
| Sacubitril/Valsartan | n.r. | n.r. | 55 | (0.1) | 509 | (0.1) |
| SGLT2 inhibitors | n.r. | n.r. | 152 | (0.3) | 791 | (0.2) |
| Comorbidities 365 days pre index | | | | | | |
| COPD | 291<br>206 | (16)<br>(17.8) )[a] | 419 | (0.8) | 152813 | (47.6) |
| Pulmonary disease | n.r. | n.r. | 14961 | (30.0) | 145776 | (45.4) |
| Valve disorder | n.r. | n.r. | 1323 | (2.6) | 196859 | (61.3) |
| Cardiac arrest | n.r. | n.r. | 264 | (0.5) | 2578 | (0.8) |
| Atrial fibrillation | 743<br>466 | (42)<br>(40.3)[a] | 26563 | (53.4) | 191079 | (59.5) |
| Obesity | n.r. | n.r. | 19418 | (39.0) | 116079 | (36.1) |
| Diabetes | 788<br>584 | (45)<br>(50.5)[a] | 31095 | (62.5) | 187418 | (58.4) |
| Pulmonary hypertension | n.r. | n.r. | 3141 | (6.3) | 38212 | (11.9) |
| Renal disease | 855<br>586 | (48)<br>(50.7)[a] | 28645 | (57.6) | 209620 | (65.3) |
| Coronary artery disease | n.r. | n.r. | 30576 | (61.4) | 221398 | (68.9) |
| Angioedema | n.r. | n.r. | 84 | (0.1) | 675 | (0.2) |
| Anemia | 517 | (45.2)[a] | 21796 | (43.8) | 177893 | (55.4) |
| Depression | n.r. | n.r. | 9080 | (18.2) | 74750 | (23.2) |
| Diabetic nephropathy | n.r. | n.r. | 8855 | (17.8) | 62709 | (19.5) |
| Endocarditis | n.r. | n.r. | 221 | (0.4) | 9931 | (3.0) |
| Hypertensive nephropathy | n.r. | n.r. | 17632 | (35.4) | 140331 | (43.7) |

| | | | | | | |
|---|---|---|---|---|---|---|
| Hyperlipidemia | n.r. | n.r. | 41122 | (82.6) | 272281 | (84.8) |
| Hypertension | n.r. | n.r. | 48454 | (97.4) | 314091 | (97.8) |
| Hypokalemia | n.r. | n.r. | 10057 | (20.2) | 79083 | (24.6) |
| Hypotension | n.r. | n.r. | 3811 | (7.6) | 47615 | (14.8) |
| Other disrythmias | n.r. | n.r. | 18800 | (37.8) | 146233 | (45.5) |
| Peripheral artery disease | n.r. | n.r. | 4895 | (9.8) | 38217 | (11.9) |
| Psychosis | n.r. | n.r. | 991 | (1.9) | 25379 | (7.9) |
| Rheumatic heart disease | n.r. | n.r. | 397 | (0.7) | 47164 | (14.6) |
| Sleep apnea | n.r. | n.r. | 10574 | (21.2) | 77011 | (23.9) |
| Stable angina | 486 | (28) | 3617 | (7.2) | 27759 | (8.6) |
| Unstable angina | | | 3953 | (7.9) | 32182 | (10.0) |
| Smoking | n.r. | n.r. | 11287 | (22.6) | 93629 | (29.1) |
| Thyroid dysfunction | n.r. | n.r. | 17166 | (34.5) | 123239 | (38.4) |
| Hemorrhage | n.r. | n.r. | 315 | (0.6) | 3343 | (1.0) |
| Outpatient HF index diagnosis | n.r. | n.r. | 48797 | (98.1) | 310746 | (96.8) |
| Metastatic cancer | n.r. | n.r. | 0 | (0.0) | 10786 | (3.3) |
| Cardiomyopathy | 0 | (0.0) | 0 | (0.0) | 18505 | (5.7) |
| Home oxygen | 0 | (0.0) | 0 | (0.0) | 48285 | (15.0) |
| Hospitalization for exacerbation | 0 | (0.0) | 0 | (0.0) | 50787 | (15.8) |
| Orthostatic hypotension | 0 | (0.0) | 0 | (0.0) | 11449 | (3.5) |
| Severe renal disease | 0 | (0.0) | 0 | (0.0) | 34853 | (10.8) |
| Hepatic disease | 0 | (0.0) | 0 | (0.0) | 37747 | (11.7) |
| Gastrointestinal disorder | 0 | (0.0) | 0 | (0.0) | 22497 | (7.0) |
| Alcohol (ab)use | n.r. | n.r. | 0 | (0.0) | 7800 | (2.4) |
| Drug (ab)use | n.r. | n.r. | 0 | (0.0) | 8623 | (2.6) |
| Alzheimers/Dementia | n.r. | n.r. | 0 | (0.0) | 44514 | (13.8) |
| MI (365-91 days pre-index) | n.r. | n.r. | 419 | (0.8) | 4842 | (1.5) |
| Stroke (365-91 days pre-index) | n.r. | n.r. | 322 | (0.6) | 3653 | (1.1) |
| MI (90 days pre-index) | 0 | (0.0) | 0 | (0.0) | 2655 | (0.8) |
| Stroke (90 days pre-index) | 0 | (0.0) | 0 | (0.0) | 1686 | (0.5) |
| Procedures | | | | | | |
| CABG (90 days pre-index) | 0 | (0.0) | 0 | (0.0) | 1602 | (0.4) |
| PCI (30 days pre-index) | 0 | (0.0) | 0 | (0.0) | 2260 | (0.7) |
| Cardioverter defibrillator (365 days pre-index) | n.r. | n.r. | 1306 | (2.6) | 10792 | (3.3) |
| Cardiac resychronization therapy (365 days pre-index) | n.r. | n.r. | 1560 | (3.1) | 12975 | (4.0) |
| Valve correction (365 days pre-index) | n.r. | n.r. | 501 | (1.0) | 1725 | (0.5) |
| Valve replacement (365 days pre-index) | n.r. | n.r. | 1048 | (2.1) | 3557 | (1.1) |
| CABG (365-91 days pre-index) | n.r. | n.r. | 245 | (0.4) | 1324 | (0.4) |
| PCI (365-31 days pre-index) | n.r. | n.r. | 996 | (2.0) | 9023 | (2.8) |
| Further medications - 30 days pre-index | | | | | | |
| Statins | 753 | (65.1)[a] | 13001 | (26.1) | 85914 | (26.7) |
| Insulin | 379 | (21) | 4909 | (9.8) | 27582 | (8.5) |
| Hypoglycemic agents | n.r. | n.r. | 9646 | (19.3) | 51242 | (15.9) |
| Antiplatelets | n.r. | n.r. | 3535 | (7.1) | 25427 | (7.9) |
| Anticoagulants | n.r. | n.r. | 8271 | (16.6) | 59746 | (18.6) |
| Gout medications | n.r. | n.r. | 2209 | (4.4) | 15811 | (4.9) |
| COPD medications | n.r. | n.r. | 4074 | (8.1) | 38641 | (12.0) |
| NSAIDs | n.r. | n.r. | 3170 | (6.3) | 17072 | (5.3) |

| | | | | | | |
|---|---|---|---|---|---|---|
| Pressor amines | n.r. | n.r. | 21 | (0.0) | 157 | (0.0) |
| Barbiturates | n.r. | n.r. | 49 | (0.0) | 181 | (0.0) |
| Narcotics | n.r. | n.r. | 7300 | (14.6) | 53855 | (16.7) |
| Skeletal muscle relaxants | n.r. | n.r. | 843 | (1.6) | 5893 | (1.8) |
| Oral steriods | n.r. | n.r. | 243 | (0.4) | 5536 | (1.7) |
| Further medications - 365-31 days pre-index | | | | | | |
| Statins | n.r. | n.r. | 30527 | (61.3) | 200064 | (62.3) |
| Insulin | n.r. | n.r. | 9737 | (19.5) | 55951 | (17.4) |
| Hypoglycemic agents | n.r. | n.r. | 19265 | (38.7) | 105652 | (32.9) |
| Antiplatelets | n.r. | n.r. | 8100 | (16.2) | 57305 | (17.8) |
| Anticoagulants | n.r. | n.r. | 15104 | (30.3) | 112985 | (35.2) |
| Gout medications | n.r. | n.r. | 4924 | (9.9) | 35764 | (11.1) |
| COPD medications | n.r. | n.r. | 8944 | (17.9) | 85775 | (26.7) |
| NSAIDs | n.r. | n.r. | 10109 | (20.3) | 60169 | (18.7) |
| Pressor amines | n.r. | n.r. | 140 | (0.2) | 1149 | (0.3) |
| Barbiturates | n.r. | n.r. | 61 | (0.1) | 296 | (0.0) |
| Narcotics | n.r. | n.r. | 17370 | (34.9) | 123630 | (38.5) |
| Skeletal muscle relaxants | n.r. | n.r. | 2986 | (6.0) | 21823 | (6.8) |
| Calcium channel blockers | n.r. | n.r. | 23014 | (46.2) | 152308 | (47.4) |
| Oral steriods | n.r. | n.r. | 1364 | (2.7) | 29139 | (9.0) |

[a] Hospitalization stratum of TOPCAT Americas[5] (n=1156)

**Supplemental Table S4. Distribution of spironolactone initiation in all individuals initiating until end of month 6 after index**

| Days after index | Overall cohort, N (%) | | Benchmarking cohort, N (%) | |
|---|---|---|---|---|
| 0-180 | 29,762 | (100%) | 4,736 | (100%) |
| 0-30 | 19164 | (64.3) | 3145 | (66.4) |
| 31-60 | 3566 | (11.9) | 505 | (10.6) |
| 61-90 | 2428 | (8.1) | 347 | (7.3) |
| 91-120 | 1826 | (6.1) | 301 | (6.3) |
| 121-150 | 1481 | (4.9) | 235 | (4.9) |
| 151-180 | 1297 | (4.3) | 203 | (4.2) |

Supplemental Table S5. Person-time in months contributed to each treatment strategy (cloned and censored population before weighting)

| Outcome | Spironolactone non-user clones | Spironolactone user clones |
|---|---|---|
| **Overall cohort** | | |
| Heart failure hospitalization | 3,498,834 | 1,289,284 |
| Composite with all-cause death | 3,490,188 | 1,286,056 |
| All-cause death | 4,390,549 | 1,543,586 |
| Composite with cardiovascular death | 1,687,971 | 711,765 |
| Cardiovascular death | 2,040,587 | 832,025 |
| Hyperkalemia hospitalization | 4,105,608 | 1,366,580 |
| Non-cardiovascular death | 2,040,611 | 832,049 |
| **Benchmarking cohort** | | |
| Heart failure hospitalization | 687,235 | 221,924 |
| Composite with all-cause death | 685,804 | 221,528 |
| All-cause death | 837,403 | 257,148 |
| Composite with cardiovascular death | 328,950 | 126,293 |
| Cardiovascular death | 386,393 | 143,325 |
| Hyperkalemia hospitalization | 790,465 | 229,120 |
| Non-cardiovascular death | 386,397 | 143,330 |

Supplemental Table S6: Size of analysis datasets for each outcome at index - overall cohort.

| | Outcome | | | | | | |
|---|---|---|---|---|---|---|---|
| | HF hospi-talisation | Composite with all-cause death | All-cause death | Composite with cardio-vascular death | Cardio-vascular death | Hyperka-lemia hospi-talisation | Non-cardio-vascular death |
| Full cohort | 320,881 | 320,881 | 320,881 | 196,212 | 196,212 | 320,881 | 196,212 |
| - At maximum one observed interval | 65,973 | 65,973 | 65,973 | 42,353 | 42,353 | 65,973 | 42,353 |
| - Outcomes in the first interval | 16,624 | 30,985 | 15,091 | 15,158 | 5,427 | 5,061 | 4,051 |
| - At maximum one observed interval & outcome in the first interval | 3,379 | 17,217 | 14,788 | 7,130 | 5,314 | 1,384 | 3,962 |
| More than one observed interval | 254,908 | 254,908 | 254,908 | 151,406 | 151,406 | 254,908 | 151,406 |
| – outcome in the first interval | 13,245 | 13,768 | 303 [a] | 7,951 | 113 [b] | 3,677 | 89 [c] |
| More than one observed interval and no outcome in the first interval: Starting sample for (administrative or competing event) censoring probability calculation; Clone Arm 1 | 241,663 | 241,140 | 254,605 | 143,455 | 151,293 | 251,231 | 151,293 |
| - intercurrent event in the first interval | 4,650 | 4,544 | 6,126 | 2,741 | 3,629 | 2,489 | 3,634 |
| Starting sample for treatment probability calculation | 237,013 | 236,596 | 248,479 | 143,090 | 150,117 | 248,742 | 150,136 |
| - Non-waivered initiators in interval 0 (baseline) | 16,200 | 16,183 | 17,486 | 9,306 | 10,078 | 17,499 | 10,078 |
| Starting sample after censoring non-waivered initiators in interval 0: Clone Arm 0 | 225,463 | 224,957 | 237,119 | 134,149 | 141,215 | 233,732 | 141,215 |

[a] There were 303 individuals with all-cause death outcomes recorded for both the first and second observed interval. For the all-cause death analysis, we removed those individuals from the dataset, whereas for analyses of other outcomes they were considered as “administratively censored” after the first observed interval, unless they had a record for the target outcome of the respective analysis during the first interval.
[b] There were 113 individuals with cardiovascular death outcomes recorded for both the first and second observed interval. For the cardiovascular death analysis, we removed those individuals from the dataset, whereas for analyses of other outcomes they were considered as “administratively censored” after the first observed interval, unless they had a record for the target outcome of the respective analysis during the first interval.
[c] There were 89 individuals with non-cardiovascular death outcomes recorded for both the first and second observed interval. For the non-cardiovascular death analysis, we removed those individuals from the dataset, whereas for analyses of other outcomes they were considered as

“administratively censored” after the first observed interval, unless they had a record for the target outcome of the respective analysis during the first interval.

**Supplemental Table S7: Reasons for end of follow-up in the cloned and censored dataset before weighting – overall cohort**

| | All | | Clone arm | | | |
|---|---|---|---|---|---|---|
| | | | Non-initiator | | Spironolactone user | |
| Reason for end of follow-up | n | (%) | n | (%) | n | (%) |
| in HF hospitalization analysis | | | | | | |
| All | 467,126 | (100.0) | 225,463 | (48.2) | 241,663 | (51.7) |
| Treatment deviation | 165,246 | (35.3) | 13,486 | (5.9) | 151,760 | (62.7) |
| Outcome | 99,880 | (21.3) | 65,966 | (29.2) | 33,914 | (14.0) |
| thereof with previous intercurrent events | 9,044 | (1.9) | 5,460 | (1.1) | 3,584 | (0.7) |
| Administrative censoring / competing risk | 201,889 | (43.2) | 146,011 | (64.7) | 55,878 | (23.1) |
| thereof with previous intercurrent events | 23,352 | (4.9) | 14,480 | (3.0) | 8,872 | (1.8) |
| in composite outcome with all-cause death analysis | | | | | | |
| All | 466,097 | (100.0) | 224,957 | (48.2) | 241,140 | (51.7) |
| Treatment deviation | 165,023 | (35.4) | 13,446 | (5.9) | 151,577 | (62.8) |
| Outcome | 150,704 | (32.3) | 98,956 | (43.9) | 51,748 | (21.4) |
| thereof with previous intercurrent events | 15,964 | (3.4) | 9,634 | (2.0) | 6,330 | (1.3) |
| Administrative censoring / competing risk | 150,238 | (32.2) | 112,555 | (50.0) | 37,683 | (15.6) |
| thereof with previous intercurrent events | 15,851 | (3.4) | 9,962 | (2.1) | 5,889 | (1.2) |
| in all-cause death analysis | | | | | | |
| All | 491,724 | (100.0) | 237,119 | (48.2) | 254,605 | (51.7) |
| Treatment deviation | 196,710 | (40.0) | 22,832 | (9.6) | 173,878 | (68.2) |
| Outcome | 78,930 | (16.0) | 52,428 | (22.1) | 26,502 | (10.4) |
| thereof with previous intercurrent events | 16,265 | (3.3) | 10,076 | (2.0) | 6,189 | (1.2) |
| Administrative censoring / competing risk | 215,989 | (43.9) | 161,859 | (68.2) | 54,130 | (21.2) |
| thereof with previous intercurrent events | 35,038 | (7.1) | 22,703 | (4.6) | 12,335 | (2.5) |
| in composite outcome with cardiovascular death analysis | | | | | | |
| All | 277,604 | (100.0) | 134,149 | (48.3) | 143,455 | (51.6) |
| Treatment deviation | 94,467 | (34.0) | 6,596 | (4.9) | 87,871 | (61.2) |
| Outcome | 69,113 | (24.8) | 44,002 | (32.8) | 25,111 | (17.5) |
| thereof with previous intercurrent events | 5,911 | (2.1) | 3,419 | (1.2) | 2,492 | (0.8) |
| Administrative censoring / competing risk | 113,966 | (41.0) | 83,551 | (62.2) | 30,415 | (21.2) |
| thereof with previous intercurrent events | 11,476 | (4.1) | 6,925 | (2.4) | 4,551 | (1.6) |

| | | | | | | |
|---|---|---|---|---|---|---|
| in cardiovascular death analysis | | | | | | |
| All | 292,508 | (100.0) | 141,215 | (48.2) | 151,293 | (51.7) |
| Treatment deviation | 111,501 | (38.1) | 10,991 | (7.7) | 100,510 | (66.4) |
| Outcome | 22,440 | (7.6) | 14,389 | (10.1) | 8,051 | (5.3) |
| thereof with previous intercurrent events | 3,871 | (1.3) | 2,307 | (0.7) | 1,564 | (0.5) |
| Administrative censoring / competing risk | 158,550 | (54.2) | 115,835 | (82.0) | 42,715 | (28.2) |
| thereof with previous intercurrent events | 23,080 | (7.8) | 14,205 | (4.8) | 8,875 | (3.0) |
| in hyperkalemia hospitalization analysis | | | | | | |
| All | 484,963 | (100.0) | 233,732 | (48.1) | 251,231 | (51.8) |
| Treatment deviation | 196,710 | (40.5) | 22,832 | (9.7) | 173,878 | (69.2) |
| Outcome | 37,118 | (7.6) | 24,939 | (10.6) | 12,179 | (4.8) |
| thereof with previous intercurrent events | 3,554 | (0.7) | 2,276 | (0.4) | 1,278 | (0.2) |
| Administrative censoring / competing risk | 251,109 | (51.7) | 185,961 | (79.5) | 65,148 | (25.9) |
| thereof with previous intercurrent events | 15,778 | (3.2) | 10,311 | (2.1) | 5,467 | (1.1) |
| in non-cardiovascular death analysis | | | | | | |
| All | 292,556 | (100.0) | 141,239 | (48.2) | 151,317 | (51.7) |
| Treatment deviation | 111,501 | (38.1) | 10,991 | (7.7) | 100,510 | (66.4) |
| Outcome | 20,768 | (7.0) | 13,442 | (9.5) | 7,326 | (4.8) |
| thereof with previous intercurrent events | 4,300 | (1.4) | 2,577 | (0.8) | 1,723 | (0.5) |
| Administrative censoring / competing risk | 160,113 | (54.7) | 116,806 | (82.6) | 43,307 | (28.6) |
| thereof with previous intercurrent events | 22,661 | (7.7) | 13,940 | (4.7) | 8,721 | (2.9) |

Supplemental Table S8. Weight distributions by outcome – overall cohort, entire follow-up

| Outcome / Weight type | N** | Mean | Std Dev | Minimum | Median | Maximum | 90th Pctl | 95th Pctl | 99th Pctl |
|---|---|---|---|---|---|---|---|---|---|
| Heart failure hospitalizations | | | | | | | | | |
| Unstabilized | 4788118 | 3.912 | 72.781 | 1.000 | 1.464 | 32410.291 | 4.126 | 6.673 | 24.464 |
| Stabilized | 4788118 | 1.014 | 0.279 | 0.100 | 0.999 | 160.877 | 1.087 | 1.200 | 1.772 |
| Stabilized, truncated* | 4788118 | 1.006 | 0.133 | 0.100 | 0.999 | 1.772 | 1.087 | 1.200 | 1.772 |
| Composite w all-cause death | | | | | | | | | |
| Unstabilized | 4776244 | 2.738 | 28.965 | 1.000 | 1.334 | 10789.622 | 3.180 | 4.805 | 15.141 |
| Stabilized | 4776244 | 1.005 | 0.162 | 0.102 | 1.000 | 94.391 | 1.061 | 1.125 | 1.405 |
| Stabilized, truncated* | 4776244 | 1.001 | 0.080 | 0.102 | 1.000 | 1.405 | 1.061 | 1.125 | 1.405 |
| All-cause death | | | | | | | | | |
| Unstabilized | 5934135 | 15.255 | 10661.106 | 1.000 | 1.477 | 16846225.531 | 4.228 | 6.819 | 25.176 |
| Stabilized | 5934135 | 2.667 | 1396.777 | 0.045 | 0.998 | 1993298.705 | 1.090 | 1.187 | 1.580 |
| Stabilized, truncated* | 5934135 | 1.003 | 0.112 | 0.045 | 0.998 | 1.580 | 1.090 | 1.187 | 1.580 |
| Composite with cardiovascular death | | | | | | | | | |
| Unstabilized | 2399736 | 2.052 | 19.196 | 1.000 | 1.283 | 5767.920 | 2.203 | 2.762 | 5.014 |
| Stabilized | 2399736 | 1.006 | 0.125 | 0.178 | 0.999 | 15.739 | 1.057 | 1.126 | 1.439 |
| Stabilized, truncated* | 2399736 | 1.002 | 0.082 | 0.178 | 0.999 | 1.439 | 1.057 | 1.126 | 1.439 |
| Cardiovascular death | | | | | | | | | |
| Unstabilized | 2872612 | 21.388 | 20679.781 | 1.000 | 1.391 | 24675638.925 | 2.725 | 3.557 | 7.037 |
| Stabilized | 2872612 | 4.070 | 3313.417 | 0.044 | 0.997 | 3951017.708 | 1.085 | 1.198 | 1.618 |
| Stabilized, truncated* | 2872612 | 1.005 | 0.116 | 0.044 | 0.997 | 1.618 | 1.085 | 1.198 | 1.618 |
| Hyperkalemia hospitalization | | | | | | | | | |
| Unstabilized | 5472188 | 5.215 | 227.813 | 1.000 | 1.590 | 188025.069 | 5.177 | 8.747 | 34.479 |
| Stabilized | 5472188 | 1.005 | 0.230 | 0.046 | 0.998 | 300.224 | 1.089 | 1.180 | 1.615 |
| Stabilized, truncated* | 5472188 | 0.999 | 0.119 | 0.046 | 0.998 | 1.615 | 1.089 | 1.180 | 1.615 |
| Non-cardiovascular death | | | | | | | | | |
| Unstabilized | 2872660 | 3.384 | 952.452 | 1.000 | 1.399 | 1130220.487 | 2.760 | 3.611 | 7.128 |
| Stabilized | 2872660 | 1.241 | 234.817 | 0.044 | 0.998 | 278381.713 | 1.084 | 1.192 | 1.587 |
| Stabilized, truncated* | 2872660 | 1.005 | 0.111 | 0.044 | 0.998 | 1.587 | 1.084 | 1.192 | 1.587 |

**Supplemental Table S8. Weight distributions by outcome – overall cohort, entire follow-up**

| Outcome / Weight type | N** | Mean | Std Dev | Minimum | Median | Maximum | 90th Pctl | 95th Pctl | 99th Pctl |
|---|---|---|---|---|---|---|---|---|---|

*at the 99th percentile

** the number of treatment intervals included in the outcome model (weights are time- and person-specific)

Supplemental Table S9. Weight distributions by outcome – overall cohort, 12 months follow-up

| Outcome / Weight type | N** | Mean | Std Dev | Minimum | Median | Maximum | 90th Pctl | 95th Pctl | 99th Pctl |
|---|---|---|---|---|---|---|---|---|---|
| Heart failure hospitalizations | | | | | | | | | |
| Unstabilized | 2830403 | 1.586 | 16.977 | 1.000 | 1.180 | 13326.472 | 1.629 | 1.896 | 3.206 |
| Stabilized | 2830403 | 1.010 | 0.231 | 0.100 | 1.002 | 160.877 | 1.052 | 1.093 | 1.260 |
| Stabilized, truncated* | 2830403 | 1.009 | 0.067 | 0.100 | 1.002 | 1.772 | 1.052 | 1.093 | 1.260 |
| Composite w all-cause death | | | | | | | | | |
| Unstabilized | 2825366 | 1.477 | 14.021 | 1.000 | 1.129 | 10789.622 | 1.465 | 1.685 | 2.770 |
| Stabilized | 2825366 | 1.005 | 0.151 | 0.102 | 1.000 | 94.391 | 1.035 | 1.061 | 1.170 |
| Stabilized, truncated* | 2825366 | 1.004 | 0.044 | 0.102 | 1.000 | 1.405 | 1.035 | 1.061 | 1.170 |
| All-cause death | | | | | | | | | |
| Unstabilized | 3207349 | 1.576 | 9.928 | 1.000 | 1.163 | 3643.056 | 1.555 | 1.808 | 3.082 |
| Stabilized | 3207349 | 1.010 | 0.636 | 0.045 | 1.002 | 677.143 | 1.051 | 1.081 | 1.212 |
| Stabilized, truncated* | 3207349 | 1.008 | 0.059 | 0.045 | 1.002 | 1.580 | 1.051 | 1.081 | 1.212 |
| Composite with cardiovascular death | | | | | | | | | |
| Unstabilized | 1636729 | 1.468 | 9.586 | 1.000 | 1.150 | 1901.883 | 1.463 | 1.592 | 2.001 |
| Stabilized | 1636729 | 1.003 | 0.068 | 0.178 | 1.000 | 13.507 | 1.035 | 1.068 | 1.202 |
| Stabilized, truncated* | 1636729 | 1.002 | 0.049 | 0.178 | 1.000 | 1.439 | 1.035 | 1.068 | 1.202 |
| Cardiovascular death | | | | | | | | | |
| Unstabilized | 1850384 | 1.584 | 13.592 | 1.000 | 1.190 | 13123.614 | 1.573 | 1.729 | 2.233 |
| Stabilized | 1850384 | 1.008 | 3.287 | 0.045 | 1.000 | 4287.169 | 1.049 | 1.092 | 1.265 |
| Stabilized, truncated* | 1850384 | 1.004 | 0.066 | 0.045 | 1.000 | 1.618 | 1.049 | 1.092 | 1.265 |
| Hyperkalemia hospitalization | | | | | | | | | |
| Unstabilized | 3080191 | 1.689 | 12.263 | 1.000 | 1.213 | 6047.976 | 1.722 | 2.021 | 3.495 |
| Stabilized | 3080191 | 1.014 | 0.203 | 0.046 | 1.005 | 300.224 | 1.063 | 1.101 | 1.268 |
| Stabilized, truncated* | 3080191 | 1.012 | 0.069 | 0.046 | 1.005 | 1.615 | 1.063 | 1.101 | 1.268 |
| Non-cardiovascular death | | | | | | | | | |
| Unstabilized | 1850432 | 1.603 | 24.030 | 1.000 | 1.195 | 29081.151 | 1.585 | 1.745 | 2.232 |
| Stabilized | 1850432 | 1.012 | 7.265 | 0.046 | 1.000 | 9491.300 | 1.049 | 1.091 | 1.255 |
| Stabilized, truncated* | 1850432 | 1.004 | 0.064 | 0.046 | 1.000 | 1.587 | 1.049 | 1.091 | 1.255 |

Supplemental Table S9. Weight distributions by outcome – overall cohort, 12 months follow-up

| Outcome / Weight type | N** | Mean | Std Dev | Minimum | Median | Maximum | 90th Pctl | 95th Pctl | 99th Pctl |
|---|---|---|---|---|---|---|---|---|---|

*at the 99th percentile

** the number of treatment intervals included in the outcome model (weights are time- and person-specific)

Supplemental Table S10. Comparison of pseudo population and pseudo event counts based on *unstabilized and untruncated weights* with observed population and events over time as a diagnostic of the weight models – overall cohort - Heart failure hospitalizations – clone arm assigned to no-treatment strategy

| interval* | Observed population numbers in interval | Observed outcome events | Observed administrative censoring events | Observed treatment deviations | Pseudo population** numbers at interval start | Outcome pseudo events** | Cumulated outcome pseudo events** in previous intervals | Sum of pseudo population** numbers at interval start and cumulated outcome pseudo events** in previous intervals |
|---|---|---|---|---|---|---|---|---|
| 1 | 225463 | 9061 | 12986 | 2306 | 235912.80 | 9474.78 | 0.00 | 235912.80 |
| 2 | 201110 | 6554 | 9268 | 1468 | 229714.81 | 7497.61 | 9474.78 | 239189.59 |
| 3 | 183820 | 4973 | 7488 | 1075 | 224640.93 | 6126.87 | 16972.38 | 241613.32 |
| 4 | 170284 | 3946 | 6317 | 815 | 219484.62 | 5122.52 | 23099.25 | 242583.88 |
| 5 | 159206 | 3511 | 5687 | 719 | 216322.61 | 4748.35 | 28221.77 | 244544.38 |
| 6 | 149289 | 2955 | 5230 | 553 | 213700.80 | 4181.29 | 32970.12 | 246670.92 |
| 7 | 140551 | 2663 | 4965 | 462 | 211545.39 | 4028.71 | 37151.41 | 248696.80 |
| 8 | 132461 | 2456 | 4533 | 483 | 209491.96 | 3877.59 | 41180.12 | 250672.08 |
| 9 | 124989 | 2166 | 4585 | 417 | 208019.25 | 3685.09 | 45057.70 | 253076.95 |
| 10 | 117821 | 2110 | 5167 | 385 | 205958.94 | 3670.08 | 48742.79 | 254701.73 |
| 11 | 110159 | 1895 | 5966 | 378 | 197558.85 | 3278.65 | 52412.88 | 249971.72 |

*Interval numbering refers to treatment intervals. Due to the lagged data structure, outcome and censoring events in each row represent the number of events recorded in the subsequent interval.

**Pseudo counts shown are based on unstabilized and untruncated weights

Supplemental Table S11. Comparison of pseudo population and pseudo event counts based on *unstabilized and untruncated* weights with observed population and events over time as a diagnostic of the weight models – overall cohort - Heart failure hospitalizations – clone arm assigned to continued spironolactone-treatment strategy

| interval* | Observed population numbers at interval start | Observed outcome events | Observed administrative censoring events | Observed treatment deviations | Pseudo population** numbers at interval start | Outcome pseudo events** | Cumulated outcome pseudo events** in previous intervals | Sum of pseudo population** numbers at interval start and cumulated outcome pseudo events* in previous intervals |
|---|---|---|---|---|---|---|---|---|
| 1 | 241663 | 9553 | 13728 | .0 | 241663.00 | 9553.00 | 0.00 | 241663.00 |
| 2 | 218382 | 7016 | 10085 | 78 | 232357.92 | 7467.26 | 9553.00 | 241910.92 |
| 3 | 201203 | 5398 | 8167 | 2055 | 227503.73 | 6122.17 | 17020.26 | 244523.98 |
| 4 | 185583 | 4218 | 6866 | 727 | 219986.82 | 5010.18 | 23142.42 | 243129.24 |
| 5 | 173772 | 3794 | 6198 | 143985 | 216086.46 | 4687.22 | 28152.60 | 244239.06 |
| 6 | 19795 | 431 | 891 | 676 | 163071.89 | 4610.91 | 32839.82 | 195911.71 |
| 7 | 17797 | 350 | 719 | 510 | 169111.38 | 3169.32 | 37450.73 | 206562.11 |
| 8 | 16218 | 310 | 577 | 552 | 179522.32 | 3425.37 | 40620.05 | 220142.38 |
| 9 | 14779 | 254 | 518 | 417 | 165602.99 | 2561.21 | 44045.43 | 209648.42 |
| 10 | 13590 | 233 | 552 | 337 | 163030.61 | 14685.20 | 46606.64 | 209637.24 |
| 11 | 12468 | 203 | 534 | 262 | 139196.17 | 1266.24 | 61291.83 | 200488.00 |

**Interval numbering refers to treatment intervals. Due to the lagged data structure, outcome and censoring events in each row represent the number of events recorded in the subsequent interval.

*Pseudo counts shown are based on unstabilized and untruncated weights

Supplemental Table S12. Comparison of pseudo population and pseudo event counts based on *stabilized untruncated* weights with observed population and events over time as a diagnostic of the weight models – Heart failure hospitalizations – overall cohort - clone arm assigned to no-treatment strategy

| interval* | Observed population numbers at interval start | Observed outcome events | Observed administrative censoring events | Observed treatment deviations | Pseudo population** numbers at interval start | Outcome pseudo events** |
|---|---|---|---|---|---|---|
| 1 | 225463 | 9061 | 12986 | 2306 | 229115.29 | 9206.81 |
| 2 | 201110 | 6554 | 9268 | 1468 | 207471.38 | 6773.79 |
| 3 | 183820 | 4973 | 7488 | 1075 | 189433.49 | 5162.36 |
| 4 | 170284 | 3946 | 6317 | 815 | 173495.23 | 4044.43 |
| 5 | 159206 | 3511 | 5687 | 719 | 160714.05 | 3546.50 |
| 6 | 149289 | 2955 | 5230 | 553 | 149477.05 | 2967.92 |
| 7 | 140551 | 2663 | 4965 | 462 | 139765.35 | 2669.31 |
| 8 | 132461 | 2456 | 4533 | 483 | 130977.25 | 2430.67 |
| 9 | 124989 | 2166 | 4585 | 417 | 123054.24 | 2161.90 |
| 10 | 117821 | 2110 | 5167 | 385 | 115578.67 | 2083.08 |
| 11 | 110159 | 1895 | 5966 | 378 | 107702.42 | 1856.89 |

**Interval numbering refers to treatment intervals. Due to the lagged data structure, outcome and censoring events in each row represent the number of events recorded in the subsequent interval.

**Pseudo counts shown are based on stabilized untruncated weights

**Supplemental Table S13. Comparison of pseudo population and pseudo event counts based on stabilized untruncated weights with observed population and events over time as a diagnostic of the weight models – overall cohort - Heart failure hospitalizations – clone arm assigned to continued spironolactone-treatment strategy**

| interval* | Observed population numbers at interval start | Observed outcome events | Observed administrative censoring events | Observed treatment deviations | Pseudo population** numbers at interval start | Outcome pseudo events** |
|---|---|---|---|---|---|---|
| 1 | 241663 | 9553 | 13728 | .0 | 241663.00 | 9553.00 |
| 2 | 218382 | 7016 | 10085 | 78 | 220450.35 | 7086.86 |
| 3 | 201203 | 5398 | 8167 | 2055 | 204711.48 | 5503.20 |
| 4 | 185583 | 4218 | 6866 | 727 | 188082.92 | 4287.12 |
| 5 | 173772 | 3794 | 6198 | 143985 | 175404.69 | 3830.62 |
| 6 | 19795 | 431 | 891 | 676 | 21060.39 | 460.88 |
| 7 | 17797 | 350 | 719 | 510 | 19115.31 | 371.94 |
| 8 | 16218 | 310 | 577 | 552 | 17587.06 | 339.94 |
| 9 | 14779 | 254 | 518 | 417 | 16104.78 | 276.83 |
| 10 | 13590 | 233 | 552 | 337 | 14908.70 | 285.62 |
| 11 | 12468 | 203 | 534 | 262 | 13700.91 | 223.32 |

**Interval numbering refers to treatment intervals. Due to the lagged data structure, outcome and censoring events in each row represent the number of events recorded in the subsequent interval.

**Pseudo counts shown are based on stabilized untruncated weights

**Supplemental Table S14. Number of initiators without previous intercurrent events in the last grace period interval – overall cohort**

| Outcome | Number of initiators in month 6 without previous intercurrent events |
|---|---|
| Heart failure hospitalization | 719 |
| Composite with all-cause death | 717 |
| All-cause death | 1156 |
| Composite with cardiovascular death | 398 |
| Cardiovascular death | 644 |
| Hyperkalemia hospitalization | 1156 |
| Non-cardiovascular death | 644 |

Supplemental Table S15. C-statistics for treatment and censoring models – overall cohort

| Outcome / Model | Treatment model - initiation denominator | Treatment model - initiation numerator | Treatment model - continuation denominator | Treatment model - continuation numerator | Censoring model - initiation denominator | Censoring model - initiation numerator | Censoring model - continuation denominator | Censoring model - continuation numerator |
|---|---|---|---|---|---|---|---|---|
| Overall cohort | | | | | | | | |
| HF hospitalization | 0.808 | 0.793 | 0.682 | 0.614 | 0.669 | 0.649 | 0.684 | 0.667 |
| composite with all-cause death | 0.808 | 0.794 | 0.682 | 0.614 | 0.675 | 0.662 | 0.696 | 0.688 |
| all-cause death | 0.791 | 0.749 | 0.673 | 0.612 | 0.664 | 0.648 | 0.683 | 0.672 |
| composite with cardiovascular death | 0.818 | 0.809 | 0.683 | 0.608 | 0.647 | 0.624 | 0.657 | 0.628 |
| cardiovascular death | 0.8 | 0.766 | 0.676 | 0.605 | 0.641 | 0.613 | 0.653 | 0.624 |
| non-cardiovascular death | 0.8 | 0.766 | 0.676 | 0.605 | 0.64 | 0.614 | 0.65 | 0.622 |
| hyperkalemia hospitalization | 0.791 | 0.749 | 0.673 | 0.612 | 0.662 | 0.643 | 0.676 | 0.659 |
| Benchmarking cohort | | | | | | | | |
| HF hospitalization | 0.803 | 0.79 | 0.694 | 0.639 | 0.67 | 0.654 | 0.699 | 0.689 |
| composite with all-cause death | 0.803 | 0.79 | 0.694 | 0.639 | 0.683 | 0.673 | 0.716 | 0.71 |
| all-cause death | 0.791 | 0.747 | 0.684 | 0.634 | 0.672 | 0.661 | 0.703 | 0.692 |
| composite with cardiovascular death | 0.822 | 0.814 | 0.698 | 0.629 | 0.645 | 0.625 | 0.68 | 0.658 |
| cardiovascular death | 0.809 | 0.777 | 0.69 | 0.632 | 0.636 | 0.612 | 0.668 | 0.647 |
| non-cardiovascular death | 0.809 | 0.777 | 0.69 | 0.632 | 0.639 | 0.616 | 0.669 | 0.646 |
| hyperkalemia hospitalization | 0.791 | 0.747 | 0.684 | 0.634 | 0.664 | 0.65 | 0.691 | 0.678 |

**Supplemental Table S16. Absolute 12-months risk estimates by treatment strategy, using stabilized inverse probability of treatment and censoring weights truncated at the 99th percentile – overall cohort**

| Outcome | Non-initiator clones | | | | | Spironolactone user clones | | | | |
|---|---|---|---|---|---|---|---|---|---|---|
| | N pseudo-population at risk | N person-time | N pseudo events | Risk | 95% Confidence limits[a] | N pseudo-population at risk | N person-time | N pseudo events | Risk | 95% Confidence limits[a] |
| Heart failure hospitalization | 229112.9 | 1,724,086 | 42804.57 | 0.227 | (0.225; 0.229) | 229112.9 | 1,131,517 | 32174.98 | 0.228 | (0.225; 0.231) |
| Composite with all-cause death | 228608.3 | 1,716,169 | 64574.18 | 0.322 | (0.320; 0.324) | 228608.3 | 1,120,339 | 48747.07 | 0.326 | (0.323; 0.328) |
| All-cause death | 245100.2 | 1,959,782 | 31695.82 | 0.136 | (0.134; 0.137) | 245100.2 | 1,273,384 | 23978.19 | 0.148 | (0.146; 0.150) |
| Composite with cardiovascular death | 135623.4 | 987,052 | 31576.84 | 0.279 | (0.276; 0.282) | 135623.4 | 653,459 | 24179.03 | 0.272 | (0.267; 0.275) |
| Cardiovascular death | 144918.9 | 1,118,464 | 9719.84 | 0.072 | (0.070; 0.074) | 144918.9 | 739,504 | 7577.04 | 0.077 | (0.074; 0.079) |
| Hyperkalemia hospitalization | 241718.7 | 190,0547 | 13829.38 | 0.067 | (0.066; 0.068) | 241718.7 | 1,215,702 | 11035.38 | 0.082 | (0.080; 0.084) |
| Non-cardiovascular death | 144942.8 | 1,118,544 | 8898.74 | 0.065 | (0.063; 0.066) | 144942.8 | 739,702 | 6828.34 | 0.069 | (0.067; 0.071) |

[a]Confidence limits derived based on bootstrapping with 200 iterations.

**Supplemental Table S17: Weighted outcome analyses results at 12 months, using stabilized inverse probability of treatment and censoring weights truncated at the 99th percentile, before net bias adjustment – overall cohort**

| Outcome | Risk Difference | 95% Confidence limits[a] | Relative Risk | 95% Confidence limits[a] |
|---|---|---|---|---|
| Heart failure hospitalization | 0.001 | (-0.001; 0.003) | 1.004 | (0.997; 1.012) |
| Composite with all-cause death | 0.004 | (0.002; 0.006) | 1.011 | (1.006; 1.018) |
| All-cause death | 0.013 | (0.011; 0.014) | 1.093 | (1.084; 1.102) |
| Composite with cardiovascular death | -0.008 | (-0.011; -0.005) | 0.973 | (0.962; 0.982) |
| Cardiovascular death | 0.005 | (0.003; 0.006) | 1.062 | (1.041; 1.083) |
| Hyperkalemia hospitalization | 0.015 | (0.013; 0.016) | 1.218 | (1.202; 1.237) |
| Non-cardiovascular death | 0.004 | (0.002; 0.005) | 1.057 | (1.037; 1.080) |

[a]Confidence limits derived based on bootstrapping with 200 iterations.

Supplemental Table S18. Baseline characteristics – overall cohort

| | | Spironolactone initiator | | | | | | |
|---|---|---|---|---|---|---|---|---|
| | | All | | No | | Yes | | Standardized |
| | | N = 320,881 (100%) | | N = 291,119 (90.7%) | | N = 29,762 (9.2%) | | Difference |
| Socioeconomic & Sociodemographic characteristics | | | | | | | | |
| | | N | % | N | % | N | % | |
| Index Year | 2013 | 62452 | (19.4) | 57221 | (19.6) | 5231 | (17.4) | -0.05 |
| | 2014 | 51826 | (16.1) | 47214 | (16.2) | 4612 | (15.3) | -0.02 |
| | 2015 | 48105 | (14.9) | 43827 | (15.0) | 4278 | (14.2) | -0.02 |
| | 2016 | 33829 | (10.5) | 30512 | (10.4) | 3317 | (11.0) | 0.01 |
| | 2017 | 31777 | (9.9) | 28653 | (9.8) | 3124 | (10.4) | 0.02 |
| | 2018 | 33044 | (10.2) | 29717 | (10.2) | 3327 | (11.0) | 0.02 |
| | 2019 | 32923 | (10.2) | 29461 | (10.1) | 3462 | (11.5) | 0.04 |
| | 2020 | 26925 | (8.3) | 24275 | (8.3) | 2650 | (8.8) | 0.01 |
| Sex | female | 199127 | (62.0) | 181857 | (62.5) | 17270 | (57.5) | -0.08 |
| Race | missing | 12847 | (4.0) | 11827 | (4.0) | 1020 | (3.3) | -0.03 |
| | White | 269705 | (84.0) | 244291 | (83.9) | 25414 | (84.7) | -0.02 |
| | Black | 31532 | (9.8) | 28675 | (9.8) | 2857 | (9.5) | -0.01 |
| | Other | 6797 | (2.1) | 6087 | (2.0) | 710 | (2.3) | -0.02 |
| Region | northeast | 67246 | (20.9) | 61702 | (21.2) | 5544 | (18.4) | -0.06 |
| | midwest | 78195 | (24.3) | 70318 | (24.1) | 7877 | (26.2) | 0.04 |
| | south | 130082 | (40.5) | 118120 | (40.6) | 11962 | (39.8) | -0.01 |
| | unknown | 579 | (0.1) | 507 | (0.1) | 72 | (0.2) | 0.01 |
| | west | 44779 | (13.9) | 40233 | (13.8) | 4546 | (15.1) | 0.03 |
| Low Income Subsidy Recipient | | 102521 | (31.9) | 93821 | (32.2) | 8700 | (28.9) | -0.06 |
| Urban area | | 7951 | (2.4) | 7170 | (2.4) | 781 | (2.6) | 0.01 |
| Socioeconomic & Sociodemographic characteristics | | | | | | | | |
| | | Mean | (SD) | Mean | (SD) | Mean | (SD) | |
| Baseline age | | 80.55 | (8.37) | 80.76 | (8.38) | 78.48 | (7.90) | **-0.28** |
| Area deprivation index | | 56.70 | (7.27) | 56.70 | (7.27) | 56.72 | (7.28) | 0.00 |
| Combined Comorbidity Index | | 6.82 | (2.71) | 6.83 | (2.71) | 6.75 | (2.69) | -0.03 |
| Claims-based Frailty Index | | 0.23 | (0.04) | 0.23 | (0.04) | 0.23 | (0.04) | **-0.16** |
| Predicted probability of HFpEF | | 0.91 | (0.04) | 0.91 | (0.04) | 0.90 | (0.04) | **-0.11** |
| Healthcare utilization (continuous variables) – 365 days pre-index | | | | | | | | |
| Number of outpatient visits | | 13.84 | (9.55) | 13.77 | (9.53) | 14.58 | (9.75) | 0.08 |
| Number of prescribers | | 5.30 | (3.43) | 5.29 | (3.42) | 5.38 | (3.49) | 0.02 |
| Number of hospitalizations | | 1.75 | (1.15) | 1.75 | (1.16) | 1.72 | (1.11) | -0.03 |
| Number of emergency room visits | | 2.52 | (2.27) | 2.53 | (2.28) | 2.41 | (2.16) | -0.05 |
| Number of cardiologist visits | | 6.61 | (7.36) | 6.56 | (7.31) | 7.12 | (7.76) | 0.07 |

| | | | | | | | |
|---|---|---|---|---|---|---|---|
| Healthcare utilization (continuous time-varying variables) – 30 days pre-index | | | | | | | |
| Number of outpatient visits | 1.36 | (1.38) | 1.34 | (1.38) | 1.50 | (1.45) | **0.12** |
| Number of hospitalizations | 1.04 | (0.48) | 1.03 | (0.48) | 1.07 | (0.42) | 0.08 |
| Number of emergency room visits | 1.03 | (0.70) | 1.03 | (0.71) | 1.02 | (0.68) | -0.02 |
| Number of cardiologist visits | 1.27 | (1.75) | 1.26 | (1.75) | 1.37 | (1.79) | 0.06 |
| Number of HF hospitalizations | 0.91 | (0.28) | 0.90 | (0.29) | 0.95 | (0.21) | **0.18** |
| Number of prescribed medications | 4.87 | (3.58) | 4.87 | (3.57) | 4.88 | (3.59) | 0.00 |
| Number of prescribers | 1.84 | (1.43) | 1.84 | (1.43) | 1.85 | (1.46) | 0.01 |
| Number of outpatient physicians | 5.49 | (4.09) | 5.47 | (4.10) | 5.68 | (4.03) | 0.05 |
| Healthcare utilization for preventive measures – 365 days pre index | | | | | | | |
| | N | % | N | % | N | % | |
| Bone mineral density testing | 19580 | (6.1) | 17667 | (6.0) | 1913 | (6.3) | -0.01 |
| Colonoscopy | 24285 | (7.5) | 21566 | (7.4) | 2719 | (9.0) | -0.05 |
| Flu vaccination | 195595 | (60.9) | 177399 | (60.9) | 18196 | (60.6) | 0.01 |
| Pneumococcal vaccination | 45024 | (14.0) | 40659 | (13.9) | 4365 | (14.5) | -0.01 |
| Herpes zoster vaccination | 206 | (0.0) | 191 | (0.0) | 15 | (0.0) | 0.01 |
| Mammography | 44560 | (13.8) | 39978 | (13.7) | 4582 | (15.2) | -0.04 |
| PAP/HPV test | 7206 | (2.2) | 6481 | (2.2) | 725 | (2.4) | -0.01 |
| Fecal occult blood test | 20708 | (6.4) | 18606 | (6.3) | 2102 | (7.0) | -0.02 |
| Prostate specific antigen test | 17530 | (5.4) | 15476 | (5.3) | 2054 | (6.8) | -0.05 |
| Medications (time-varying variables) 30 days pre-index | | | | | | | |
| Angiotensin-converting enzyme inhibitors | 45137 | (14.0) | 41017 | (14.1) | 4120 | (13.7) | -0.01 |
| Angiotensin receptor blockers | 40562 | (12.6) | 36609 | (12.5) | 3953 | (13.1) | 0.01 |
| Beta blockers | 118597 | (36.9) | 107648 | (37.0) | 10949 | (36.4) | -0.01 |
| Calcium channel blockers | 69381 | (21.6) | 63389 | (21.7) | 5992 | (19.9) | -0.04 |
| Loop diuretics | 112124 | (34.9) | 101245 | (34.8) | 10879 | (36.2) | 0.02 |
| Thiazide diuretics | 17544 | (5.4) | 15555 | (5.3) | 1989 | (6.6) | 0.04 |
| Hydralazine | 14583 | (4.5) | 13330 | (4.5) | 1253 | (4.1) | -0.02 |
| Nitrates | 27768 | (8.6) | 25312 | (8.7) | 2456 | (8.1) | -0.02 |
| Digoxin | 12345 | (3.8) | 11188 | (3.8) | 1157 | (3.8) | 0.00 |
| Sacubitril/Valsartan | 509 | (0.1) | 442 | (0.1) | 67 | (0.2) | 0.01 |
| SGLT2 inhibitors | 791 | (0.2) | 693 | (0.2) | 98 | (0.3) | 0.01 |
| Comorbidities 365 days pre index | | | | | | | |
| COPD | 152813 | (47.6) | 138876 | (47.7) | 13937 | (46.4) | -0.02 |
| Pulmonary disease | 145776 | (45.4) | 131627 | (45.2) | 14149 | (47.1) | 0.03 |
| Valve disorder | 196859 | (61.3) | 177878 | (61.1) | 18981 | (63.2) | 0.04 |
| Cardiac arrest | 2578 | (0.8) | 2418 | (0.8) | 160 | (0.5) | -0.03 |
| Atrial fibrillation | 191079 | (59.5) | 172935 | (59.4) | 18144 | (60.4) | 0.02 |

| | | | | | | | |
|---|---|---|---|---|---|---|---|
| Obesity | 116079 | (36.1) | 103686 | (35.6) | 12393 | (41.3) | **0.10** |
| Diabetes | 187418 | (58.4) | 168801 | (58.0) | 18617 | (62.0) | 0.07 |
| Pulmonary hypertension | 38212 | (11.9) | 34164 | (11.7) | 4048 | (13.4) | 0.04 |
| Renal disease | 209620 | (65.3) | 191063 | (65.6) | 18557 | (61.8) | -0.07 |
| Coronary artery disease | 221398 | (68.9) | 199745 | (68.6) | 21653 | (72.1) | 0.06 |
| Angioedema | 675 | (0.2) | 619 | (0.2) | 56 | (0.1) | -0.00 |
| Anemia | 177893 | (55.4) | 161713 | (55.5) | 16180 | (53.9) | -0.03 |
| Depression | 74750 | (23.2) | 67829 | (23.3) | 6921 | (23.0) | -0.00 |
| Diabetic nephropathy | 62709 | (19.5) | 56957 | (19.5) | 5752 | (19.1) | -0.01 |
| Endocarditis | 9931 | (3.0) | 8931 | (3.0) | 1000 | (3.3) | 0.01 |
| Hypertensive nephropathy | 140331 | (43.7) | 128706 | (44.2) | 11625 | (38.7) | -0.09 |
| Hyperlipidemia | 272281 | (84.8) | 246333 | (84.6) | 25948 | (86.4) | 0.04 |
| Hypertension | 314091 | (97.8) | 284736 | (97.8) | 29355 | (97.8) | -0.00 |
| Hypokalemia | 79083 | (24.6) | 69967 | (24.0) | 9116 | (30.3) | **0.12** |
| Hypotension | 47615 | (14.8) | 43433 | (14.9) | 4182 | (13.9) | -0.02 |
| Other dysrhythmias | 146233 | (45.5) | 132143 | (45.4) | 14090 | (46.9) | 0.03 |
| Peripheral artery disease | 38217 | (11.9) | 34826 | (11.9) | 3391 | (11.3) | -0.02 |
| Psychosis | 25379 | (7.9) | 23681 | (8.1) | 1698 | (5.6) | -0.08 |
| Rheumatic heart disease | 47164 | (14.6) | 42691 | (14.6) | 4473 | (14.9) | 0.01 |
| Sleep apnea | 77011 | (23.9) | 68313 | (23.4) | 8698 | (28.9) | **0.10** |
| Stable angina | 27759 | (8.6) | 25028 | (8.6) | 2731 | (9.1) | 0.01 |
| Smoking | 93629 | (29.1) | 84789 | (29.1) | 8840 | (29.4) | 0.01 |
| Thyroid dysfunction | 123239 | (38.4) | 111913 | (38.4) | 11326 | (37.7) | -0.01 |
| Unstable angina | 32182 | (10.0) | 29136 | (10.0) | 3046 | (10.1) | 0.00 |
| Hemorrhage | 3343 | (1.0) | 3049 | (1.0) | 294 | (0.9) | -0.01 |
| Outpatient HF index diagnosis | 310746 | (96.8) | 281378 | (96.7) | 29368 | (97.8) | 0.06 |
| Metastatic cancer | 10786 | (3.3) | 9890 | (3.4) | 896 | (2.9) | -0.02 |
| Cardiomyopathy | 18505 | (5.7) | 16162 | (5.5) | 2343 | (7.8) | -0.08 |
| Home oxygen | 48285 | (15.0) | 44052 | (15.1) | 4233 | (14.1) | -0.02 |
| Hospitalization for exacerbation | 50787 | (15.8) | 46657 | (16.0) | 4130 | (13.7) | -0.05 |
| Orthostatic hypotension | 11449 | (3.5) | 10498 | (3.6) | 951 | (3.1) | -0.02 |
| Severe renal disease | 34853 | (10.8) | 32678 | (11.2) | 2175 | (7.2) | **-0.11** |
| Hepatic disease | 37747 | (11.7) | 32723 | (11.2) | 5024 | (16.7) | **0.13** |
| Gastrointestinal disorder | 22497 | (7.0) | 20229 | (6.9) | 2268 | (7.5) | 0.02 |
| Alcohol (ab)use | 7800 | (2.4) | 6922 | (2.3) | 878 | (2.9) | 0.03 |
| Drug (ab)use | 8623 | (2.6) | 7827 | (2.6) | 796 | (2.6) | -0.00 |
| Alzheimer's/Dementia | 44514 | (13.8) | 41691 | (14.3) | 2823 | (9.4) | **-0.12** |
| MI (365-91 days pre-index) | 4842 | (1.5) | 4365 | (1.5) | 477 | (1.5) | 0.01 |
| Stroke (365-91 days pre-index) | 3653 | (1.1) | 3306 | (1.1) | 347 | (1.1) | 0.00 |
| MI (90 days pre-index) | 2655 | (0.8) | 2396 | (0.8) | 259 | (0.8) | --0.00 |
| Stroke (90 days pre-index) | 1686 | (0.5) | 1566 | (0.5) | 120 | (0.3) | -0.02 |
| Procedures | | | | | | | |
| CABG (90 days pre-index) | 1602 | (0.4) | 1396 | (0.4) | 206 | (0.6) | -0.02 |
| PCI (30 days pre-index) | 2260 | (0.7) | 1983 | (0.6) | 277 | (0.9) | -0.02 |

| | | | | | | | |
|---|---|---|---|---|---|---|---|
| Cardioverter defibrillator (365 days pre-index) | 10792 | (3.3) | 9306 | (3.1) | 1486 | (4.9) | -0.08 |
| Cardiac resynchronization therapy (365 days pre-index) | 12975 | (4.0) | 11246 | (3.8) | 1729 | (5.7) | -0.07 |
| Valve correction (365 days pre-index) | 1725 | (0.5) | 1517 | (0.5) | 208 | (0.6) | -0.02 |
| Valve replacement (365 days pre-index) | 3557 | (1.1) | 3124 | (1.0) | 433 | (1.4) | -0.03 |
| CABG (365-91 days pre-index) | 1324 | (0.4) | 1129 | (0.3) | 195 | (0.6) | -0.03 |
| PCI (365-31 days pre-index) | 9023 | (2.8) | 8076 | (2.7) | 947 | (3.1) | -0.02 |
| Further medications - 30 days pre-index | | | | | | | |
| Statins | 85914 | (26.7) | 77990 | (26.8) | 7924 | (26.4) | -0.01 |
| Insulin | 27582 | (8.5) | 24817 | (8.5) | 2765 | (9.2) | 0.02 |
| Hypoglycemic agents | 51242 | (15.9) | 45946 | (15.7) | 5296 | (17.6) | 0.04 |
| Antiplatelets | 25427 | (7.9) | 23106 | (7.9) | 2321 | (7.7) | -0.01 |
| Anticoagulants | 59746 | (18.6) | 53652 | (18.4) | 6094 | (20.3) | 0.04 |
| Gout medications | 15811 | (4.9) | 14309 | (4.9) | 1502 | (5.0) | 0.00 |
| COPD medications | 38641 | (12.0) | 23447 | (8.0) | 2572 | (8.5) | 0.01 |
| NSAIDs | 17072 | (5.3) | 34951 | (12.0) | 3690 | (12.3) | -0.00 |
| Pressor amines | 157 | (0.0) | 138 | (0.0) | 19 | (0.0) | 0.01 |
| Barbiturates | 181 | (0.0) | 168 | (0.0) | 13 | (0.0) | -0.01 |
| Narcotics | 53855 | (16.7) | 48896 | (16.8) | 4959 | (16.5) | -0.01 |
| Skeletal muscle relaxants | 5893 | (1.8) | 5342 | (1.8) | 551 | (1.8) | 0.00 |
| Oral steroids | 5536 | (1.7) | 4992 | (1.7) | 544 | (1.8) | 0.01 |
| Further medications - 365-31 days pre-index | | | | | | | |
| Statins | 200064 | (62.3) | 180987 | (62.2) | 19077 | (63.5) | 0.02 |
| Insulin | 55951 | (17.4) | 50328 | (17.3) | 5623 | (18.7) | 0.03 |
| Hypoglycemic agents | 105652 | (32.9) | 94572 | (32.5) | 11080 | (36.9) | 0.08 |
| Antiplatelets | 57305 | (17.8) | 51862 | (17.8) | 5443 | (18.1) | 0.01 |
| Anticoagulants | 112985 | (35.2) | 101617 | (34.9) | 11368 | (37.8) | 0.05 |
| Gout medications | 35764 | (11.1) | 32337 | (11.1) | 3427 | (11.4) | 0.01 |
| COPD medications | 85775 | (26.7) | 77703 | (26.6) | 8072 | (27.1) | 0.01 |
| NSAIDs | 60169 | (18.7) | 54263 | (18.6) | 5906 | (19.6) | 0.02 |
| Pressor amines | 1149 | (0.3) | 1020 | (0.3) | 129 | (0.4) | 0.01 |
| Barbiturates | 296 | (0.0) | 273 | (0.0) | 23 | (0.0) | -0.00 |
| Narcotics | 123630 | (38.5) | 111725 | (38.4) | 11905 | (39.6) | 0.02 |
| Skeletal muscle relaxants | 21823 | (6.8) | 19564 | (6.7) | 2259 | (7.5) | 0.03 |
| Calcium channel blockers | 152308 | (47.4) | 138445 | (47.5) | 13863 | (46.2) | -0.02 |
| Oral steroids | 29139 | (9.0) | 26328 | (9.0) | 2811 | (9.3) | 0.01 |

**Supplemental Table S19: Weighted outcome analyses results at 12 months, using stabilized inverse probability of treatment and censoring weights truncated at the 99th percentile, before net bias adjustment – benchmarking cohort**

| Outcome | Risk Difference | 95% Confidence limits[a] | | Relative Risk | 95% Confidence limits[a] | |
|---|---|---|---|---|---|---|
| Heart failure hospitalization | 0.002 | (-0.002; | 0.006) | 1.010 | (0.990; | 1.038) |
| Composite with all-cause death | 0.005 | (0.000; | 0.009) | 1.022 | (1.002; | 1.039) |
| All-cause death | 0.011 | (0.008; | 0.014) | 1.141 | (1.107; | 1.176) |
| Composite with cardiovascular death | -0.006 | (-0.013; | 0.001) | 0.971 | (0.938; | 1.007) |
| Cardiovascular death | 0.003 | (0.000; | 0.007) | 1.080 | (1.011; | 1.158) |
| Hyperkalemia hospitalization | 0.014 | (0.011; | 0.017) | 1.290 | (1.230; | 1.351) |
| Non-cardiovascular death | 0.004 | (0.001; | 0.007) | 1.102 | (1.022; | 1.187) |

[a]Confidence limits derived based on bootstrapping with 200 iterations.

**Supplemental Table S20: Parameters for assessing the probability of estimate agreement – benchmarking cohort**

| | TOPCAT logarithmised estimate | | logarithmised estimate from the benchmarking cohort | | Estimate agreement | | Z[3] score | |
|---|---|---|---|---|---|---|---|---|
| Outcome | lower bound | Upper bound | without net bias adjustment | with net bias adjustment using non cardio-vascular death | without net bias adjustment | with net bias adjustment using non-cardiovascula r death | without net bias adjustment | with net bias adjustment using non cardio-vascular death |
| Heart failure hospitalization[1] | -0.24846 | 0.05827 | 0.01034 | -0.08692 | yes | yes | -1.32152 | 0.09327 |
| All-cause death[2] | -0.38566 | 0.01980 | 0.13211 | 0.03484 | no | no | -2.44721 | 1.36611 |
| Composite outcome with cardiovascular death[2] | -0.37106 | -0.02020 | -0.02983 | -0.12710 | yes | yes | -1.84680 | 0.78146 |
| Cardiovascular death[2] | -0.56212 | -0.03046 | 0.07739 | -0.01988 | no | no | -2.70377 | 2.00893 |

*expressed as the difference between the targeted logarithmised effect parameters from TOPCAT vs benchmarking cohort in units of standard errors of the TOPCAT estimate

[1]based on hospitalization stratum in the Americas

[2]based on the Americas, no further sub-stratification by randomization stratum available

[3]Z values <1.96 and > -1.96 indicate no statistically significant difference and Z>1.96 or <-1.96 indicate a significant difference

### References

1. Webster-Clark M, Li Y, Aniello SD, Platt RW. The Complex Estimand of Clone-Censor-Weighting When Studying Treatment Initiation Windows. *arXiv preprint arXiv:240415073*. 2024.
2. Hernan MA, Robins JM. *Causal Inference: What if*. Boca Raton: Chapman & Hall/CRC; 2020.
3. Pfeffer MA, Claggett B, Assmann SF, Boineau R, Anand IS, Clausell N, Desai AS, Diaz R, Fleg JL, Gordeev I. Regional variation in patients and outcomes in the Treatment of Preserved Cardiac Function Heart Failure With an Aldosterone Antagonist (TOPCAT) trial. *Circulation*. 2015;131:34–42.
4. Pitt B, Pfeffer MA, Assmann SF, Boineau R, Anand IS, Claggett B, Clausell N, Desai AS, Diaz R, Fleg JL. Spironolactone for heart failure with preserved ejection fraction. *New England Journal of Medicine*. 2014;370:1383–1392.
5. Szabo B, Benson L, Savarese G, Hage C, Fudim M, Devore A, Pitt B, Lund LH. Previous heart failure hospitalization, spironolactone and outcomes in heart failure with preserved ejection fraction–a secondary analysis of TOPCAT. *American Heart Journal*. 2024.
